# Orphanhood and caregiver death among children in the United States due to all-cause mortality 2000-2021: A Modeling Study

**DOI:** 10.1101/2024.03.25.24304835

**Authors:** Andrés Villaveces, Yu Chen, Sydney Tucker, Alexandra Blenkinsop, Lucie Cluver, Lorraine Sherr, Jan L. Losby, Linden Graves, Rita Noonan, Francis Annor, Victor Kojey-Merle, Douhan Wang, Greta Massetti, Laura Rawlins, Charles A. Nelson, H Juliette T Unwin, Seth Flaxman, Susan Hillis, Oliver Ratmann

**Affiliations:** National Center for Injury Prevention and Control, Centers for Disease Control and Prevention, Atlanta, Georgia, USA; Department of Mathematics, Imperial College London, London, UK; Centre for Evidence-Based Social Intervention, Department of Social Policy and Intervention, University of Oxford, Oxford, UK; Institute of Global Health, University College London, London, UK; Gender Group, World Bank, Washington DC, USA; Harvard Medical School and Boston Children’s Hospital, Harvard University, Boston, MA, USA; School of Mathematics, University of Bristol, Bristol, UK; Department of Computer Science, University of Oxford, Oxford, UK; Global Reference Group on Children Affected by COVID-19 and in Crisis, University of Oxford, Oxford, UK

## Abstract

**Importance:** Deaths of parents and grandparent caregivers linked to social and health crises threaten child wellbeing due to losses of nurturance, financial support, physical safety, family stability, and care. Little is known about the full burden of all-causes and leading cause-specific orphanhood and caregiver death beyond estimates from select causes.

**Objective:** To estimate 2000-2021 prevalence and incidence trends of all-cause orphanhood and caregiver death among children <18, by cause, age, race/ethnicity, and state.

**Data Sources:** National Center for Health Statistics (NCHS) birth, death, race/ethnicity, and population data to estimate fertility rates and identify causes of death; 1983-1998 ICD-9 causes-of-death harmonized to ICD-10 classifications; 1999-2021 ICD-10 causes-of-death; CDC WONDER for state-specific estimates; and American Community Survey for grandparent population estimates.

**Data extraction and synthesis:** We extracted U.S. population-level death, birth, population size, race, and ethnicity data from NCHS and attributed to each deceased individual the average number of children left behind according to subgroup-specific fertility rates in the previous 0-17 years. We examined prevalence and incidence of orphanhood by leading causes-of-death, including COVID-19, the leading 5 causes-of-death for 1983-2021, and additional leading causes for ages 15-44. We extended these to obtain state-level outcome estimates.

**Main outcome measures:** National incidence and prevalence of orphanhood and caregiver death from 2000-2021, with orphanhood by year, parental cause-of-death and sex, child age, race/ethnicity, and state.

**Results:** From 2000-2021, orphanhood and custodial/co-residing grandparent caregiver loss annual incidence and prevalence trends increased 49.2% and 8.3%, respectively. By 2021, 2.9 million children (4% of all children) had experienced prevalent orphanhood and caregiver death. Populations disproportionately affected by orphanhood included 5.0% of all adolescents; 6.5%, 4.8%, and 3.9% respectively of non-Hispanic American Indian/Alaska Native, non-Hispanic Black, and non-Hispanic White children; and children in New Mexico and Southern and Eastern States. Parental death due to drug overdose during 2020-2021 surpassed COVID-19 as the leading cause of incident and prevalent orphanhood during the COVID-19 pandemic.

**Conclusions and Relevance:** Policies, programs, and practices aimed at orphanhood prevention, identification, and linkage to services and support of nearly 3 million bereaved children are needed, foremost prioritizing rapidly increasing overdose-linked orphanhood.

**Key Point:** **Question**: What are U.S. trends in all-cause and cause-specific orphanhood and caregiver death among children <18?

**Findings**: From 2000-2021, orphanhood and caregiver loss incidence and prevalence increased 49.2% and 8.3%, respectively. By 2021, 2.9 million children (4% of all children) were affected. Populations disproportionately affected by orphanhood included 1.7 million adolescents ages 10-17; 6.5%, 4.8%, and 3.9% respectively of non-Hispanic American Indian/Alaska Native, non-Hispanic Black, and non-Hispanic White children; and children in New Mexico, Southern and Eastern States. Drug overdose was the leading cause of orphanhood during the COVID-19 pandemic.

**Meaning**: Evidence-based programs and policies are needed to prevent orphanhood and support these bereaved children.

## Introduction

Deaths of parents and co-residing/custodial grandparent caregivers, in the context of intersecting social and health crises, threaten child wellbeing^1^ due to losses of nurturance, financial support, housing, and care.^2–4^ Orphanhood (death of one or both parents)^5,6^ and caregiver loss (death of co-resident/custodial grandparents) are considered Adverse Childhood Experiences (ACEs),^7,8^ and may have enduring consequences that persist well into adulthood.^9–12^ ACE’s are associated with increased chronic stress and lifelong risks of mental health illness, suicide, post-traumatic stress disorder, violence, insecure housing,^13^ and chronic and infectious diseases.^14,15^ These impacts often lead to ongoing needs for mental health, parenting, educational, and economic support for affected children, and for foster care or adoption services for children bereft of care.^15^

In the United States, mortality data informs public health prevention and response policies according to leading causes-of-death.^16^ However, one disproportionately affected population often remains largely invisible – those children left behind by adult deaths.^2,6,17^ One exception are children experiencing COVID-19-associated orphanhood, where real-time incidence estimates^18–20^ led to policies to support children, including recommendations in the National COVID-19 Pandemic Preparedness Plan^21^ and investments in some states,^22^ such as financial, bereavement, and mental health support for bereaved children and surviving caregivers. These established frameworks raise the possibility that new standards of care can be extended for any child experiencing orphanhood, regardless of cause.^23^

To inform evidence-based, compassionate prevention and response strategies for affected children and families, understanding the numbers, time-trends, causes, locations and disparities in all-cause orphanhood is essential. Reports show COVID-19 excess deaths caused a surge in orphanhood and caregiver death, affecting over 10.5 million children globally.^24^ In the U.S., the pandemic was linked to increased challenges including substance use, economic crises, mental health distress, and may have been linked to deaths due to overdose, suicide, and excessive alcohol use.^25,26^ Though reports have described the U.S. orphanhood burden related to HIV,^27^ maternal cancers,^23^ COVID-19,^15^ and child bereavement,^28^ up-to-date estimates on all-cause orphanhood do not exist.

Therefore, we estimate prevalence, incidence, and trends from 2000-2021 in all-cause orphanhood and caregiver death among children in the U.S. We further characterize leading causes of orphanhood and caregiver loss and identify whether the compound crises of the COVID-19 pandemic and drug overdose epidemic in 2020 and 2021 were associated with escalating orphanhood. We aim to identify populations disproportionately affected, by age, race/ethnicity, and state, to strengthen evidence-based response strategies. Our findings can contribute to identifying children at risk among population subgroups and geographic areas, and to targeting effective prevention and protection programming for affected children and parents/caregivers.

## Methods

### Overview

We adapted methods from Hillis et al^19^ to estimate the magnitude of orphanhood and caregiver loss among children ages 0-17 from all causes of deaths, according to Guidelines for Accurate and Transparent Health Estimates Reporting (GATHER). We extracted subgroup-specific individual live birth and death data from National Center for Health Statistics (NCHS), along with population size estimates. We then attributed to each deceased individual the average number of children orphaned using subgroup-specific fertility rates in the previous 0-17 years. Our outcomes were orphanhood incidence and prevalence from 2000 through 2021, disaggregated by year, child age, and sex, race/ethnicity, cause-of-death of the deceased parent (numbers and percent of child population). We extended these estimates to include loss of co-residing/custodial grandparent caregivers, and state-level estimates of incidence and prevalence of orphanhood and grandparent caregiver loss (Supplementary Text (ST) S1, Fig. 2). We calculated 95% uncertainty intervals around estimates using bootstrap resampling (ST S1).

### National mortality data

We extracted mortality data of U.S. residents by underlying cause-of-death from NCHS Vital Statistics for 1983-2021.^29^ The World Health Organization (WHO) defines underlying cause-of-death as “the disease or injury which initiated the train of events leading directly to death, or the circumstances of the accident or violence which produced the fatal injury”.^30^ Mortality records^29^ were mapped and aggregated to one of 53 caregiver loss causes-of-death that are closely related to the 52 rankable causes-of-death in the NCHS 113 cause-of-death list^31^, or ‘Other’ caregiver loss cause-of-death (ST S1). Cause-of-death classifications changed with the Tenth Revision of the International Classification of Disease (ICD-10), and we harmonized 1983-1998 cause-of-death data to ICD-10 classifications to avoid discontinuities in orphanhood estimates from 1998-1999 (ST S1).

Our cause-specific analyses for orphanhood estimates focused on leading causes of death in 2021: COVID-19, drug overdose, the remaining top five causes-of-death overall (heart disease, cancer, unintentional injuries, cerebrovascular diseases, chronic lung disease), and any additional causes of death in the top five for men and women ages 15-44 (suicide and homicide), as these ages include those more likely to be parents. We used a hierarachical approach for classifying cause of death; therefore, we consistently excluded drug overdose from each of these three categories: suicide, homicide, and unintentional injuries. Race and ethnicity in death certificates are typically reported by next of kin. We converted data that were incrementally reported by multiple races to standardised race categories (Table S7).

### National orphanhood estimates

To estimate the number of children each death leaves behind, we obtained age, race, and ethnicity data of fathers and mothers from NCHS live birth records.^29,31^ We used corresponding 1990-2021 population sizes^32^ to calculate annualized age- and sex-specific national-level fertility rates by standardised race/ethnicity categories (ST S1). Before 1990, we assumed fertility rates of 1990 because population sizes by race/ethnicity were unavailable before 1990, and report corresponding sensitivity analyses (ST S2).

### Incidence

We estimated annual incidence of orphanhood for a given year among children by multiplying the number of cause-specific deaths among adults stratified by sex, race/ethnicity, and five-year age bands in each year 1983-2021, with the expected number of children aged 0-17 born to each adult, with the corresponding characteristics in the same year. Calculations were performed by race/ethnicity groups and then aggregated to U.S. totals due to correlations between mortality and fertility rates by race/ethnicity (ST S1). Age of child was attributed by considering fertility rates in each of the current and preceding 17 years, after adjustment for pediatric survival probabilities (Table S4).

### Prevalence

We estimated annual prevalence of orphanhood among children in any given year in 2000-2021 by cumulatively summing incidence estimates by one-year age of child in the current and previous 17 years, excluding children who would have since turned 18; calculations of orphanhood prevalence for 2000-2015 required including annual orphanhood incidence estimates for 1983-1999, in order to include the cumulative prior 17-year risk for experiencing prevalent parental death. We removed duplicate orphanhood counts by adjusting for those who had lost both parents (ST 1, Table S5). Incidence and prevalence rates were calculated relative to child population sizes in the corresponding year.

### National grandparent caregiver loss

We estimated annual incidence of children losing grandparent caregivers in 1983-2021 by multiplying the number of cause-specific deaths in each year in adults aged 30 years and above stratified by sex and race/ethnicity with the corresponding proportions of adults co-residing with their grandchild(ren) and providing some or all basic needs for grandchild(ren), using U.S. Census American Community Surveys (ACS) 2010-2021 data (ST S1).^33^ We assumed only one child was affected by each grandparent caregiver death. We also developed estimates for the age of children affected (Fig S21).

### State-level orphanhood and grandparent caregiver death estimates

We generated state-level estimates of cause-specific incidence and prevalence of orphanhood and caregiver death for 2021, using data from CDC WONDER.^34,35^ (ST S1). State-level orphanhood and caregiver death were calculated as above, with exceptions for male fertility rates for 2005-2015 (ST S1). We developed correction factors to adjust for bias resulting from data not stratified by race and ethnicity (ST S1).

## Results

### All-cause orphanhood and caregiver loss

In 2021, we estimate 498,249 (95% CI, 496,938-499,639) childr experienced incident orphanhood or caregiver death from any cause (0.72% of children, Table 1, Fig. 2). Most children lost a parent (81.6%), while the remainder lost a grandparent caregiver (18.4%). Before the COVID-19 pandemic, incidence of orphanhood, and of orphanhood/caregiver loss (combined) increased rapidly from 2000 to 2019 by 11.4% and 11.7%, respectively (Table 1). Through the pandemic, incidence of orphanhood, and of orphanhood/caregiver loss (combined) increased further from 2019 to 2021 by 39.5% and 33.6%, respectively. Prevalence of orphanhood, and of orphanhood/caregiver loss decreased slightly by 2.4% and 0.9% from 2000 to 2019, then increased by 10.1% and 9.3% during the period spanning the pandemic years (2019 to 2021). In 2021, we estimate 2,966,008 (95%CI, 2,963,030-2,968,931) children experienced prevalent orphanhood or caregiver death, representing 4.0% of all children.

**Table 1.** Trends in All-cause Orphanhood and Caregiver (Co-Residing/custodial Grandparent) Loss from 2000 to 2021, before and during the COVID-19 pandemic.

|  |  | Before the COVID-19 pandemic |  |  | Since the COVID-19 pandemic |  |  |  |
| --- | --- | --- | --- | --- | --- | --- | --- | --- |
|  |  | 2000<br>(Number<br>of<br>Children) | 2019<br>(Number<br>of<br>Children) | 2000-2019<br>change | 2020<br>(Number<br>of<br>Children) | 2021<br>(Number<br>of<br>Children) | 2019-2021<br>change | 2000-2021<br>change |
| **Incidence (n, (95% uncertainty interval))** |  |  |  |  |  |  |  |  |
|  | **Orphanhood** | 261,633<br>(260,485,<br>262,656) | 291,574<br>(290,613,<br>292,657) | +11.4%<br>(+10.9%,<br>+12.1%) | 359,431<br>(358,301,<br>360,531) | 406,733<br>(405,437,<br>407,928) | +39.5%<br>(+38.8%,<br>+40.1%) | +55.5%<br>(+54.7%,<br>+56.3%) |
|  | **Orphanhood**<br>**and**<br>**Caregiver**<br>**Loss** | 333,959<br>(332,701,<br>335,049) | 372,869<br>(371,737,<br>374,132) | +11.7%<br>(+11.2%,<br>+12.2%) | 454,403<br>(453,107,<br>455,706) | 498,249<br>(496,938,<br>499,639) | +33.6%<br>(+33.1%,<br>+34.2%) | +49.2%<br>(+48.5%,<br>+49.9%) |
| **Incidence rate per 100 children (rate, (95% uncertainty interval))** |  |  |  |  |  |  |  |  |
|  | **Orphanhood** | 0.36 (0.36,<br>0.36) | 0.40 (0.40,<br>0.40) | +10.2%<br>(+9.9%,<br>+11.1%) | 0.49 (0.49,<br>0.50) | 0.59 (0.58,<br>0.59) | +46.9%<br>(+46.2%,<br>+47.6%) | +62.2%<br>(+61.3%,<br>+63.1%) |
|  | **Orphanhood**<br>**and**<br>**Caregiver**<br>**Loss** | 0.46 (0.46,<br>0.46) | 0.51 (0.51,<br>0.51) | +10.6%<br>(+10.2%,<br>+11.1%) | 0.62 (0.62,<br>0.63) | 0.72 (0.72,<br>0.72) | +40.8%<br>(+40.1%,<br>+41.3%) | +55.6%<br>(+54.9%,<br>+56.3%) |
| **Prevalence (n, (95% uncertainty interval))** |  |  |  |  |  |  |  |  |
|  | **Orphanhood** | 2,228,940<br>(2,225,912,<br>2,232,100) | 2,174,707<br>(2,172,340,<br>2,177,566) | -2.4 (-2.6, -<br>2.3) | 2,266,808<br>(2,264,259,<br>2,269,533) | 2,394,604<br>(2,391,948,<br>2,397,308) | +10.1%<br>(+10.0%,<br>+10.2%) | +7.4%<br>(+7.2%,<br>+7.6%) |
|  | **Orphanhood**<br>**and**<br>**Caregiver**<br>**Loss** | 2,738,807<br>(2,735,607,<br>2,742,282) | 2,712,927<br>(2,710,251,<br>2,715,769) | -0.9 (-1.1, -<br>0.8) | 2,824,422<br>(2,821,579,<br>2,827,337) | 2,966,008<br>(2,963,030,<br>2,968,931) | +9.3%<br>(+9.3%,<br>+9.4%) | +8.3%<br>(+8.1%,<br>+8.5%) |
| **Prevalence rate per 100 children (rate, (95% uncertainty interval))** |  |  |  |  |  |  |  |  |
|  | **Orphanhood** | 3.08 (3.08,<br>3.08) | 2.98 (2.97,<br>2.98) | -3.4 (-3.6, -<br>3.2) | 3.11 (3.11,<br>3.12) | 3.45 (3.45,<br>3.45) | +16.0%<br>(+15.9%,<br>+16.1%) | +12.0%<br>(+11.8%,<br>+12.3%) |
|  | **Orphanhood**<br>**and**<br>**Caregiver**<br>**Loss** | 3.78 (3.78,<br>3.79) | 3.71 (3.71,<br>3.72) | -1.9 (-2.1, -<br>1.8) | 3.88 (3.87,<br>3.88) | 4.27 (4.27,<br>4.28) | +15.1%<br>(+15.1%,<br>+15.2%) | +12.9%<br>(+12.8%,<br>+13.1%) |

### Cause-specific trends

From 2000-2021, orphanhood caused by drug overdose increased markedly, as did orphanhood due to COVID-19 between 2019-2021. Orphanhood due to suicide decreased until 2010 and subsequently increased. Orphanhood due to unintentional injuries and homicide decreased until 2019, then increased until 2021; orphanhood due to diseases of the heart remained relatively unchanged until 2019, then increased. Only orphanhood due to malignant neoplasms consistently decreased from 2000-2021. By 2021, both incidence and prevalence of orphanhood due to fatal injuries – including drug overdose, suicide, homicide, and unintentional injuries (e.g. motor vehicle crashes) – exceeded orphanhood due to chronic diseases (Fig 1A, B and Table S1). Orphanhood caused by drug overdose rose substantially from 2012-2019, then jumped sharply during 2020-2021 (Fig 1C). From 2000-2019, orphanhood incidence due to drug overdose increased from an estimated 0.015% of children in 2000, to 0.06% in 2019, 0.085% in 2020 after the pandemic onset, and 0.09% in 2021. Drug overdose was the leading cause of orphanhood incidence and prevalence in 2020 and 2021, surpassing both chronic diseases and COVID-19 (Fig 1C, D). The disproportionate impact of fatal injuries on orphanhood was evident in 2021, with drug overdose contributing 17.5% to orphanhood incidence, yet only 3.1% to adult mortality (Fig 1E); similarly, unintentional injuries, suicide, and homicide contributed 8.3%, 5.0%, and 3.7% to orphanhood, versus 3.6%, 1.2% and 0.7%, respectively, to adult deaths.

**Figure 1.**
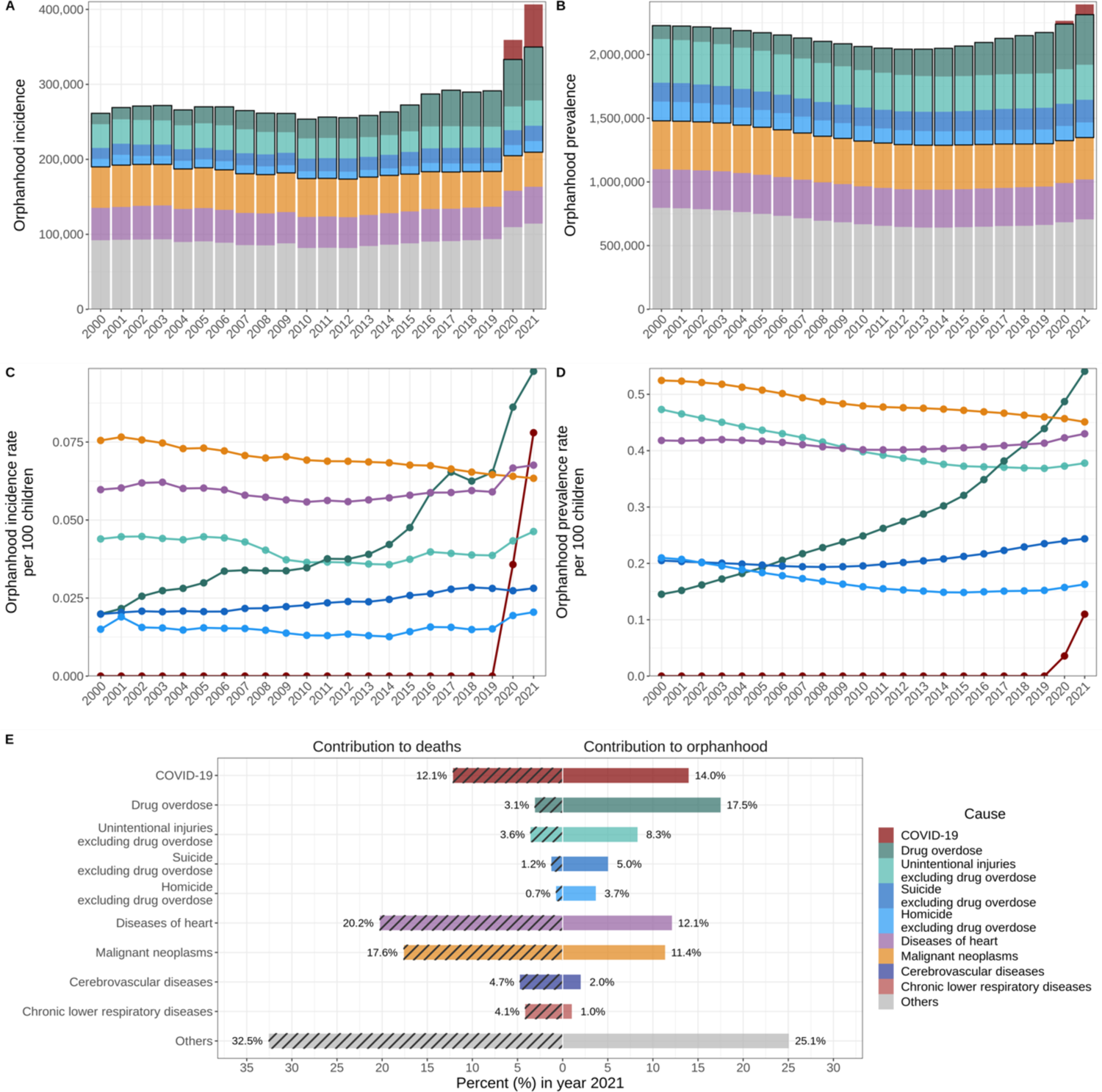
Magnitude of children experiencing orphanhood in the United States. (**A**) Estimated number of U.S. children newly experiencing orphanhood by any cause and over time. (**B**) Estimated number of U.S. children experiencing orphanhood in their lifetime (over the past 18 years) by any cause and over time. (**C**) Incidence rates of orphanhood among US children. (**D**) Prevalence rates of orphanhood among US children. (**E**) Main contributors to children experiencing orphanhood (parental death) versus the main contributors to adult deaths in 2021.

### Disparities by age and race/ethnicity

From 2000-2021, children ages 10-17 years were 5.4 times more likely than children ages 0-4 to experience prevalent orphanhood (Fig 2A, Table S2). By 2021, orphanhood prevalence among ages 10-17 was 5.0%, affecting 1,704,671 children. Substantial disparities across race and ethnicity occurred, accentuated by the COVID-19 pandemic. By 2021, orphanhood affected 6.5% of non-Hispanic American Indian or Alaska Native children, 4.8% of non-Hispanic Black children, 3.9% of non-Hispanic White children, 1.7% of Hispanic children, and 1.9% of non-Hispanic Asian children (Fig 2C).

**Figure 2.**
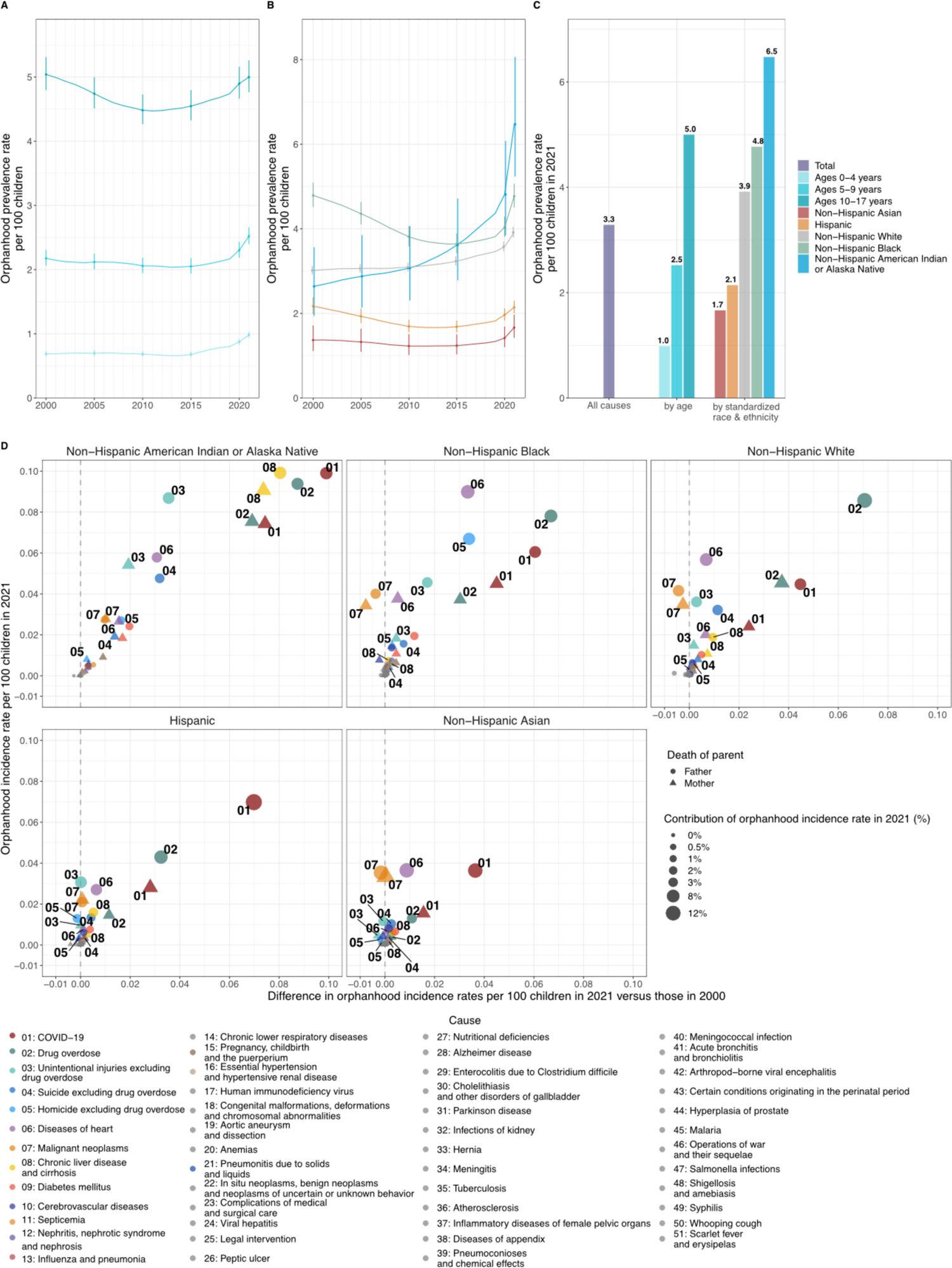
Time trends in orphanhood in the United States by age of child, standardized race & ethnicity, and cause. (**A**) Prevalence rates of orphanhood among US children aged 0-4 years, 5-9 years, and 10-17 years. (**B**) Prevalence rates of orphanhood among children by race and ethnicity of the deceased parent. (**C**) Prevalence rate of all-cause orphanhood by age and standardized race & ethnicity in 2021. (**D**) Time trends in sex and cause-specific incidence rates of orphanhood according to race among US children. On the y-axis are shown 2021 incidence rates of orphanhood among US children, by sex and cause of death of the deceased parent. On the x-axis are shown the differences in incidence rates in 2021 minus those in 2000, with positive differences indicating increases in incidence rates and negative differences indicating decreases in incidence rates. Numbers show the cause of orphanhood. Shapes show the sex of the deceased parent. The size of points indicates the contribution of each cause to new cases of orphanhood respectively among US children in 2021.

Causes underpinning these disparities differed by parental race/ethnicity and sex (Fig 2D, Table S3). Among non-Hispanic American Indian or Alaska Native children, leading causes of maternal orphanhood in 2021 were, in order, COVID-19, drug overdose, and chronic liver disease and cirrhosis; while leading causes of paternal orphanhood were chronic liver disease and cirrhosis, unintentional injuries, and diseases of the heart – all of which increased as causes of orphanhood since 2000. Among non-Hispanic Black children, leading causes of maternal orphanhood were COVID-19, diseases of heart, and drug overdose; while paternal orphanhood was primariy caused by diseases of heart, drug overdose, and homicide. In non-Hispanic White children, drug overdose deaths among fathers increased substantially over the past two decades; in Hispanic children, COVID-19 deaths among fathers predominated. In non-Hispanic Asian children, primary causes of orphanhood were heart disease, malignant neoplasms, and COVID-19.

### State-level Estimates

In 2021, 33 states had >3% of all children experiencing prevalent orphanhood; Southeastern, Northeastern, Southern Border, and Midwest states were most affected (Table 2, Fig. 3, and Tables S4-S6). California, Texas, and Florida had the highest numbers of children experiencing orphanhood incidence and prevalence in 2021. West Virginia and New Mexico had the highest incidence (0.8-0.9%) and prevalence rates (4.5-5.0%) of orphanhood in 2021. Injury-associated parental deaths – including overdose, suicide, and/or unintentional injury – were among the top two causes of orphanhood prevalence in 47 states (Table 2). Drug overdose was the leading cause of orphanhood prevalence in 30 states, particularly affecting Southeastern, Southwestern, Southern Border, and Midwest states, and Southwestern States with large American Indian and Alaska Native populations.

**Figure 3.**
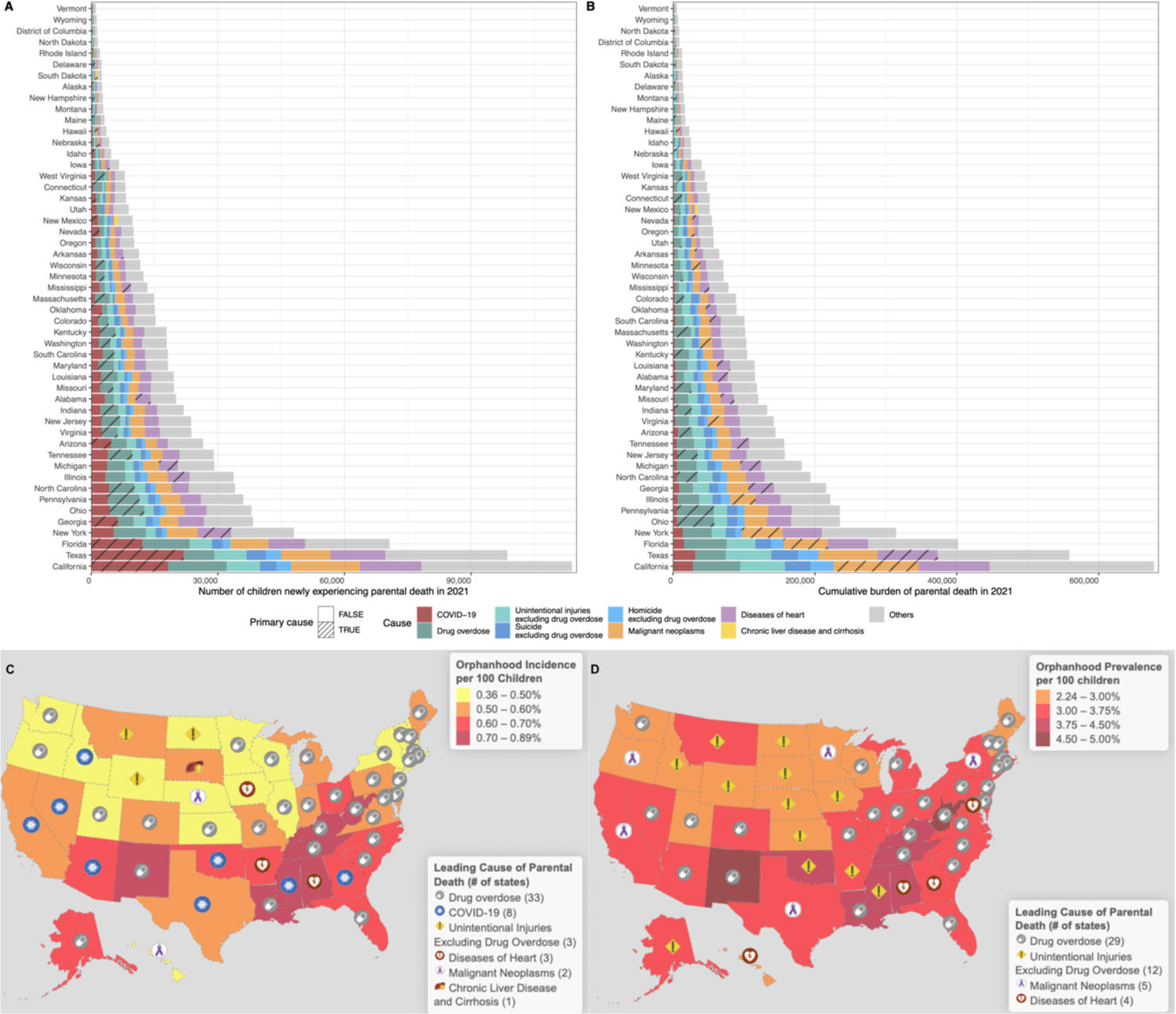
Spatial distribution of US children experiencing orphanhood in 2021. (**A**) Estimated number of U.S. children newly experiencing parental death as the sum of orphanhood, by state and cause. (**B**) Estimated number of U.S. children experiencing parental death in their lifetime (over the past 18 years), by state and cause. Throughout (A) and (B), the leading cause of orphanhood in each state is indicated with hatch marks. (**C**) Map of orphanhood incidence rates per 100 children, by state and leading cause. (**D**) Map of orphanhood prevalence rate per 100 children, by state and leading cause.

**Table 2.** Primary contributors to orphanhood in each US state in 2021.

|  | Orphanhood in 2021 |  |  |  |  |  |  |  |
| --- | --- | --- | --- | --- | --- | --- | --- | --- |
|  | Orphanhood Incidence in 2021 |  |  |  | Orphanhood Prevalence in 2021 |  |  |  |
| U.S. state | Incidence<br>(n, 95% UI) | Incidence<br>rate<br>per 100<br>children<br>(rate, 95%<br>UI) | First ranked<br>cause<br>(name,<br>% contribution) | Second ranked<br>cause<br>(name,<br>% contribution) | Prevalence<br>(n, 95% UI) | Prevalence<br>rate<br>per 100<br>children<br>(rate, 95%<br>UI) | First ranked<br>cause<br>(name,<br>% contribution) | Second ranked<br>cause<br>(name,<br>% contribution) |
| Alabama | 7,991<br>(7,585,<br>8,397) | 0.71 (0.68,<br>0.75) | Diseases of<br>heart<br>(16.4%) | COVID-19<br>(16.2%) | 45,696<br>(42,504,<br>49,079) | 4.07 (3.79,<br>4.37) | Diseases of<br>heart<br>(16.9%) | Unintentional<br>injuries<br>excluding drug<br>overdose<br>(14.0%) |
| Alaska | 1,122 (795,<br>1,478) | 0.63 (0.44,<br>0.82) | Drug overdose<br>(16.0%) | Unintentional<br>injuries<br>excluding drug<br>overdose<br>(15.2%) | 5,922<br>(3,970,<br>9,170) | 3.30 (2.21,<br>5.11) | Unintentional<br>injuries<br>excluding drug<br>overdose<br>(23.8%) | Suicide<br>excluding drug<br>overdose<br>(10.7%) |
| Arizona | 10,898<br>(10,511,<br>11,250) | 0.68 (0.65,<br>0.70) | COVID-19<br>(17.8%) | Drug overdose<br>(16.5%) | 59,002<br>(55,837,<br>62,261) | 3.66 (3.46,<br>3.86) | Drug overdose<br>(16.4%) | Unintentional<br>injuries<br>excluding drug<br>overdose<br>(12.9%) |
| Arkansas | 4,461<br>(4,053,<br>4,945) | 0.63 (0.58,<br>0.70) | Diseases of<br>heart<br>(15.2%) | COVID-19<br>(13.2%) | 25,644<br>(22,348,<br>29,481) | 3.65 (3.18,<br>4.19) | Unintentional<br>injuries<br>excluding drug<br>overdose<br>(16.6%) | Diseases of<br>heart<br>(15.4%) |
| California | 45,004<br>(45,713,<br>44,534) | 0.51 (0.52,<br>0.51) | COVID-19<br>(16.5%) | Drug overdose<br>(14.6%) | 264,622<br>(268,326,<br>261,397) | 3.02 (3.06,<br>2.98) | Malignant<br>neoplasms<br>(16.8%) | Diseases of<br>heart<br>(12.8%) |
| Colorado | 6,587<br>(6,178,<br>6,985) | 0.53 (0.50,<br>0.56) | Drug overdose<br>(18.0%) | COVID-19<br>(10.4%) | 37,868<br>(34,890,<br>41,412) | 3.05 (2.81,<br>3.33) | Drug overdose<br>(16.6%) | Suicide<br>excluding drug<br>overdose<br>(13.5%) |
| Connecticut | 3,330<br>(2,922,<br>3,741) | 0.46 (0.40,<br>0.51) | Drug overdose<br>(29.0%) | Malignant<br>neoplasms<br>(13.3%) | 20,874<br>(17,911,<br>24,220) | 2.86 (2.45,<br>3.32) | Drug overdose<br>(26.5%) | Malignant<br>neoplasms<br>(14.9%) |
| Delaware | 962 (667,<br>1,279) | 0.46 (0.32,<br>0.61) | Drug overdose<br>(34.4%) | Malignant<br>neoplasms<br>(9.7%) | 5,467<br>(3,582,<br>7,985) | 2.62 (1.72,<br>3.83) | Drug overdose<br>(29.4%) | Unintentional<br>injuries<br>excluding drug<br>overdose<br>(10.2%) |
| District of<br>Columbia | 612 (432,<br>823) | 0.49 (0.34,<br>0.65) | Drug overdose<br>(18.3%) | Diseases of<br>heart<br>(10.8%) | 3,828<br>(2,465,<br>5,519) | 3.04 (1.96,<br>4.39) | Diseases of<br>heart<br>(13.0%) | Drug overdose<br>(12.4%) |
| Florida | 28,456<br>(28,600,<br>28,393) | 0.66 (0.67,<br>0.66) | Drug overdose<br>(19.0%) | COVID-19<br>(17.9%) | 159,629<br>(159,761,<br>159,719) | 3.72 (3.72,<br>3.72) | Drug overdose<br>(18.2%) | Malignant<br>neoplasms<br>(14.1%) |
| Georgia | 15,723<br>(15,459,<br>16,039) | 0.62 (0.61,<br>0.64) | COVID-19<br>(16.5%) | Diseases of<br>heart<br>(15.2%) | 87,557<br>(84,749,<br>90,515) | 3.47 (3.36,<br>3.59) | Diseases of<br>heart<br>(15.6%) | Malignant<br>neoplasms<br>(13.0%) |
| Hawaii | 1,231 (950,<br>1,580) | 0.40 (0.31,<br>0.52) | Malignant<br>neoplasms<br>(16.2%) | Diseases of<br>heart<br>(15.4%) | 7,692<br>(5,510,<br>10,706) | 2.53 (1.81,<br>3.52) | Diseases of<br>heart<br>(15.3%) | Malignant<br>neoplasms<br>(15.3%) |
| Idaho | 1,940<br>(1,544,<br>2,372) | 0.41 (0.33,<br>0.51) | COVID-19<br>(14.5%) | Drug overdose<br>(13.0%) | 10,474<br>(7,785,<br>14,079) | 2.23 (1.66,<br>3.00) | Unintentional<br>injuries<br>excluding drug<br>overdose<br>(20.3%) | Suicide<br>excluding drug<br>overdose<br>(13.0%) |
| Illinois | 13,831<br>(13,528,<br>14,158) | 0.49 (0.48,<br>0.51) | Drug overdose<br>(16.4%) | Diseases of<br>heart<br>(13.9%) | 89,775<br>(87,129,<br>92,407) | 3.20 (3.11,<br>3.30) | Drug overdose<br>(16.5%) | Malignant<br>neoplasms<br>(15.1%) |
| Indiana | 9,148<br>(8,705,<br>9,532) | 0.58 (0.55,<br>0.60) | Drug overdose<br>(22.6%) | Diseases of<br>heart<br>(11.9%) | 54,889<br>(51,659,<br>58,427) | 3.46 (3.26,<br>3.68) | Drug overdose<br>(20.2%) | Diseases of<br>heart<br>(13.0%) |
| Iowa | 2,784<br>(2,381,<br>3,218) | 0.38 (0.32,<br>0.44) | Diseases of<br>heart<br>(14.8%) | Malignant<br>neoplasms<br>(12.5%) | 16,673<br>(13,710,<br>20,323) | 2.26 (1.86,<br>2.76) | Unintentional<br>injuries<br>excluding drug<br>overdose<br>(15.7%) | Malignant<br>neoplasms<br>(14.5%) |
| Kansas | 3,455<br>(3,041,<br>3,947) | 0.49 (0.43,<br>0.56) | Drug overdose<br>(14.4%) | COVID-19<br>(13.1%) | 19,915<br>(16,740,<br>23,836) | 2.83 (2.38,<br>3.39) | Unintentional<br>injuries<br>excluding drug<br>overdose<br>(15.8%) | Malignant<br>neoplasms<br>(12.5%) |
| Kentucky | 7,183<br>(6,792,<br>7,634) | 0.71 (0.67,<br>0.75) | Drug overdose<br>(24.3%) | Diseases of<br>heart<br>(13.9%) | 42,002<br>(38,999,<br>45,578) | 4.13 (3.84,<br>4.49) | Drug overdose<br>(22.9%) | Diseases of<br>heart<br>(14.0%) |
| Louisiana | 8,108<br>(7,719,<br>8,511) | 0.75 (0.71,<br>0.79) | Drug overdose<br>(23.3%) | Diseases of<br>heart<br>(12.7%) | 46,778<br>(43,522,<br>50,175) | 4.32 (4.02,<br>4.63) | Drug overdose<br>(17.3%) | Diseases of<br>heart<br>(14.2%) |
| Maine | 1,347<br>(1,056,<br>1,679) | 0.53 (0.42,<br>0.67) | Drug overdose<br>(30.1%) | Diseases of<br>heart<br>(10.8%) | 7,245<br>(5,099,<br>9,975) | 2.88 (2.02,<br>3.96) | Drug overdose<br>(26.8%) | Unintentional<br>injuries<br>excluding drug<br>overdose<br>(13.6%) |
| Maryland | 7,419<br>(6,996,<br>7,815) | 0.54 (0.51,<br>0.57) | Drug overdose<br>(23.5%) | Diseases of<br>heart<br>(13.8%) | 48,147<br>(44,993,<br>51,637) | 3.53 (3.30,<br>3.79) | Drug overdose<br>(22.7%) | Diseases of<br>heart<br>(14.6%) |
| Massachusetts | 6,038<br>(5,612,<br>6,457) | 0.44 (0.41,<br>0.47) | Drug overdose<br>(27.6%) | Malignant<br>neoplasms<br>(14.7%) | 41,234<br>(38,098,<br>44,636) | 3.03 (2.80,<br>3.28) | Drug overdose<br>(27.4%) | Malignant<br>neoplasms<br>(15.8%) |
| Michigan | 12,179<br>(11,820,<br>12,509) | 0.57 (0.55,<br>0.58) | Drug overdose<br>(17.0%) | Diseases of<br>heart<br>(13.6%) | 75,255<br>(72,209,<br>78,059) | 3.49 (3.35,<br>3.62) | Drug overdose<br>(17.9%) | Diseases of<br>heart<br>(14.4%) |
| Minnesota | 5,403<br>(4,951,<br>5,883) | 0.41 (0.38,<br>0.45) | Drug overdose<br>(17.6%) | Malignant<br>neoplasms<br>(13.8%) | 30,442<br>(27,178,<br>34,423) | 2.31 (2.06,<br>2.61) | Malignant<br>neoplasms<br>(16.8%) | Drug overdose<br>(14.6%) |
| Mississippi | 5,251<br>(4,791,<br>5,668) | 0.76 (0.69,<br>0.82) | COVID-19<br>(15.0%) | Diseases of<br>heart<br>(14.7%) | 30,280<br>(26,942,<br>33,913) | 4.37 (3.89,<br>4.89) | Unintentional<br>injuries<br>excluding drug<br>overdose<br>(16.7%) | Diseases of<br>heart<br>(15.5%) |
| Missouri | 8,128<br>(7,727,<br>8,525) | 0.59 (0.56,<br>0.62) | Drug overdose<br>(18.3%) | Diseases of<br>heart<br>(13.5%) | 49,665<br>(46,695,<br>52,986) | 3.59 (3.37,<br>3.83) | Drug overdose<br>(18.1%) | Diseases of<br>heart<br>(13.9%) |
| Montana | 1,216 (912,<br>1,568) | 0.52 (0.39,<br>0.67) | Unintentional<br>injuries<br>excluding drug<br>overdose<br>(17.3%) | Suicide<br>excluding drug<br>overdose<br>(10.7%) | 6,553<br>(4,473,<br>9,605) | 2.79 (1.90,<br>4.09) | Unintentional<br>injuries<br>excluding drug<br>overdose<br>(24.6%) | Suicide<br>excluding drug<br>overdose<br>(12.7%) |
| Nebraska | 1,809<br>(1,412,<br>2,261) | 0.37 (0.29,<br>0.47) | Malignant<br>neoplasms<br>(13.7%) | Unintentional<br>injuries<br>excluding drug<br>overdose<br>(12.1%) | 10,680<br>(8,043,<br>14,384) | 2.21 (1.67,<br>2.98) | Unintentional<br>injuries<br>excluding drug<br>overdose<br>(18.6%) | Malignant<br>neoplasms<br>(14.3%) |
| Nevada | 4,034<br>(3,650,<br>4,450) | 0.58 (0.52,<br>0.64) | COVID-19<br>(19.3%) | Diseases of<br>heart<br>(13.6%) | 21,946<br>(18,876,<br>25,409) | 3.14 (2.70,<br>3.64) | Drug overdose<br>(15.5%) | Diseases of<br>heart<br>(14.8%) |
| New<br>Hampshire | 1,094 (814,<br>1,405) | 0.43 (0.32,<br>0.55) | Drug overdose<br>(25.4%) | Malignant<br>neoplasms<br>(10.9%) | 6,869<br>(4,778,<br>9,552) | 2.68 (1.86,<br>3.73) | Drug overdose<br>(30.8%) | Malignant<br>neoplasms<br>(11.4%) |
| New Jersey | 9,644<br>(9,235,<br>10,114) | 0.48 (0.46,<br>0.50) | Drug overdose<br>(21.9%) | Malignant<br>neoplasms<br>(13.7%) | 63,595<br>(60,303,<br>67,269) | 3.14 (2.98,<br>3.32) | Drug overdose<br>(21.5%) | Malignant<br>neoplasms<br>(15.3%) |
| New Mexico | 4,137<br>(3,701,<br>4,558) | 0.87 (0.78,<br>0.96) | Drug overdose<br>(16.7%) | COVID-19<br>(16.2%) | 21,632<br>(18,554,<br>25,154) | 4.57 (3.92,<br>5.32) | Drug overdose<br>(17.6%) | Unintentional<br>injuries<br>excluding drug<br>overdose<br>(15.1%) |
| New York | 19,199<br>(19,000,<br>19,436) | 0.47 (0.46,<br>0.47) | Drug overdose<br>(19.2%) | Malignant<br>neoplasms<br>(14.8%) | 124,546<br>(122,607,<br>126,477) | 3.03 (2.98,<br>3.07) | Malignant<br>neoplasms<br>(17.8%) | Drug overdose<br>(15.9%) |
| North Carolina | 14,097<br>(13,786,<br>14,457) | 0.61 (0.60,<br>0.63) | Drug overdose<br>(20.5%) | COVID-19<br>(12.4%) | 79,120<br>(76,394,<br>82,029) | 3.44 (3.32,<br>3.56) | Drug overdose<br>(17.7%) | Malignant<br>neoplasms<br>(12.6%) |
| North Dakota | 695 (469,<br>1,081) | 0.37 (0.25,<br>0.58) | Unintentional<br>injuries<br>excluding drug<br>overdose<br>(15.0%) | Drug overdose<br>(10.2%) | 3,808<br>(2,409,<br>6,887) | 2.05 (1.30,<br>3.71) | Unintentional<br>injuries<br>excluding drug<br>overdose<br>(17.7%) | Suicide<br>excluding drug<br>overdose<br>(7.9%) |
| Ohio | 15,767<br>(15,525,<br>16,051) | 0.61 (0.60,<br>0.62) | Drug overdose<br>(24.9%) | Diseases of<br>heart<br>(12.2%) | 97,161<br>(94,914,<br>99,413) | 3.73 (3.64,<br>3.82) | Drug overdose<br>(26.0%) | Diseases of<br>heart<br>(12.9%) |
| Oklahoma | 6,119<br>(5,684,<br>6,576) | 0.64 (0.59,<br>0.68) | COVID-19<br>(16.9%) | Diseases of<br>heart<br>(14.3%) | 36,240<br>(32,901,<br>39,983) | 3.77 (3.42,<br>4.16) | Unintentional<br>injuries<br>excluding drug<br>overdose<br>(14.8%) | Diseases of<br>heart<br>(14.7%) |
| Oregon | 4,153<br>(3,757,<br>4,561) | 0.48 (0.44,<br>0.53) | Drug overdose<br>(15.7%) | Malignant<br>neoplasms<br>(13.3%) | 22,994<br>(19,901,<br>26,282) | 2.67 (2.31,<br>3.05) | Malignant<br>neoplasms<br>(14.9%) | Unintentional<br>injuries<br>excluding drug<br>overdose<br>(13.6%) |
| Pennsylvania | 14,609<br>(14,349,<br>14,918) | 0.55 (0.54,<br>0.56) | Drug overdose<br>(24.7%) | Malignant<br>neoplasms<br>(12.0%) | 95,425<br>(93,303,<br>97,704) | 3.57 (3.49,<br>3.65) | Drug overdose<br>(26.3%) | Malignant<br>neoplasms<br>(13.2%) |
| Rhode Island | 791 (565,<br>1,048) | 0.38 (0.27,<br>0.50) | Drug overdose<br>(35.7%) | Diseases of<br>heart<br>(9.6%) | 4,826<br>(3,084,<br>7,091) | 2.31 (1.48,<br>3.40) | Drug overdose<br>(30.3%) | Malignant<br>neoplasms<br>(11.3%) |
| South Carolina | 7,274<br>(6,907,<br>7,638) | 0.65 (0.62,<br>0.68) | Drug overdose<br>(18.1%) | COVID-19<br>(14.6%) | 39,654<br>(36,661,<br>42,890) | 3.55 (3.28,<br>3.84) | Drug overdose<br>(14.8%) | Unintentional<br>injuries<br>excluding drug<br>overdose<br>(14.7%) |
| South Dakota | 1,093 (794,<br>1,510) | 0.50 (0.36,<br>0.69) | Chronic liver<br>disease and<br>cirrhosis<br>(17.7%) | Unintentional<br>injuries<br>excluding drug<br>overdose<br>(11.7%) | 5,552<br>(3,631,<br>9,040) | 2.52 (1.65,<br>4.10) | Unintentional<br>injuries<br>excluding drug<br>overdose<br>(19.5%) | Suicide<br>excluding drug<br>overdose<br>(8.6%) |
| Tennessee | 11,908<br>(11,590,<br>12,260) | 0.77 (0.75,<br>0.80) | Drug overdose<br>(23.3%) | COVID-19<br>(13.5%) | 63,260<br>(60,342,<br>66,515) | 4.11 (3.92,<br>4.32) | Drug overdose<br>(18.8%) | Diseases of<br>heart<br>(14.5%) |
| Texas | 40,272<br>(40,738,<br>39,926) | 0.54 (0.54,<br>0.53) | COVID-19<br>(23.2%) | Diseases of<br>heart<br>(12.0%) | 224,453<br>(226,165,<br>223,024) | 3.00 (3.03,<br>2.98) | Malignant<br>neoplasms<br>(13.9%) | Diseases of<br>heart<br>(13.7%) |
| Utah | 3,772<br>(3,283,<br>4,293) | 0.40 (0.35,<br>0.45) | Drug overdose<br>(14.5%) | COVID-19<br>(14.2%) | 24,529<br>(20,856,<br>29,053) | 2.59 (2.20,<br>3.07) | Drug overdose<br>(19.9%) | Suicide<br>excluding drug<br>overdose<br>(15.6%) |
| Vermont | 479 (308,<br>678) | 0.41 (0.26,<br>0.58) | Drug overdose<br>(32.8%) | Unintentional<br>injuries<br>excluding drug<br>overdose<br>(10.4%) | 2,394<br>(1,244,<br>4,255) | 2.05 (1.06,<br>3.64) | Drug overdose<br>(21.0%) | Unintentional<br>injuries<br>excluding drug<br>overdose<br>(11.3%) |
| Virginia | 9,638<br>(9,245,<br>10,045) | 0.51 (0.49,<br>0.53) | Drug overdose<br>(18.6%) | Diseases of<br>heart<br>(13.9%) | 56,961<br>(53,825,<br>60,501) | 3.02 (2.86,<br>3.21) | Drug overdose<br>(16.2%) | Malignant<br>neoplasms<br>(15.3%) |
| Washington | 7,386<br>(6,998,<br>7,814) | 0.44 (0.42,<br>0.47) | Drug overdose<br>(17.8%) | Malignant<br>neoplasms<br>(13.6%) | 42,455<br>(39,491,<br>46,039) | 2.53 (2.36,<br>2.75) | Drug overdose<br>(15.9%) | Malignant<br>neoplasms<br>(15.6%) |
| West Virginia | 3,142<br>(2,754,<br>3,552) | 0.88 (0.77,<br>0.99) | Drug overdose<br>(35.2%) | COVID-19<br>(10.6%) | 17,888<br>(15,069,<br>21,094) | 4.98 (4.20,<br>5.88) | Drug overdose<br>(32.8%) | Unintentional<br>injuries<br>excluding drug<br>overdose<br>(11.2%) |
| Wisconsin | 4,913<br>(4,528,<br>5,358) | 0.39 (0.36,<br>0.42) | Drug overdose<br>(21.6%) | Malignant<br>neoplasms<br>(12.1%) | 29,802<br>(26,821,<br>33,307) | 2.34 (2.10,<br>2.61) | Drug overdose<br>(19.9%) | Malignant<br>neoplasms<br>(14.7%) |
| Wyoming | 542 (348, 880) | 0.41 (0.26, 0.66) | Unintentional injuries excluding drug overdose (13.5%) | Suicide excluding drug overdose (10.3%) | 3,021 (1,734, 5,830) | 2.28 (1.31, 4.40) | Unintentional injuries excluding drug overdose (25.4%) | Suicide excluding drug overdose (11.2%) |

UI = Uncertainty interval.

## Discussion

Over the past two decades, prevalence of all-cause orphanhood and caregiver death decreased slightly until 2019 and then increased markedly during 2020-2021 with intersecting crises of the overdose epidemic and COVID-19 pandemic.^36,37^ Parental death due to drug overdose was the leading cause of orphanhood incidence and prevalence during the pandemic. By 2021, over 2.9 million children – 4.0% of all children in the U.S. – had lost a parent or caregiver in their lifetime. Populations disproportionately affected by orphanhood included over 1.7 million adolescents ages 10-17 years (1 of every 20 adolescents); and children of non-Hispanic American Indian or Alaska Native, non-Hispanic Black, and non-Hispanic White race/ethnicities (approximately 1 of 15, 1 of 20, and 1 of 25 children, respectively). Orphanhood prevalence was highest in West Virginia, New Mexico, Mississippi, Louisiana, and Kentucky, affecting 1 of every 25 children. Compared to previous cause-specific reports (an estimated 97,376 children orphaned by HIV/AIDS (1998),^27^ 157,183 by maternal cancers (2020),^23^ and 218,800 by COVID-19 in 2020-2022^38^), data on all-cause orphanhood and co-residing grandparent caregiver death are over ten times greater, quantifying the full burden.

By 2021, orphanhood incidence and prevalence due to fatal injuries (drug overdose, suicide, homicide, and unintentional injuries), exceeded those linked to chronic diseases. Together, injuries and chronic diseases accounted for over half of all children orphaned. Fatalities due to injuries and chronic diseases among minoritized populations, are more common among younger adults still caring for children. For 47 states, drug overdose, suicide, and/or unintentional injuries were among the top two causes of orphanhood. Although drug overdose deaths were increasing before 2020, pandemic-linked increases of these deaths exceeded those forecasted; and the pandemic appeared to increase drug overdose risk.^39,40^ The evolving nature of drug supply, including the proliferation of illegally made fentanyl and resurgence of stimulants such as methamphetamine, has affected almost every state.^41,42^ By 2021, our data show drug overdose contributed 17.5% to orphanhood compared to 3.1% to mortality; establishing comprehensive care programs for children orphaned due to drug overdose could better mitigate the long-term negative effects that these children are disproportionately facing.

Our findings reveal social and health disparities in orphanhood rates and leading caregiver causes-of-death between age, sex, and racial/ethnic groups, and across geographies. These results can inform and contextualize prevention and response programming for children affected by orphanhood. For non-Hispanic American Indian or Alaska Native persons, for example, alcohol-related deaths were a leading cause, while non-Hispanic Black populations showed homicide among fathers as a leading cause. Many of these disparities are related to historical racism and derived practices that foster inequitable access to housing, education, and employment. Populations most affected in different states may have been exposed to such inequalities.^43^ In parts of the northeastern and midwestern U.S., rising unemployment, limited access to health and social services during the pandemic, increased access to a more lethal supply of illicit drugs or availability of other substances such as alcohol particularly among affected white men, may be linked to drug overdose emerging as the top cause of incident orphanhood in these regions.^44^ Circumstances of parental death influence grief-related psychopathology in surviving children,^45^ and evidence-based responses can improve short- and long-term outcomes.^9,11,12^ It is paramount to keep children in their families, whenever possible. This requires ensuring bereaved families receive support, and those needing kinship or foster care are served rapidly.^46^ Child resilience after parental loss can be strengthened by programs and policies that promote safe, stable, and nurturing relationships and environments, and that address childhood adversity, including preventing violence and abuse.^46^

Despite serious risks for bereaved children,^47^ timely, multifactored responses using an age- and life-stage approach may restore hope and build resilience.^48^ As grief is an immediate reaction that can extend years after parental loss, healing and support are urgent needs for children, who may be less equipped to recover than adults, particularly when the loss is sudden and unexpected.^11,27^ The governor of Utah^49^ and others^50^ have called for the addition to the death certificate of a checkbox to identify children living in the home of the deceased, so that bereaved children can be linked to support and services. Such responses typically provide parenting, economic, and education support, and are consistent with the WHO INSPIRE package for ending violence against children.^51,52^

Our study has several limitations. First, cause-specific estimates of children experiencing orphanhood and caregiver deaths are derived from mortality statistics on underlying causes-of-death, and may be underestimates^53^ for causes associated with erroneous or incomplete reporting, uncertainty in the chain of events preceding death, or coding limitations - such as for COVID-19, drug overdose, or suicide.^54^ Second, as we attributed only one child per grandparent caregiver death, our estimates of grandparent caregiver loss are minimum estimates. Third, we assumed current mortality is unrelated to historic fertility for years before 1990, and sensitivity analyses suggest this may lead to inaccuracies in orphanhood prevalence estimates until 2007 (ST S2). Fourth, publicly available state-specific data were partly suppressed due to small counts; therefore, we cannot exclude bias in state-specific orphanhood estimates. Finally, to characterise orphanhood prevalence in 2000-2021, we had to consider vital statistics since 1983 and consolidate changes in cause-of-death and race/ethnicity coding. Our sensitivity analyses, including detailed comparisons of NCHS death data to CDC WONDER vital statistics for each of the 21 years’ calculations (ST S2), suggest our incidence and prevalence estimates are robust, and should be interpreted as minimum estimates of orphanhood and caregiver death.

In conclusion, evidence-based policies and programs that provide healing and support for nearly 3 million children and adolescents in the U.S. who have experienced orphanhood and caregiver loss are urgently needed to reduce long term negative affects of this adverse childhood experience. This is especially relevant given unprecedented rates of drug overdoses, increasing the possibility that children who are living in a household where a parent or caregiver is negatively affected by substance use, may also experience the loss of a parent.^52,55^ Evidence highlights three essential components of orphanhood prevention and response that effectively promote their recovery and resilience: (1) prevent death of parents and caregivers through timely prevention and treatment of leading causes of death and ensured access to affordable compassionate health and mental health care for all; (2) prepare families to provide safe and nurturing alternative care; and (3) protect children affected by orphanhood and vulnerabilities, through grief and mental health counseling, and parenting, economic, and educational support that can be contextualized and delivered at scale.^19,56^

## Ethical approval

All data used for this study are publicly available through U.S. Government websites.

## Data sharing statement

All data used to calculate estimates of mortality and orphanhood are publicly available.

## Code availability

Code to reproduce all analyses is freely available on Github version 1.1.2 under the GNU General Public License version 3.0 at the repository (https://github.com/MLGlobalHealth/orphanhood-caregiver-death-in-US-from-all-causes-of-mortality).

## Data Availability

All data used for this study are publicly available through U.S. Government websites.
All data used to calculate estimates of mortality and orphanhood are publicly available.

https://www.cdc.gov/nchs/data_access/vitalstatsonline.htm

https://wonder.cdc.gov/

## Acknowledgements

We thank the Global Reference Group for Children In Crisis, reviewers at the CDC and NCHS especially Dr. Robert Anderson for his helpful suggestions on interpreting and classifying disease groups and race groups using existing NCHS data. We also thank Prof. Chris Desmond for his comments on early versions of this work. We thank the Imperial College Research Computing Service (https://doi.org/10.14469/hpc/2232) for providing the computational resources to perform this study; and Zulip for sponsoring team communications through the Zulip Cloud Standard chat app. This study was supported by the Oak Foundation (to LC, LS); the Moderna Charitable Foundation (to OR); the World Health Organisation (to SF); the Engineering and Physical Sciences Research Council (EPSRC) through the EPSRC Centre for Doctoral Training in Modern Statistics and Statistical Machine Learning at Imperial College London and Oxford University (EP/S023151/1 to A. Gandy); the Imperial College London President’s PhD Scholarship fund to YC; Imperial College London Undergraduate Research Bursaries to LG and VKM; and London Mathematical Society Undergraduate Research Bursary to DW (URB-2023-86). The funders had no role in study design, data collection and analysis, decision to publish or preparation of the manuscript. The findings and conclusions in this report are those of the author(s) and do not necessarily represent the official position of the Centers for Disease Control and Prevention.

## Copyright statement

For the purpose of open access, the authors have applied a ‘Creative Commons Attribution’ (CC BY) license to any Author Accepted Manuscript version arising.

## Author contributions

AV, SH and OR designed the study; AV, YC, ST, AB, LG, VKM, DW, SH and OR performed the analysis; LG, VKM, DW, JTU and SF checked analysis; AV, LC, LS, FA, GM, JL, LR, RN, CAN, SF, JTU, SH and OR oversaw data interpretation; AV, YC, ST, SH and OR wrote the first draft; All authors discussed the first draft and contributed to the final manuscript.

## Supplementary Materials

### S0 Supplementary Figures and Tables

**Supplementary Fig. S1:**
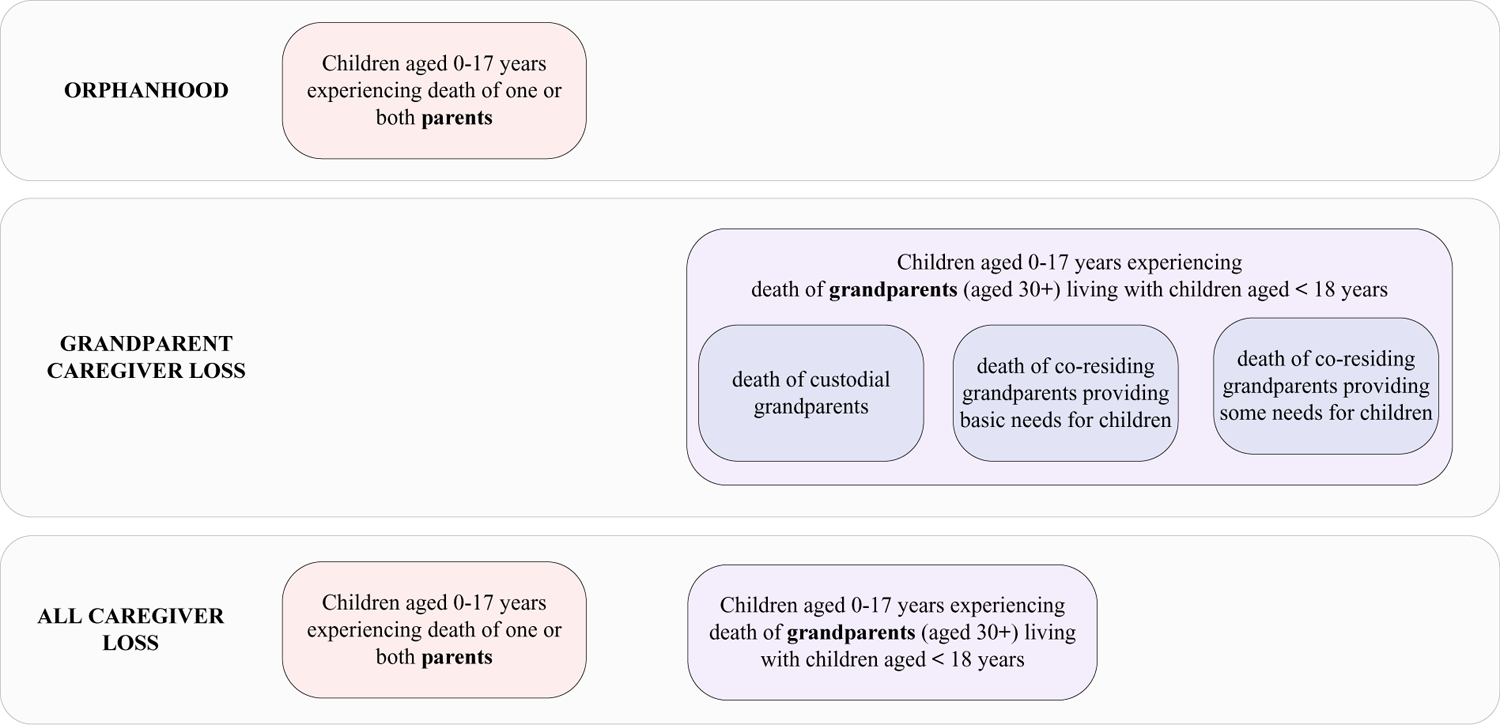
Definitions of orphanhood and grandparent caregiver loss used in this study (orange and purple), with comparison to definitions in previous studies [2021a] (blue).

**Supplementary Fig. S2:**
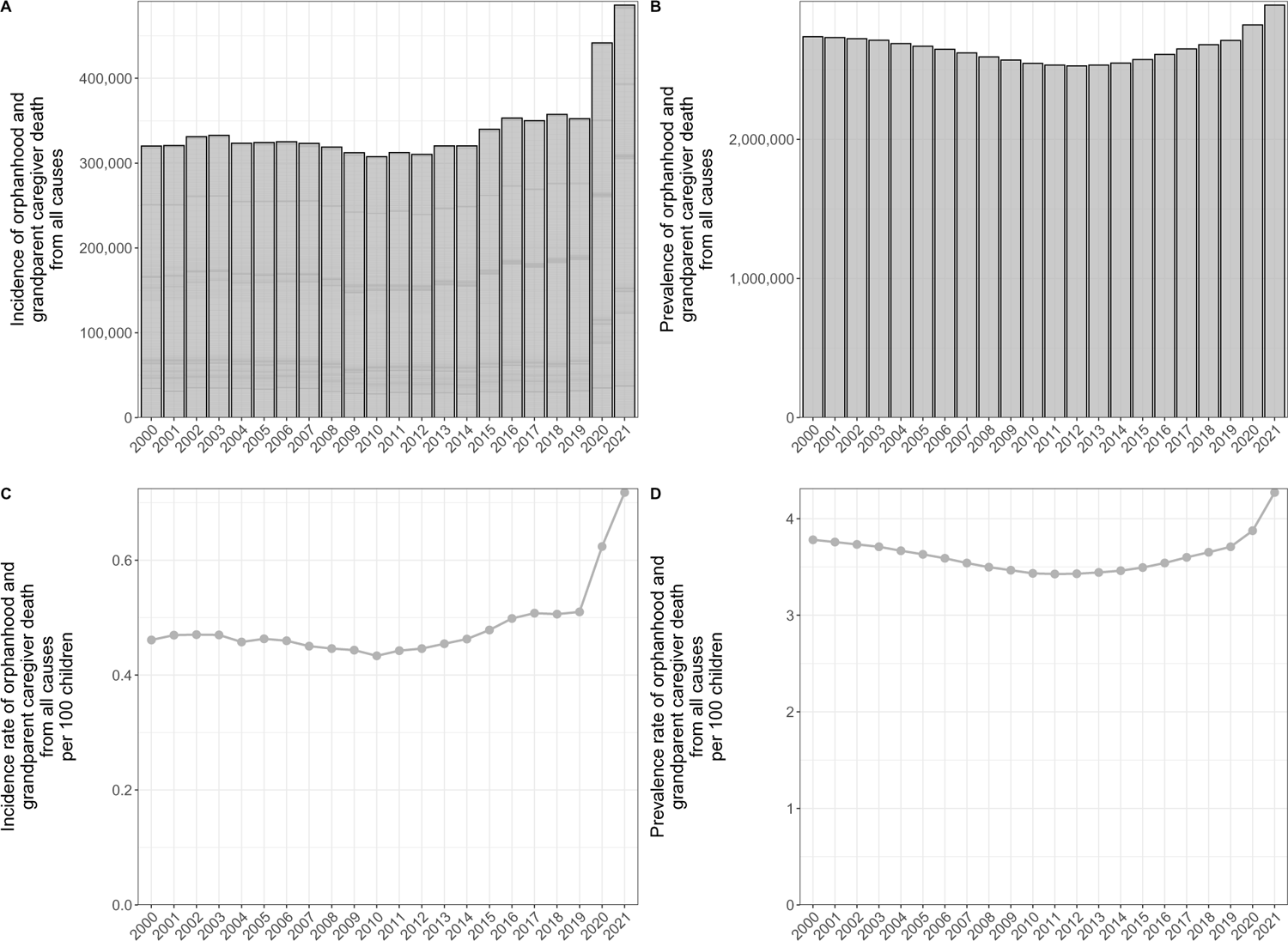
Magnitude of children experiencing the death of a caregiver in the U.S. (A) Estimated number of U.S. children newly experiencing caregiver death as the sum of orphanhood and grandparent caregiver death from all causes (incidence). (B) Estimated number of U.S. children experiencing caregiver death from all causes in their lifetime (prevalence). (C) Incidence rates of caregiver death from all causes among U.S. children. (D) Prevalence rates of caregiver death from all causes among U.S. children.

**Supplementary Fig. S3:**
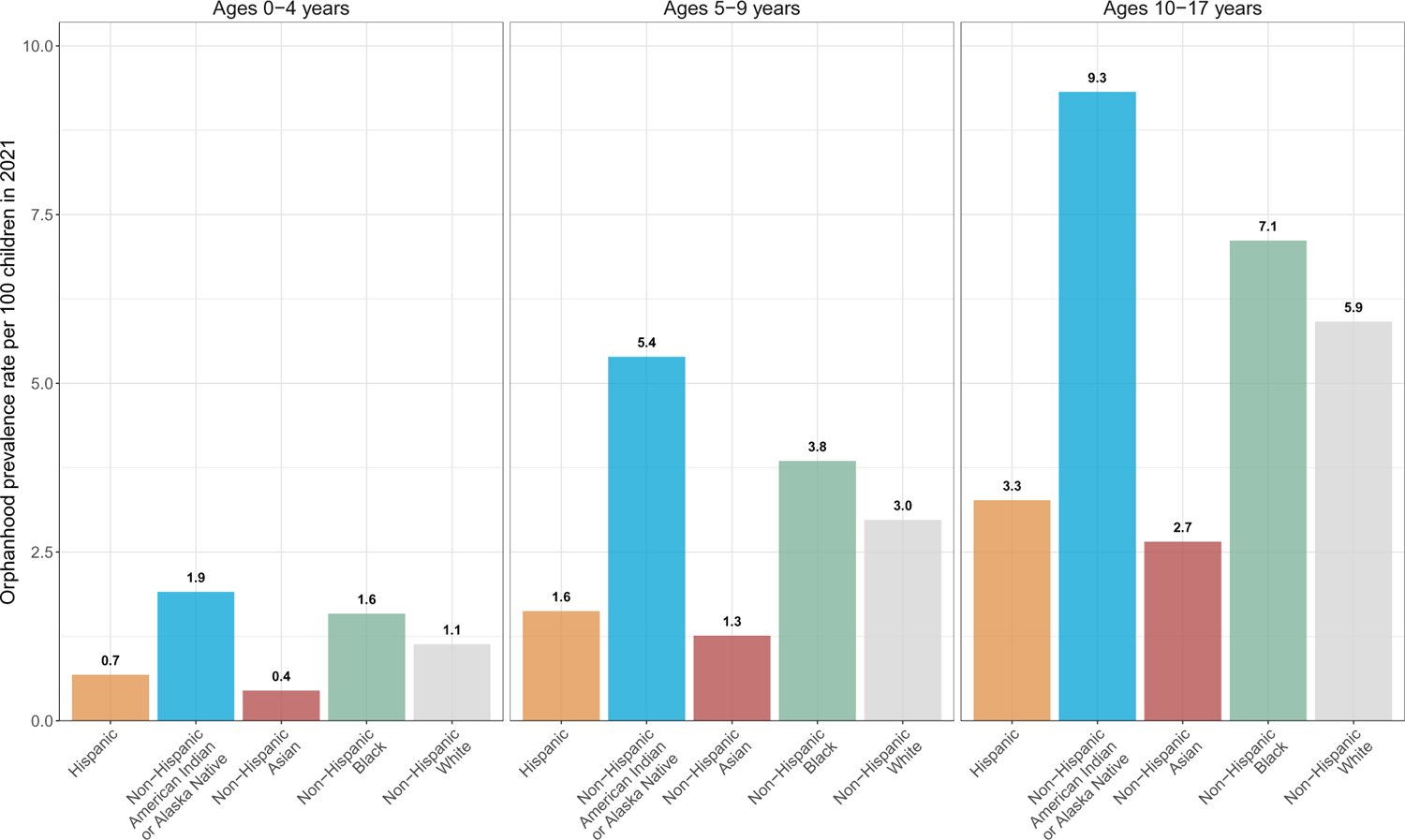
Orphanhood prevalence rates by standardized race & ethnicity in each age group among U.S. children.

**Supplementary Table S1:** Children newly experiencing orphanhood in the U.S. in 2021, by all caregiver loss causes-of-death.

| Caregiver loss<br>cause-of-death | Children newly experiencing<br>orphanhood |  |  | Adult deaths |  |  | Ratio |
| --- | --- | --- | --- | --- | --- | --- | --- |
|  | (rank) | (n) | (proportion) | (rank) | (n) | (proportion) |  |
| Drug overdose | #1 | 71,112 | 17.5% | #8 | 105,143 | 3.1% | 0.68 |
| COVID-19 | #2 | 56,778 | 14.0% | #3 | 414,781 | 12.1% | 0.14 |
| Diseases of heart | #3 | 49,213 | 12.1% | #1 | 692,150 | 20.2% | 0.07 |
| Malignant neoplasms | #4 | 46,145 | 11.4% | #2 | 601,370 | 17.6% | 0.08 |
| Unintentional injuries excluding drug overdose | #5 | 33,732 | 8.3% | #6 | 121,610 | 3.6% | 0.28 |
| Suicide excluding drug overdose | #6 | 20,493 | 5.0% | #12 | 42,681 | 1.2% | 0.48 |
| Chronic liver disease and cirrhosis | #7 | 17,311 | 4.3% | #10 | 56,196 | 1.6% | 0.31 |
| Homicide excluding drug overdose | #8 | 14,917 | 3.7% | #17 | 24,548 | 0.7% | 0.61 |
| Diabetes mellitus | #9 | 11,182 | 2.8% | #9 | 102,641 | 3.0% | 0.11 |
| Cerebrovascular diseases | #10 | 8,171 | 2.0% | #4 | 161,922 | 4.7% | 0.05 |
| Chronic lower respiratory diseases | #11 | 4,137 | 1.0% | #5 | 141,656 | 4.1% | 0.03 |
| Septicemia | #12 | 3,663 | 0.9% | #15 | 40,947 | 1.2% | 0.09 |
| Nephritis, nephrotic syndrome and nephrosis | #13 | 3,619 | 0.9% | #11 | 54,085 | 1.6% | 0.07 |
| Influenza and pneumonia | #14 | 2,810 | 0.7% | #14 | 41,520 | 1.2% | 0.07 |
| Essential hypertension and<br>hypertensive renal disease | #15 | 2,514 | 0.6% | #13 | 42,625 | 1.2% | 0.06 |
| Human immunodeficiency virus | #16 | 2,178 | 0.5% | #24 | 4,938 | 0.1% | 0.44 |
| Pregnancy, childbirth and the puerperium | #17 | 2,014 | 0.5% | #32 | 1,659 | 0.0% | 1.21 |
| Congenital malformations, deformations<br>and chromosomal abnormalities | #18 | 1,578 | 0.4% | #25 | 4,905 | 0.1% | 0.32 |
| Aortic aneurysm and dissection | #19 | 1,107 | 0.3% | #21 | 9,962 | 0.3% | 0.11 |
| Pneumonitis due to solids and liquids | #20 | 947 | 0.2% | #18 | 19,879 | 0.6% | 0.05 |
| In situ neoplasms, benign neoplasms and<br>neoplasms of uncertain or unknown behavior | #21 | 789 | 0.2% | #20 | 16,067 | 0.5% | 0.05 |
| Complications of medical and surgical care | #22 | 738 | 0.2% | #22 | 5,971 | 0.2% | 0.12 |
| Anemias | #23 | 630 | 0.2% | #23 | 5,928 | 0.2% | 0.11 |
| Viral hepatitis | #24 | 558 | 0.1% | #30 | 3,557 | 0.1% | 0.16 |
| Legal intervention | #25 | 420 | 0.1% | #35 | 648 | 0.0% | 0.65 |
| Peptic ulcer | #26 | 333 | 0.1% | #29 | 3,972 | 0.1% | 0.08 |
| Nutritional deficiencies | #27 | 316 | 0.1% | #19 | 17,433 | 0.5% | 0.02 |
| Alzheimer disease | #28 | 184 | 0.0% | #7 | 119,072 | 3.5% | 0.00 |
| Parkinson disease | #29 | 128 | 0.0% | #16 | 38,446 | 1.1% | 0.00 |
| Cholelithiasis and other disorders of gallbladder | #30 | 123 | 0.0% | #26 | 4,480 | 0.1% | 0.03 |
| Enterocolitis due to Clostridium difficile | #31 | 119 | 0.0% | #28 | 4,094 | 0.1% | 0.03 |
| Infections of kidney | #32 | 89 | 0.0% | #33 | 1,232 | 0.0% | 0.07 |
| Hernia | #33 | 85 | 0.0% | #31 | 2,452 | 0.1% | 0.03 |
| Meningitis | #34 | 45 | 0.0% | #38 | 454 | 0.0% | 0.10 |
| Atherosclerosis | #35 | 40 | 0.0% | #27 | 4,185 | 0.1% | 0.01 |
| Tuberculosis | #36 | 24 | 0.0% | #36 | 595 | 0.0% | 0.04 |
| Inflammatory diseases of female pelvic organs | #37 | 21 | 0.0% | #40 | 211 | 0.0% | 0.10 |
| Acute bronchitis and bronchiolitis | #38 | 0 | 0.0% | #41 | 84 | 0.0% | 0.00 |
| Arthropod-borne viral encephalitis | #39 | 0 | 0.0% | #47 | 6 | 0.0% | 0.00 |
| Certain conditions originating in the<br>perinatal period | #40 | 0 | 0.0% | #43 | 40 | 0.0% | 0.00 |
| Diseases of appendix | #41 | 0 | 0.0% | #39 | 427 | 0.0% | 0.00 |
| Hyperplasia of prostate | #42 | 0 | 0.0% | #34 | 708 | 0.0% | 0.00 |
| Malaria | #43 | 0 | 0.0% | #48 | 6 | 0.0% | 0.00 |
| Meningococcal infection | #44 | 0 | 0.0% | #45 | 18 | 0.0% | 0.00 |
| Operations of war and their sequelae | #45 | 0 | 0.0% | #46 | 9 | 0.0% | 0.00 |
| Pneumoconioses and chemical effects | #46 | 0 | 0.0% | #37 | 562 | 0.0% | 0.00 |
| Salmonella infections | #47 | 0 | 0.0% | #42 | 58 | 0.0% | 0.00 |
| Shigellosis and amebiasis | #48 | 0 | 0.0% | #49 | 6 | 0.0% | 0.00 |
| Syphilis | #49 | 0 | 0.0% | #44 | 38 | 0.0% | 0.00 |
| Whooping cough | #50 | 0 | 0.0% | #50 | 2 | 0.0% | 0.00 |
| Acute poliomyelitis | #51 | 0 | 0.0% | #51 | 0 | 0.0% | 0.00 |
| Measles | #52 | 0 | 0.0% | #52 | 0 | 0.0% | 0.00 |
| Scarlet fever and erysipelas | #53 | 0 | 0.0% | #53 | 0 | 0.0% | 0.00 |
| Others | - | 48,297 | 11.9% | - | 502,198 | 14.7% | 0.10 |

**Supplementary Table S2:**
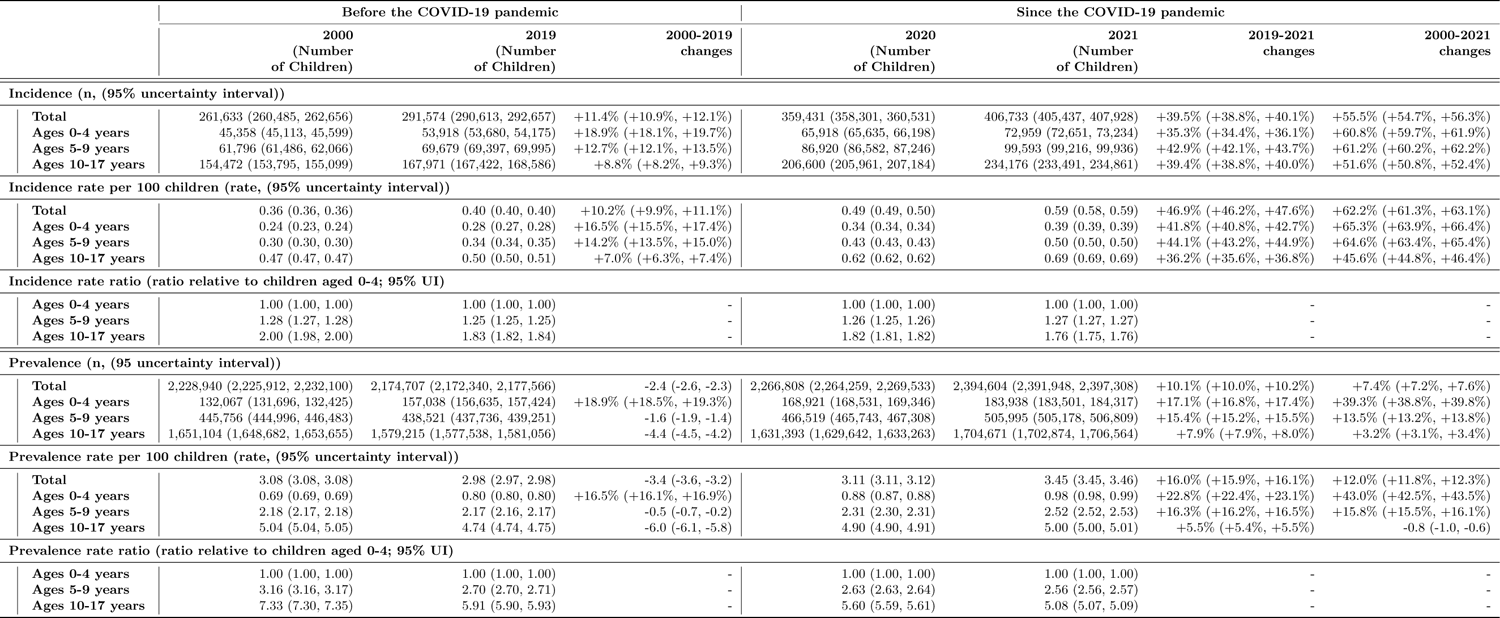
Age groups of children newly experiencing orphanhood in the U.S. from 2000 to 2021.

|  |  | Before the COVID-19 pandemic |  |  | Since the COVID-19 pandemic |  |  |  |
| --- | --- | --- | --- | --- | --- | --- | --- | --- |
|  |  | 2000<br>(Number<br>of Children) | 2019<br>(Number<br>of Children) | 2000-2019<br>changes | 2020<br>(Number<br>of Children) | 2021<br>(Number<br>of Children) | 2019-2021<br>changes | 2000-2021<br>changes |
| Incidence (n, (95% uncertainty interval)) |  |  |  |  |  |  |  |  |
|  | **Total** | 261,633 (260,485, 262,656) | 291,574 (290,613, 292,657) | +11.4% (+10.9%, +12.1%) | 359,431 (358,301, 360,531) | 406,733 (405,437, 407,928) | +39.5% (+38.8%, +40.1%) | +55.5% (+54.7%, +56.3%) |
|  | **Ages 0-4 years** | 45,358 (45,113, 45,599) | 53,918 (53,680, 54,175) | +18.9% (+18.1%, +19.7%) | 65,918 (65,635, 66,198) | 72,959 (72,651, 73,234) | +35.3% (+34.4%, +36.1%) | +60.8% (+59.7%, +61.9%) |
|  | **Ages 5-9 years** | 61,796 (61,486, 62,066) | 69,679 (69,397, 69,995) | +12.7% (+12.1%, +13.5%) | 86,920 (86,582, 87,246) | 99,593 (99,216, 99,936) | +42.9% (+42.1%, +43.7%) | +61.2% (+60.2%, +62.2%) |
|  | **Ages 10-17 years** | 154,472 (153,795, 155,099) | 167,971 (167,422, 168,586) | +8.8% (+8.2%, +9.3%) | 206,600 (205,961, 207,184) | 234,176 (233,491, 234,861) | +39.4% (+38.8%, +40.0%) | +51.6% (+50.8%, +52.4%) |
| Incidence rate per 100 children (rate, (95% uncertainty interval)) |  |  |  |  |  |  |  |  |
|  | **Total** | 0.36 (0.36, 0.36) | 0.40 (0.40, 0.40) | +10.2% (+9.9%, +11.1%) | 0.49 (0.49, 0.50) | 0.59 (0.58, 0.59) | +46.9% (+46.2%, +47.6%) | +62.2% (+61.3%, +63.1%) |
|  | **Ages 0-4 years** | 0.24 (0.23, 0.24) | 0.28 (0.27, 0.28) | +16.5% (+15.5%, +17.4%) | 0.34 (0.34, 0.34) | 0.39 (0.39, 0.39) | +41.8% (+40.8%, +42.7%) | +65.3% (+63.9%, +66.4%) |
|  | **Ages 5-9 years** | 0.30 (0.30, 0.30) | 0.34 (0.34, 0.35) | +14.2% (+13.5%, +15.0%) | 0.43 (0.43, 0.43) | 0.50 (0.50, 0.50) | +44.1% (+43.2%, +44.9%) | +64.6% (+63.4%, +65.4%) |
|  | **Ages 10-17 years** | 0.47 (0.47, 0.47) | 0.50 (0.50, 0.51) | +7.0% (+6.3%, +7.4%) | 0.62 (0.62, 0.62) | 0.69 (0.69, 0.69) | +36.2% (+35.6%, +36.8%) | +45.6% (+44.8%, +46.4%) |
| Incidence rate ratio (ratio relative to children aged 0-4; 95% UI) |  |  |  |  |  |  |  |  |
|  | **Ages 0-4 years** | 1.00 (1.00, 1.00) | 1.00 (1.00, 1.00) | - | 1.00 (1.00, 1.00) | 1.00 (1.00, 1.00) | - | - |
|  | **Ages 5-9 years** | 1.28 (1.27, 1.28) | 1.25 (1.25, 1.25) | - | 1.26 (1.25, 1.26) | 1.27 (1.27, 1.27) | - | - |
|  | **Ages 10-17 years** | 2.00 (1.98, 2.00) | 1.83 (1.82, 1.84) | - | 1.82 (1.81, 1.82) | 1.76 (1.75, 1.76) | - | - |
| Prevalence (n, (95 uncertainty interval)) |  |  |  |  |  |  |  |  |
|  | **Total** | 2,228,940 (2,225,912, 2,232,100) | 2,174,707 (2,172,340, 2,177,566) | -2.4 (-2.6, -2.3) | 2,266,808 (2,264,259, 2,269,533) | 2,394,604 (2,391,948, 2,397,308) | +10.1% (+10.0%, +10.2%) | +7.4% (+7.2%, +7.6%) |
|  | **Ages 0-4 years** | 132,067 (131,696, 132,425) | 157,038 (156,635, 157,424) | +18.9% (+18.5%, +19.3%) | 168,921 (168,531, 169,346) | 183,938 (183,501, 184,317) | +17.1% (+16.8%, +17.4%) | +39.3% (+38.8%, +39.8%) |
|  | **Ages 5-9 years** | 445,756 (444,996, 446,483) | 438,521 (437,736, 439,251) | -1.6 (-1.9, -1.4) | 466,519 (465,743, 467,308) | 505,995 (505,178, 506,809) | +15.4% (+15.2%, +15.5%) | +13.5% (+13.2%, +13.8%) |
|  | **Ages 10-17 years** | 1,651,104 (1,648,682, 1,653,655) | 1,579,215 (1,577,538, 1,581,056) | -4.4 (-4.5, -4.2) | 1,631,393 (1,629,642, 1,633,263) | 1,704,671 (1,702,874, 1,706,564) | +7.9% (+7.9%, +8.0%) | +3.2% (+3.1%, +3.4%) |
| Prevalence rate per 100 children (rate, (95% uncertainty interval)) |  |  |  |  |  |  |  |  |
|  | **Total** | 3.08 (3.08, 3.08) | 2.98 (2.97, 2.98) | -3.4 (-3.6, -3.2) | 3.11 (3.11, 3.12) | 3.45 (3.45, 3.46) | +16.0% (+15.9%, +16.1%) | +12.0% (+11.8%, +12.3%) |
|  | **Ages 0-4 years** | 0.69 (0.69, 0.69) | 0.80 (0.80, 0.80) | +16.5% (+16.1%, +16.9%) | 0.88 (0.87, 0.88) | 0.98 (0.98, 0.99) | +22.8% (+22.4%, +23.1%) | +43.0% (+42.5%, +43.5%) |
|  | **Ages 5-9 years** | 2.18 (2.17, 2.18) | 2.17 (2.16, 2.17) | -0.5 (-0.7, -0.2) | 2.31 (2.30, 2.31) | 2.52 (2.52, 2.53) | +16.3% (+16.2%, +16.5%) | +15.8% (+15.5%, +16.1%) |
|  | **Ages 10-17 years** | 5.04 (5.04, 5.05) | 4.74 (4.74, 4.75) | -6.0 (-6.1, -5.8) | 4.90 (4.90, 4.91) | 5.00 (5.00, 5.01) | +5.5% (+5.4%, +5.5%) | -0.8 (-1.0, -0.6) |
| Prevalence rate ratio (ratio relative to children aged 0-4; 95% UI) |  |  |  |  |  |  |  |  |
|  | **Ages 0-4 years** | 1.00 (1.00, 1.00) | 1.00 (1.00, 1.00) | - | 1.00 (1.00, 1.00) | 1.00 (1.00, 1.00) | - | - |
|  | **Ages 5-9 years** | 3.16 (3.16, 3.17) | 2.70 (2.70, 2.71) | - | 2.63 (2.63, 2.64) | 2.56 (2.56, 2.57) | - | - |
|  | **Ages 10-17 years** | 7.33 (7.30, 7.35) | 5.91 (5.90, 5.93) | - | 5.60 (5.59, 5.61) | 5.08 (5.07, 5.09) | - | - |

**Supplementary Table S3:** Racial and ethnic disparities in children newly experiencing orphanhood in the U.S. from 2000 to 2021.

|  | Before the COVID-19 pandemic |  |  | Since the COVID-19 pandemic |  |  |  |
| --- | --- | --- | --- | --- | --- | --- | --- |
|  | 2000<br>(Number<br>of Children) | 2019<br>(Number<br>of Children) | 2000-2019<br>changes | 2020<br>(Number<br>of Children) | 2021<br>(Number<br>of Children) | 2019-2021<br>changes | 2000-2021<br>changes |
| **Incidence (n, (95% uncertainty interval))** |  |  |  |  |  |  |  |
| **Total** | 261,633 (260,485, 262,656) | 291,574 (290,613, 292,657) | +11.4% (+10.9%, +12.1%) | 359,431 (358,301, 360,531) | 406,733 (405,437, 407,928) | +39.5% (+38.8%, +40.1%) | +55.5% (+54.7%, +56.3%) |
| **Non-Hispanic American Indian or Alaska Native** | 2,529 (2,426, 2,646) | 4,254 (4,116, 4,407) | +68.2% (+58.3%, +77.7%) | 6,228 (6,063, 6,397) | 6,943 (6,771, 7,114) | +63.2% (+56.5%, +70.2%) | +174.7% (+160.7%, +188.4%) |
| **Non-Hispanic Asian** | 5,559 (5,394, 5,719) | 9,147 (8,957, 9,335) | +64.6% (+58.9%, +70.7%) | 11,560 (11,354, 11,787) | 11,873 (11,646, 12,080) | +29.7% (+26.3%, +33.3%) | +113.6% (+106.9%, +120.5%) |
| **Non-Hispanic Black** | 58,337 (57,868, 58,818) | 57,488 (57,074, 57,927) | -1.4 (-2.5, -0.3) | 75,409 (74,894, 75,952) | 84,023 (83,464, 84,568) | +46.2% (+44.7%, +47.6%) | +44.0% (+42.5%, +45.6%) |
| **Hispanic** | 31,027 (30,666, 31,412) | 46,680 (46,237, 47,138) | +50.5% (+48.2%, +53.0%) | 66,849 (66,320, 67,377) | 77,977 (77,333, 78,523) | +67.0% (+64.9%, +69.0%) | +151.4% (+147.8%, +154.8%) |
| **Non-Hispanic White** | 164,168 (163,296, 164,974) | 173,997 (173,269, 174,787) | +6.0% (+5.3%, +6.7%) | 199,394 (198,551, 200,192) | 225,924 (224,990, 226,794) | +29.8% (+29.1%, +30.6%) | +37.6% (+36.7%, +38.4%) |
| **Incidence rate per 100 children (rate, (95% uncertainty interval))** |  |  |  |  |  |  |  |
| **Total** | 0.36 (0.36, 0.36) | 0.40 (0.40, 0.40) | +10.2% (+9.9%, +11.1%) | 0.49 (0.49, 0.50) | 0.59 (0.58, 0.59) | +46.9% (+46.2%, +47.6%) | +62.2% (+61.3%, +63.1%) |
| **Non-Hispanic American Indian or Alaska Native** | 0.32 (0.31, 0.34) | 0.60 (0.58, 0.62) | +85.2% (+74.4%, +95.6%) | 0.88 (0.86, 0.91) | 1.21 (1.18, 1.24) | +101.5% (+93.2%, +110.0%) | +273.4% (+254.1%, +292.2%) |
| **Non-Hispanic Asian** | 0.19 (0.19, 0.20) | 0.20 (0.20, 0.20) | +4.6% (+1.0%, +8.5%) | 0.25 (0.25, 0.26) | 0.29 (0.28, 0.29) | +42.5% (+38.9%, +46.5%) | +49.0% (+44.5%, +54.0%) |
| **Non-Hispanic Black** | 0.52 (0.52, 0.52) | 0.52 (0.51, 0.52) | -0.8 (-1.9, +0.4%) | 0.68 (0.67, 0.68) | 0.83 (0.83, 0.84) | +61.6% (+59.9%, +63.2%) | +60.3% (+58.6%, +62.1%) |
| **Hispanic** | 0.25 (0.25, 0.25) | 0.25 (0.25, 0.25) | +0.8% (-0.8, +2.4%) | 0.36 (0.36, 0.36) | 0.42 (0.41, 0.42) | +65.6% (+63.5%, +67.6%) | +66.9% (+64.5%, +69.2%) |
| **Non-Hispanic White** | 0.36 (0.36, 0.37) | 0.46 (0.46, 0.46) | +25.3% (+24.4%, +26.2%) | 0.53 (0.53, 0.53) | 0.63 (0.63, 0.63) | +37.9% (+37.1%, +38.7%) | +72.9% (+71.6%, +73.9%) |
| **Incidence rate ratio (ratio relative to Non-Hispanic White children; 95% UI)** |  |  |  |  |  |  |  |
| **Non-Hispanic American Indian or Alaska Native** | 1.68 (1.60, 1.78) | 2.98 (2.86, 3.10) | - | 3.50 (3.40, 3.63) | 4.21 (4.08, 4.35) | - | - |
| **Non-Hispanic Asian** | 1.00 (1.00, 1.00) | 1.00 (1.00, 1.00) | - | 1.00 (1.00, 1.00) | 1.00 (1.00, 1.00) | - | - |
| **Non-Hispanic Black** | 2.70 (2.62, 2.79) | 2.57 (2.52, 2.63) | - | 2.69 (2.63, 2.74) | 2.91 (2.86, 2.97) | - | - |
| **Hispanic** | 1.30 (1.26, 1.34) | 1.25 (1.22, 1.28) | - | 1.42 (1.39, 1.45) | 1.45 (1.42, 1.48) | - | - |
| **Non-Hispanic White** | 1.90 (1.84, 1.96) | 2.27 (2.22, 2.32) | - | 2.09 (2.05, 2.13) | 2.20 (2.16, 2.24) | - | - |
| **Prevalence (n, (95% uncertainty interval))** |  |  |  |  |  |  |  |
| **Total** | 2,228,940 (2,225,912, 2,232,100) | 2,174,707 (2,172,340, 2,177,566) | -2.4 (-2.6, -2.3) | 2,266,808 (2,264,259, 2,269,533) | 2,394,604 (2,391,948, 2,397,308) | +10.1% (+10.0%, +10.2%) | +7.4% (+7.2%, +7.6%) |
| **Non-Hispanic American Indian or Alaska Native** | 20,891 (20,547, 21,233) | 31,280 (30,928, 31,614) | +49.8% (+46.8%, +52.7%) | 34,169 (33,793, 34,524) | 37,545 (37,150, 37,936) | +20.1% (+19.1%, +21.0%) | +79.7% (+76.3%, +83.2%) |
| **Non-Hispanic Asian** | 39,880 (39,460, 40,334) | 61,105 (60,619, 61,555) | +53.2% (+51.2%, +55.4%) | 65,307 (64,808, 65,744) | 69,235 (68,753, 69,717) | +13.3% (+12.8%, +13.8%) | +73.6% (+71.3%, +76.0%) |
| **Non-Hispanic Black** | 537,243 (535,666, 538,833) | 424,647 (423,574, 425,780) | -21.0 (-21.3, -20.6) | 449,171 (448,091, 450,349) | 480,610 (479,510, 481,790) | +13.2% (+13.0%, +13.4%) | -10.6 (-10.9, -10.2) |
| **Hispanic** | 270,638 (269,515, 271,850) | 338,784 (337,712, 339,877) | +25.2% (+24.5%, +25.8%) | 366,635 (365,440, 367,796) | 402,161 (400,924, 403,344) | +18.7% (+18.4%, +19.0%) | +48.6% (+47.8%, +49.4%) |
| **Non-Hispanic White** | 1,360,301 (1,357,973, 1,362,583) | 1,318,925 (1,317,104, 1,320,941) | -3.0 (-3.2, -2.8) | 1,351,530 (1,349,757, 1,353,610) | 1,405,015 (1,403,095, 1,406,966) | +6.5% (+6.4%, +6.6%) | +3.3% (+3.1%, +3.5%) |
| **Prevalence rate per 100 children (rate, (95% uncertainty interval))** |  |  |  |  |  |  |  |
| **Total** | 3.08 (3.08, 3.08) | 2.98 (2.97, 2.98) | -3.38 (-3.57, -3.22) | 3.11 (3.11, 3.12) | 3.45 (3.45, 3.46) | +15.97% (+15.88%, +16.07%) | +12.05% (+11.84%, +12.26%) |
| **Non-Hispanic American Indian or Alaska Native** | 2.67 (2.63, 2.71) | 4.40 (4.35, 4.45) | +64.89% (+61.59%, +68.16%) | 4.85 (4.80, 4.90) | 6.52 (6.45, 6.59) | +48.22% (+47.09%, +49.39%) | +144.29% (+139.61%, +148.99%) |
| **Non-Hispanic Asian** | 1.38 (1.36, 1.40) | 1.34 (1.33, 1.35) | -2.62 (-3.95, -1.25) | 1.43 (1.41, 1.43) | 1.67 (1.66, 1.68) | +24.46% (+23.88%, +25.02%) | +21.20% (+19.55%, +22.81%) |
| **Non-Hispanic Black** | 4.79 (4.78, 4.81) | 3.82 (3.81, 3.83) | -20.41 (-20.70, -20.08) | 4.04 (4.03, 4.05) | 4.77 (4.76, 4.78) | +25.10% (+24.89%, +25.32%) | -0.42 (-0.81, -0.02) |
| **Hispanic** | 2.17 (2.16, 2.18) | 1.82 (1.82, 1.83) | -16.17 (-16.62, -15.73) | 1.97 (1.96, 1.97) | 2.15 (2.14, 2.15) | +17.72% (+17.47%, +17.97%) | -1.33 (-1.88, -0.79) |
| **Non-Hispanic White** | 3.02 (3.02, 3.02) | 3.46 (3.46, 3.47) | +14.61% (+14.36%, +14.88%) | 3.58 (3.57, 3.58) | 3.92 (3.91, 3.92) | +13.20% (+13.09%, +13.30%) | +29.74% (+29.44%, +30.02%) |
| **Prevalence rate ratio (ratio relative to Non-Hispanic White children; 95% UI)** |  |  |  |  |  |  |  |
| **Non-Hispanic American Indian or Alaska Native** | 1.93 (1.90, 1.97) | 3.28 (3.23, 3.32) | - | 3.41 (3.36, 3.45) | 3.90 (3.85, 3.95) | - | - |
| **Non-Hispanic Asian** | 1.00 (1.00, 1.00) | 1.00 (1.00, 1.00) | - | 1.00 (1.00, 1.00) | 1.00 (1.00, 1.00) | - | - |
| **Non-Hispanic Black** | 3.47 (3.43, 3.51) | 2.84 (2.82, 2.86) | - | 2.83 (2.81, 2.86) | 2.86 (2.83, 2.88) | - | - |
| **Hispanic** | 1.58 (1.56, 1.59) | 1.36 (1.35, 1.37) | - | 1.38 (1.37, 1.39) | 1.28 (1.27, 1.29) | - | - |
| **Non-Hispanic White** | 2.19 (2.16, 2.21) | 2.58 (2.56, 2.60) | - | 2.51 (2.49, 2.53) | 2.34 (2.33, 2.36) | - | - |

**Supplementary Table S4:**
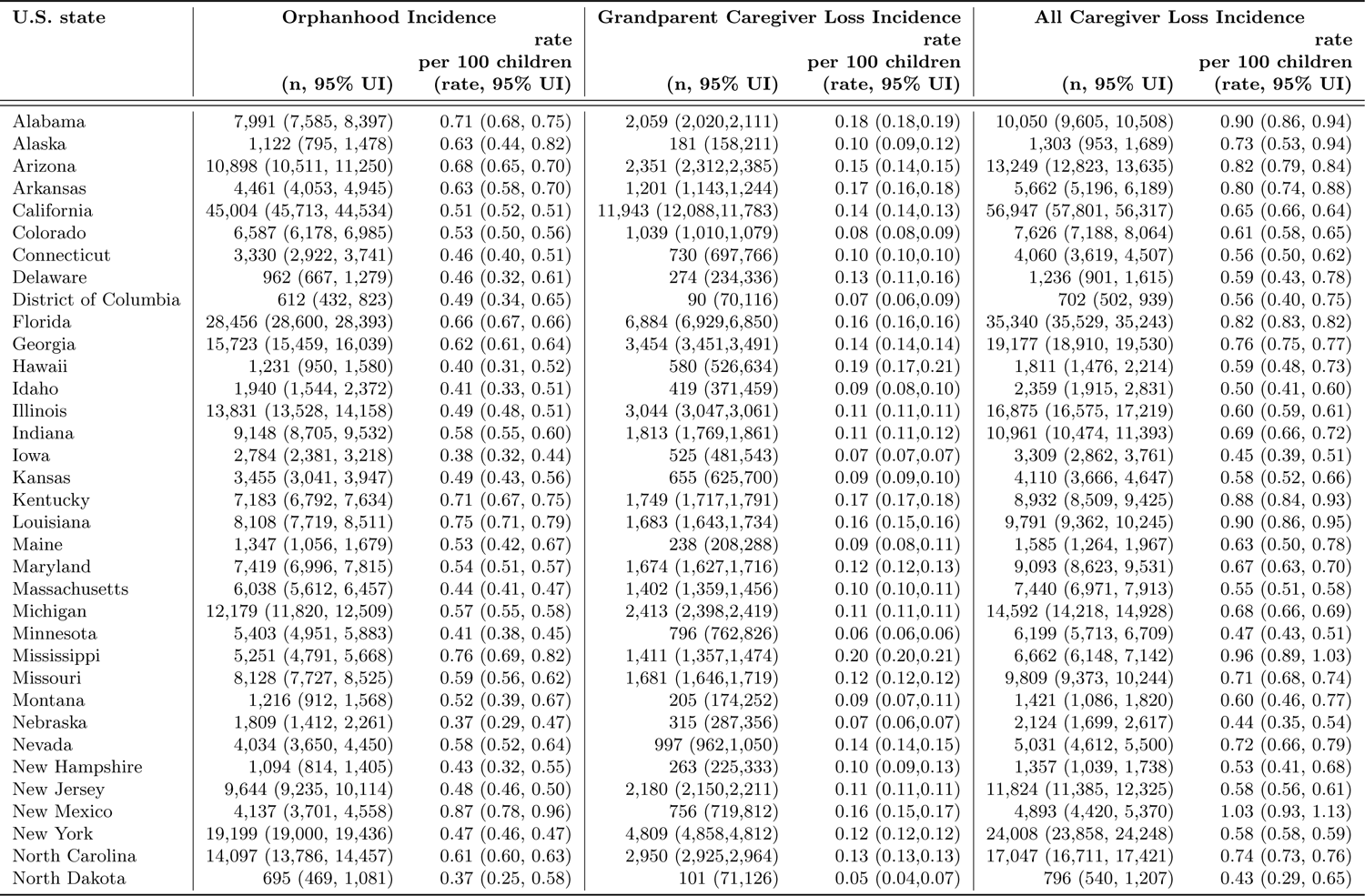
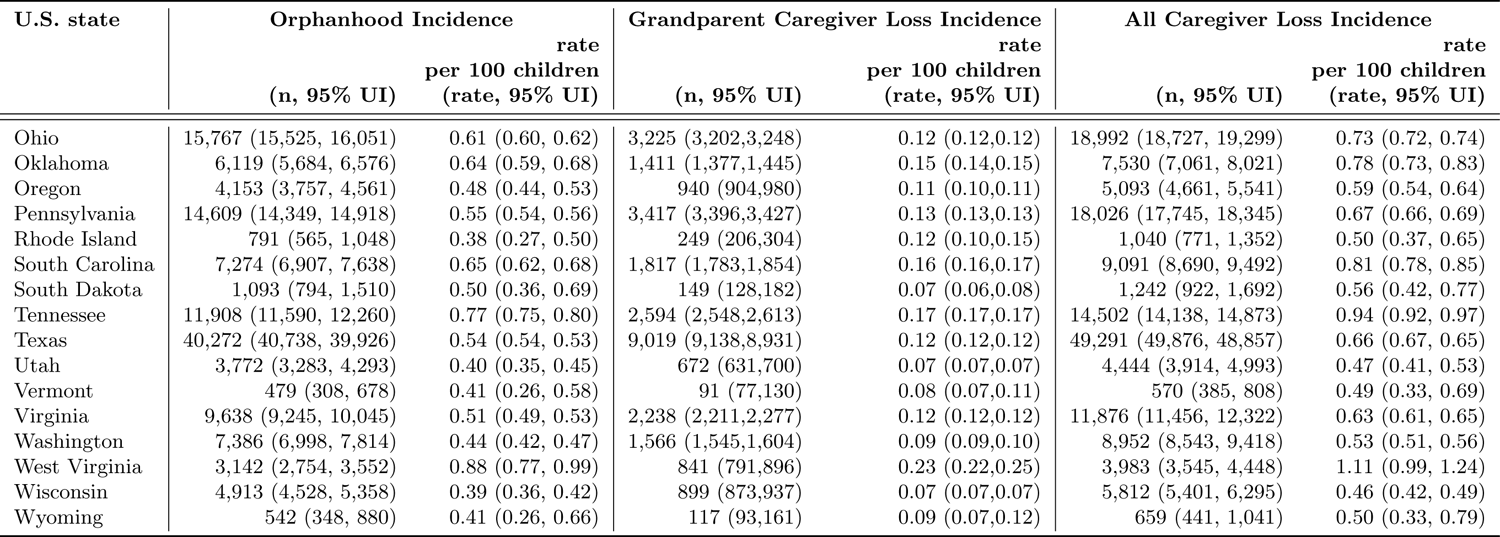
Incidence of orphanhood, grandparent caregiver loss and all caregiver loss in 2021 by U.S. states.

| U.S. state | Orphanhood Incidence |  | Grandparent Caregiver Loss Incidence |  | All Caregiver Loss Incidence |  |
| --- | --- | --- | --- | --- | --- | --- |
|  | (n, 95% UI) | rate<br>per 100 children<br>(rate, 95% UI) | (n, 95% UI) | rate<br>per 100 children<br>(rate, 95% UI) | (n, 95% UI) | rate<br>per 100 children<br>(rate, 95% UI) |
| Alabama | 7,991 (7,585, 8,397) | 0.71 (0.68, 0.75) | 2,059 (2,020,2,111) | 0.18 (0.18,0.19) | 10,050 (9,605, 10,508) | 0.90 (0.86, 0.94) |
| Alaska | 1,122 (795, 1,478) | 0.63 (0.44, 0.82) | 181 (158,211) | 0.10 (0.09,0.12) | 1,303 (953, 1,689) | 0.73 (0.53, 0.94) |
| Arizona | 10,898 (10,511, 11,250) | 0.68 (0.65, 0.70) | 2,351 (2,312,2,385) | 0.15 (0.14,0.15) | 13,249 (12,823, 13,635) | 0.82 (0.79, 0.84) |
| Arkansas | 4,461 (4,053, 4,945) | 0.63 (0.58, 0.70) | 1,201 (1,143,1,244) | 0.17 (0.16,0.18) | 5,662 (5,196, 6,189) | 0.80 (0.74, 0.88) |
| California | 45,004 (45,713, 44,534) | 0.51 (0.52, 0.51) | 11,943 (12,088,11,783) | 0.14 (0.14,0.13) | 56,947 (57,801, 56,317) | 0.65 (0.66, 0.64) |
| Colorado | 6,587 (6,178, 6,985) | 0.53 (0.50, 0.56) | 1,039 (1,010,1,079) | 0.08 (0.08,0.09) | 7,626 (7,188, 8,064) | 0.61 (0.58, 0.65) |
| Connecticut | 3,330 (2,922, 3,741) | 0.46 (0.40, 0.51) | 730 (697,766) | 0.10 (0.10,0.10) | 4,060 (3,619, 4,507) | 0.56 (0.50, 0.62) |
| Delaware | 962 (667, 1,279) | 0.46 (0.32, 0.61) | 274 (234,336) | 0.13 (0.11,0.16) | 1,236 (901, 1,615) | 0.59 (0.43, 0.78) |
| District of Columbia | 612 (432, 823) | 0.49 (0.34, 0.65) | 90 (70,116) | 0.07 (0.06,0.09) | 702 (502, 939) | 0.56 (0.40, 0.75) |
| Florida | 28,456 (28,600, 28,393) | 0.66 (0.67, 0.66) | 6,884 (6,929,6,850) | 0.16 (0.16,0.16) | 35,340 (35,529, 35,243) | 0.82 (0.83, 0.82) |
| Georgia | 15,723 (15,459, 16,039) | 0.62 (0.61, 0.64) | 3,454 (3,451,3,491) | 0.14 (0.14,0.14) | 19,177 (18,910, 19,530) | 0.76 (0.75, 0.77) |
| Hawaii | 1,231 (950, 1,580) | 0.40 (0.31, 0.52) | 580 (526,634) | 0.19 (0.17,0.21) | 1,811 (1,476, 2,214) | 0.59 (0.48, 0.73) |
| Idaho | 1,940 (1,544, 2,372) | 0.41 (0.33, 0.51) | 419 (371,459) | 0.09 (0.08,0.10) | 2,359 (1,915, 2,831) | 0.50 (0.41, 0.60) |
| Illinois | 13,831 (13,528, 14,158) | 0.49 (0.48, 0.51) | 3,044 (3,047,3,061) | 0.11 (0.11,0.11) | 16,875 (16,575, 17,219) | 0.60 (0.59, 0.61) |
| Indiana | 9,148 (8,705, 9,532) | 0.58 (0.55, 0.60) | 1,813 (1,769,1,861) | 0.11 (0.11,0.12) | 10,961 (10,474, 11,393) | 0.69 (0.66, 0.72) |
| Iowa | 2,784 (2,381, 3,218) | 0.38 (0.32, 0.44) | 525 (481,543) | 0.07 (0.07,0.07) | 3,309 (2,862, 3,761) | 0.45 (0.39, 0.51) |
| Kansas | 3,455 (3,041, 3,947) | 0.49 (0.43, 0.56) | 655 (625,700) | 0.09 (0.09,0.10) | 4,110 (3,666, 4,647) | 0.58 (0.52, 0.66) |
| Kentucky | 7,183 (6,792, 7,634) | 0.71 (0.67, 0.75) | 1,749 (1,717,1,791) | 0.17 (0.17,0.18) | 8,932 (8,509, 9,425) | 0.88 (0.84, 0.93) |
| Louisiana | 8,108 (7,719, 8,511) | 0.75 (0.71, 0.79) | 1,683 (1,643,1,734) | 0.16 (0.15,0.16) | 9,791 (9,362, 10,245) | 0.90 (0.86, 0.95) |
| Maine | 1,347 (1,056, 1,679) | 0.53 (0.42, 0.67) | 238 (208,288) | 0.09 (0.08,0.11) | 1,585 (1,264, 1,967) | 0.63 (0.50, 0.78) |
| Maryland | 7,419 (6,996, 7,815) | 0.54 (0.51, 0.57) | 1,674 (1,627,1,716) | 0.12 (0.12,0.13) | 9,093 (8,623, 9,531) | 0.67 (0.63, 0.70) |
| Massachusetts | 6,038 (5,612, 6,457) | 0.44 (0.41, 0.47) | 1,402 (1,359,1,456) | 0.10 (0.10,0.11) | 7,440 (6,971, 7,913) | 0.55 (0.51, 0.58) |
| Michigan | 12,179 (11,820, 12,509) | 0.57 (0.55, 0.58) | 2,413 (2,398,2,419) | 0.11 (0.11,0.11) | 14,592 (14,218, 14,928) | 0.68 (0.66, 0.69) |
| Minnesota | 5,403 (4,951, 5,883) | 0.41 (0.38, 0.45) | 796 (762,826) | 0.06 (0.06,0.06) | 6,199 (5,713, 6,709) | 0.47 (0.43, 0.51) |
| Mississippi | 5,251 (4,791, 5,668) | 0.76 (0.69, 0.82) | 1,411 (1,357,1,474) | 0.20 (0.20,0.21) | 6,662 (6,148, 7,142) | 0.96 (0.89, 1.03) |
| Missouri | 8,128 (7,727, 8,525) | 0.59 (0.56, 0.62) | 1,681 (1,646,1,719) | 0.12 (0.12,0.12) | 9,809 (9,373, 10,244) | 0.71 (0.68, 0.74) |
| Montana | 1,216 (912, 1,568) | 0.52 (0.39, 0.67) | 205 (174,252) | 0.09 (0.07,0.11) | 1,421 (1,086, 1,820) | 0.60 (0.46, 0.77) |
| Nebraska | 1,809 (1,412, 2,261) | 0.37 (0.29, 0.47) | 315 (287,356) | 0.07 (0.06,0.07) | 2,124 (1,699, 2,617) | 0.44 (0.35, 0.54) |
| Nevada | 4,034 (3,650, 4,450) | 0.58 (0.52, 0.64) | 997 (962,1,050) | 0.14 (0.14,0.15) | 5,031 (4,612, 5,500) | 0.72 (0.66, 0.79) |
| New Hampshire | 1,094 (814, 1,405) | 0.43 (0.32, 0.55) | 263 (225,333) | 0.10 (0.09,0.13) | 1,357 (1,039, 1,738) | 0.53 (0.41, 0.68) |
| New Jersey | 9,644 (9,235, 10,114) | 0.48 (0.46, 0.50) | 2,180 (2,150,2,211) | 0.11 (0.11,0.11) | 11,824 (11,385, 12,325) | 0.58 (0.56, 0.61) |
| New Mexico | 4,137 (3,701, 4,558) | 0.87 (0.78, 0.96) | 756 (719,812) | 0.16 (0.15,0.17) | 4,893 (4,420, 5,370) | 1.03 (0.93, 1.13) |
| New York | 19,199 (19,000, 19,436) | 0.47 (0.46, 0.47) | 4,809 (4,858,4,812) | 0.12 (0.12,0.12) | 24,008 (23,858, 24,248) | 0.58 (0.58, 0.59) |
| North Carolina | 14,097 (13,786, 14,457) | 0.61 (0.60, 0.63) | 2,950 (2,925,2,964) | 0.13 (0.13,0.13) | 17,047 (16,711, 17,421) | 0.74 (0.73, 0.76) |
| North Dakota | 695 (469, 1,081) | 0.37 (0.25, 0.58) | 101 (71,126) | 0.05 (0.04,0.07) | 796 (540, 1,207) | 0.43 (0.29, 0.65) |
|  | (n, 95% UI) | rate<br>per 100 children<br>(rate, 95% UI) | (n, 95% UI) | rate<br>per 100 children<br>(rate, 95% UI) | (n, 95% UI) | rate<br>per 100 children<br>(rate, 95% UI) |
| Ohio | 15,767 (15,525, 16,051) | 0.61 (0.60, 0.62) | 3,225 (3,202,3,248) | 0.12 (0.12,0.12) | 18,992 (18,727, 19,299) | 0.73 (0.72, 0.74) |
| Oklahoma | 6,119 (5,684, 6,576) | 0.64 (0.59, 0.68) | 1,411 (1,377,1,445) | 0.15 (0.14,0.15) | 7,530 (7,061, 8,021) | 0.78 (0.73, 0.83) |
| Oregon | 4,153 (3,757, 4,561) | 0.48 (0.44, 0.53) | 940 (904,980) | 0.11 (0.10,0.11) | 5,093 (4,661, 5,541) | 0.59 (0.54, 0.64) |
| Pennsylvania | 14,609 (14,349, 14,918) | 0.55 (0.54, 0.56) | 3,417 (3,396,3,427) | 0.13 (0.13,0.13) | 18,026 (17,745, 18,345) | 0.67 (0.66, 0.69) |
| Rhode Island | 791 (565, 1,048) | 0.38 (0.27, 0.50) | 249 (206,304) | 0.12 (0.10,0.15) | 1,040 (771, 1,352) | 0.50 (0.37, 0.65) |
| South Carolina | 7,274 (6,907, 7,638) | 0.65 (0.62, 0.68) | 1,817 (1,783,1,854) | 0.16 (0.16,0.17) | 9,091 (8,690, 9,492) | 0.81 (0.78, 0.85) |
| South Dakota | 1,093 (794, 1,510) | 0.50 (0.36, 0.69) | 149 (128,182) | 0.07 (0.06,0.08) | 1,242 (922, 1,692) | 0.56 (0.42, 0.77) |
| Tennessee | 11,908 (11,590, 12,260) | 0.77 (0.75, 0.80) | 2,594 (2,548,2,613) | 0.17 (0.17,0.17) | 14,502 (14,138, 14,873) | 0.94 (0.92, 0.97) |
| Texas | 40,272 (40,738, 39,926) | 0.54 (0.54, 0.53) | 9,019 (9,138,8,931) | 0.12 (0.12,0.12) | 49,291 (49,876, 48,857) | 0.66 (0.67, 0.65) |
| Utah | 3,772 (3,283, 4,293) | 0.40 (0.35, 0.45) | 672 (631,700) | 0.07 (0.07,0.07) | 4,444 (3,914, 4,993) | 0.47 (0.41, 0.53) |
| Vermont | 479 (308, 678) | 0.41 (0.26, 0.58) | 91 (77,130) | 0.08 (0.07,0.11) | 570 (385, 808) | 0.49 (0.33, 0.69) |
| Virginia | 9,638 (9,245, 10,045) | 0.51 (0.49, 0.53) | 2,238 (2,211,2,277) | 0.12 (0.12,0.12) | 11,876 (11,456, 12,322) | 0.63 (0.61, 0.65) |
| Washington | 7,386 (6,998, 7,814) | 0.44 (0.42, 0.47) | 1,566 (1,545,1,604) | 0.09 (0.09,0.10) | 8,952 (8,543, 9,418) | 0.53 (0.51, 0.56) |
| West Virginia | 3,142 (2,754, 3,552) | 0.88 (0.77, 0.99) | 841 (791,896) | 0.23 (0.22,0.25) | 3,983 (3,545, 4,448) | 1.11 (0.99, 1.24) |
| Wisconsin | 4,913 (4,528, 5,358) | 0.39 (0.36, 0.42) | 899 (873,937) | 0.07 (0.07,0.07) | 5,812 (5,401, 6,295) | 0.46 (0.42, 0.49) |
| Wyoming | 542 (348, 880) | 0.41 (0.26, 0.66) | 117 (93,161) | 0.09 (0.07,0.12) | 659 (441, 1,041) | 0.50 (0.33, 0.79) |

**Supplementary Table S5:**
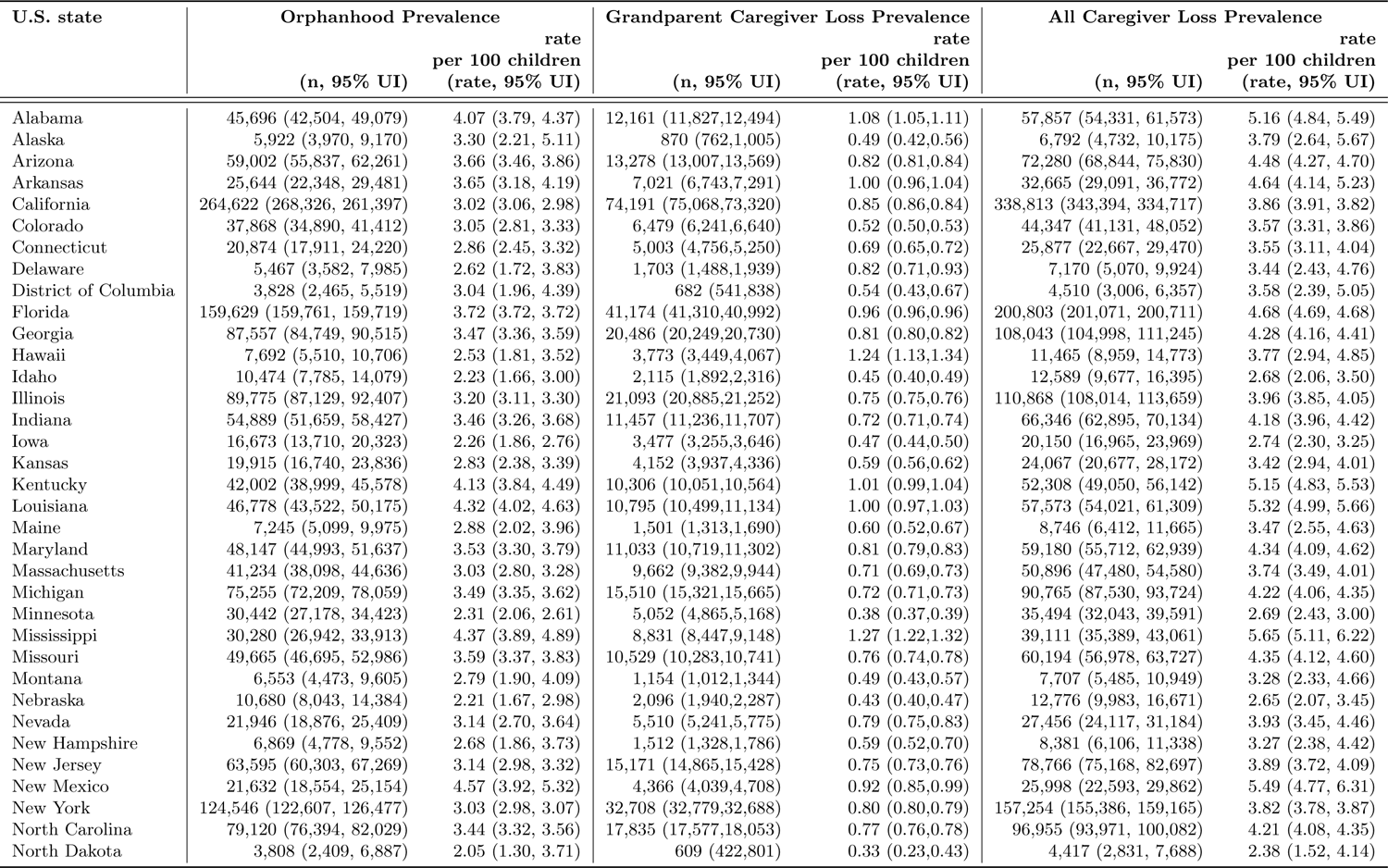
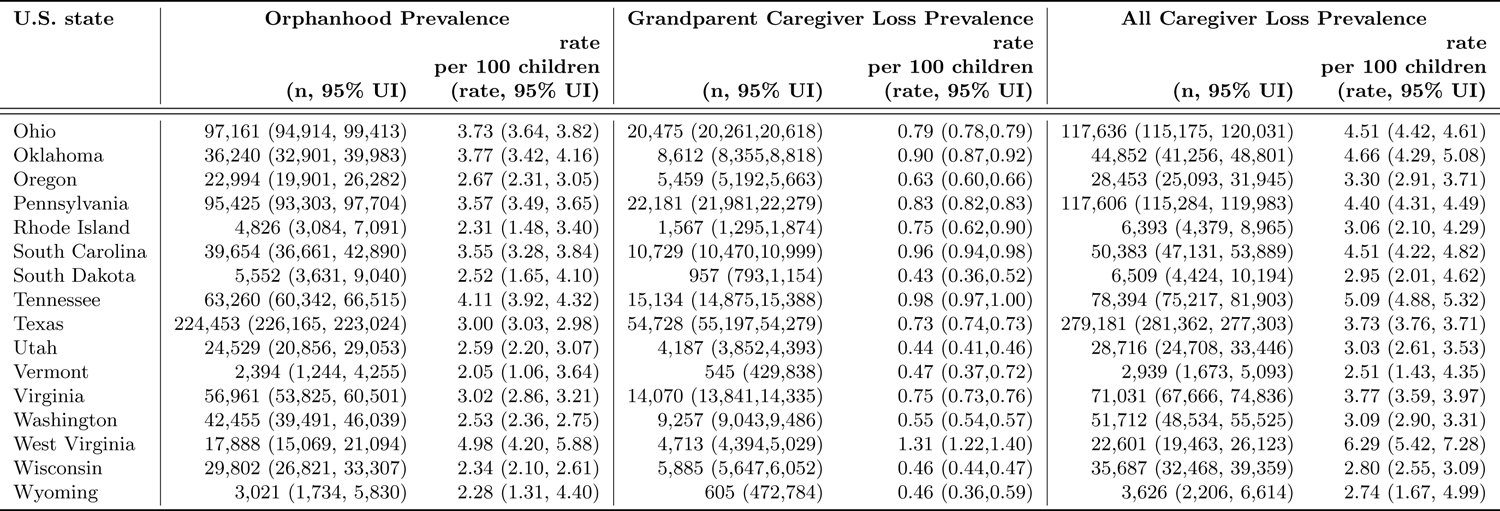
Prevalence of orphanhood, grandparent caregiver loss and all caregiver loss in 2021 by U.S. states.

| U.S. state | Orphanhood Prevalence |  | Grandparent Caregiver Loss Prevalence |  | All Caregiver Loss Prevalence |  |
| --- | --- | --- | --- | --- | --- | --- |
|  |  | rate<br>per 100 children |  | rate<br>per 100 children |  | rate<br>per 100 children |
|  | (n, 95% UI) | (rate, 95% UI) | (n, 95% UI) | (rate, 95% UI) | (n, 95% UI) | (rate, 95% UI) |
| Alabama | 45,696 (42,504, 49,079) | 4.07 (3.79, 4.37) | 12,161 (11,827,12,494) | 1.08 (1.05,1.11) | 57,857 (54,331, 61,573) | 5.16 (4.84, 5.49) |
| Alaska | 5,922 (3,970, 9,170) | 3.30 (2.21, 5.11) | 870 (762,1,005) | 0.49 (0.42,0.56) | 6,792 (4,732, 10,175) | 3.79 (2.64, 5.67) |
| Arizona | 59,002 (55,837, 62,261) | 3.66 (3.46, 3.86) | 13,278 (13,007,13,569) | 0.82 (0.81,0.84) | 72,280 (68,844, 75,830) | 4.48 (4.27, 4.70) |
| Arkansas | 25,644 (22,348, 29,481) | 3.65 (3.18, 4.19) | 7,021 (6,743,7,291) | 1.00 (0.96,1.04) | 32,665 (29,091, 36,772) | 4.64 (4.14, 5.23) |
| California | 264,622 (268,326, 261,397) | 3.02 (3.06, 2.98) | 74,191 (75,068,73,320) | 0.85 (0.86,0.84) | 338,813 (343,394, 334,717) | 3.86 (3.91, 3.82) |
| Colorado | 37,868 (34,890, 41,412) | 3.05 (2.81, 3.33) | 6,479 (6,241,6,640) | 0.52 (0.50,0.53) | 44,347 (41,131, 48,052) | 3.57 (3.31, 3.86) |
| Connecticut | 20,874 (17,911, 24,220) | 2.86 (2.45, 3.32) | 5,003 (4,756,5,250) | 0.69 (0.65,0.72) | 25,877 (22,667, 29,470) | 3.55 (3.11, 4.04) |
| Delaware | 5,467 (3,582, 7,985) | 2.62 (1.72, 3.83) | 1,703 (1,488,1,939) | 0.82 (0.71,0.93) | 7,170 (5,070, 9,924) | 3.44 (2.43, 4.76) |
| District of Columbia | 3,828 (2,465, 5,519) | 3.04 (1.96, 4.39) | 682 (541,838) | 0.54 (0.43,0.67) | 4,510 (3,006, 6,357) | 3.58 (2.39, 5.05) |
| Florida | 159,629 (159,761, 159,719) | 3.72 (3.72, 3.72) | 41,174 (41,310,40,992) | 0.96 (0.96,0.96) | 200,803 (201,071, 200,711) | 4.68 (4.69, 4.68) |
| Georgia | 87,557 (84,749, 90,515) | 3.47 (3.36, 3.59) | 20,486 (20,249,20,730) | 0.81 (0.80,0.82) | 108,043 (104,998, 111,245) | 4.28 (4.16, 4.41) |
| Hawaii | 7,692 (5,510, 10,706) | 2.53 (1.81, 3.52) | 3,773 (3,449,4,067) | 1.24 (1.13,1.34) | 11,465 (8,959, 14,773) | 3.77 (2.94, 4.85) |
| Idaho | 10,474 (7,785, 14,079) | 2.23 (1.66, 3.00) | 2,115 (1,892,2,316) | 0.45 (0.40,0.49) | 12,589 (9,677, 16,395) | 2.68 (2.06, 3.50) |
| Illinois | 89,775 (87,129, 92,407) | 3.20 (3.11, 3.30) | 21,093 (20,885,21,252) | 0.75 (0.75,0.76) | 110,868 (108,014, 113,659) | 3.96 (3.85, 4.05) |
| Indiana | 54,889 (51,659, 58,427) | 3.46 (3.26, 3.68) | 11,457 (11,236,11,707) | 0.72 (0.71,0.74) | 66,346 (62,895, 70,134) | 4.18 (3.96, 4.42) |
| Iowa | 16,673 (13,710, 20,323) | 2.26 (1.86, 2.76) | 3,477 (3,255,3,646) | 0.47 (0.44,0.50) | 20,150 (16,965, 23,969) | 2.74 (2.30, 3.25) |
| Kansas | 19,915 (16,740, 23,836) | 2.83 (2.38, 3.39) | 4,152 (3,937,4,336) | 0.59 (0.56,0.62) | 24,067 (20,677, 28,172) | 3.42 (2.94, 4.01) |
| Kentucky | 42,002 (38,999, 45,578) | 4.13 (3.84, 4.49) | 10,306 (10,051,10,564) | 1.01 (0.99,1.04) | 52,308 (49,050, 56,142) | 5.15 (4.83, 5.53) |
| Louisiana | 46,778 (43,522, 50,175) | 4.32 (4.02, 4.63) | 10,795 (10,499,11,134) | 1.00 (0.97,1.03) | 57,573 (54,021, 61,309) | 5.32 (4.99, 5.66) |
| Maine | 7,245 (5,099, 9,975) | 2.88 (2.02, 3.96) | 1,501 (1,313,1,690) | 0.60 (0.52,0.67) | 8,746 (6,412, 11,665) | 3.47 (2.55, 4.63) |
| Maryland | 48,147 (44,993, 51,637) | 3.53 (3.30, 3.79) | 11,033 (10,719,11,302) | 0.81 (0.79,0.83) | 59,180 (55,712, 62,939) | 4.34 (4.09, 4.62) |
| Massachusetts | 41,234 (38,098, 44,636) | 3.03 (2.80, 3.28) | 9,662 (9,382,9,944) | 0.71 (0.69,0.73) | 50,896 (47,480, 54,580) | 3.74 (3.49, 4.01) |
| Michigan | 75,255 (72,209, 78,059) | 3.49 (3.35, 3.62) | 15,510 (15,321,15,665) | 0.72 (0.71,0.73) | 90,765 (87,530, 93,724) | 4.22 (4.06, 4.35) |
| Minnesota | 30,442 (27,178, 34,423) | 2.31 (2.06, 2.61) | 5,052 (4,865,5,168) | 0.38 (0.37,0.39) | 35,494 (32,043, 39,591) | 2.69 (2.43, 3.00) |
| Mississippi | 30,280 (26,942, 33,913) | 4.37 (3.89, 4.89) | 8,831 (8,447,9,148) | 1.27 (1.22,1.32) | 39,111 (35,389, 43,061) | 5.65 (5.11, 6.22) |
| Missouri | 49,665 (46,695, 52,986) | 3.59 (3.37, 3.83) | 10,529 (10,283,10,741) | 0.76 (0.74,0.78) | 60,194 (56,978, 63,727) | 4.35 (4.12, 4.60) |
| Montana | 6,553 (4,473, 9,605) | 2.79 (1.90, 4.09) | 1,154 (1,012,1,344) | 0.49 (0.43,0.57) | 7,707 (5,485, 10,949) | 3.28 (2.33, 4.66) |
| Nebraska | 10,680 (8,043, 14,384) | 2.21 (1.67, 2.98) | 2,096 (1,940,2,287) | 0.43 (0.40,0.47) | 12,776 (9,983, 16,671) | 2.65 (2.07, 3.45) |
| Nevada | 21,946 (18,876, 25,409) | 3.14 (2.70, 3.64) | 5,510 (5,241,5,775) | 0.79 (0.75,0.83) | 27,456 (24,117, 31,184) | 3.93 (3.45, 4.46) |
| New Hampshire | 6,869 (4,778, 9,552) | 2.68 (1.86, 3.73) | 1,512 (1,328,1,786) | 0.59 (0.52,0.70) | 8,381 (6,106, 11,338) | 3.27 (2.38, 4.42) |
| New Jersey | 63,595 (60,303, 67,269) | 3.14 (2.98, 3.32) | 15,171 (14,865,15,428) | 0.75 (0.73,0.76) | 78,766 (75,168, 82,697) | 3.89 (3.72, 4.09) |
| New Mexico | 21,632 (18,554, 25,154) | 4.57 (3.92, 5.32) | 4,366 (4,039,4,708) | 0.92 (0.85,0.99) | 25,998 (22,593, 29,862) | 5.49 (4.77, 6.31) |
| New York | 124,546 (122,607, 126,477) | 3.03 (2.98, 3.07) | 32,708 (32,779,32,688) | 0.80 (0.80,0.79) | 157,254 (155,386, 159,165) | 3.82 (3.78, 3.87) |
| North Carolina | 79,120 (76,394, 82,029) | 3.44 (3.32, 3.56) | 17,835 (17,577,18,053) | 0.77 (0.76,0.78) | 96,955 (93,971, 100,082) | 4.21 (4.08, 4.35) |
| North Dakota | 3,808 (2,409, 6,887) | 2.05 (1.30, 3.71) | 609 (422,801) | 0.33 (0.23,0.43) | 4,417 (2,831, 7,688) | 2.38 (1.52, 4.14) |
|  | (n, 95% UI) | rate<br>per 100 children<br>(rate, 95% UI) | (n, 95% UI) | rate<br>per 100 children<br>(rate, 95% UI) | (n, 95% UI) | rate<br>per 100 children<br>(rate, 95% UI) |
| Ohio | 97,161 (94,914, 99,413) | 3.73 (3.64, 3.82) | 20,475 (20,261,20,618) | 0.79 (0.78,0.79) | 117,636 (115,175, 120,031) | 4.51 (4.42, 4.61) |
| Oklahoma | 36,240 (32,901, 39,983) | 3.77 (3.42, 4.16) | 8,612 (8,355,8,818) | 0.90 (0.87,0.92) | 44,852 (41,256, 48,801) | 4.66 (4.29, 5.08) |
| Oregon | 22,994 (19,901, 26,282) | 2.67 (2.31, 3.05) | 5,459 (5,192,5,663) | 0.63 (0.60,0.66) | 28,453 (25,093, 31,945) | 3.30 (2.91, 3.71) |
| Pennsylvania | 95,425 (93,303, 97,704) | 3.57 (3.49, 3.65) | 22,181 (21,981,22,279) | 0.83 (0.82,0.83) | 117,606 (115,284, 119,983) | 4.40 (4.31, 4.49) |
| Rhode Island | 4,826 (3,084, 7,091) | 2.31 (1.48, 3.40) | 1,567 (1,295,1,874) | 0.75 (0.62,0.90) | 6,393 (4,379, 8,965) | 3.06 (2.10, 4.29) |
| South Carolina | 39,654 (36,661, 42,890) | 3.55 (3.28, 3.84) | 10,729 (10,470,10,999) | 0.96 (0.94,0.98) | 50,383 (47,131, 53,889) | 4.51 (4.22, 4.82) |
| South Dakota | 5,552 (3,631, 9,040) | 2.52 (1.65, 4.10) | 957 (793,1,154) | 0.43 (0.36,0.52) | 6,509 (4,424, 10,194) | 2.95 (2.01, 4.62) |
| Tennessee | 63,260 (60,342, 66,515) | 4.11 (3.92, 4.32) | 15,134 (14,875,15,388) | 0.98 (0.97,1.00) | 78,394 (75,217, 81,903) | 5.09 (4.88, 5.32) |
| Texas | 224,453 (226,165, 223,024) | 3.00 (3.03, 2.98) | 54,728 (55,197,54,279) | 0.73 (0.74,0.73) | 279,181 (281,362, 277,303) | 3.73 (3.76, 3.71) |
| Utah | 24,529 (20,856, 29,053) | 2.59 (2.20, 3.07) | 4,187 (3,852,4,393) | 0.44 (0.41,0.46) | 28,716 (24,708, 33,446) | 3.03 (2.61, 3.53) |
| Vermont | 2,394 (1,244, 4,255) | 2.05 (1.06, 3.64) | 545 (429,838) | 0.47 (0.37,0.72) | 2,939 (1,673, 5,093) | 2.51 (1.43, 4.35) |
| Virginia | 56,961 (53,825, 60,501) | 3.02 (2.86, 3.21) | 14,070 (13,841,14,335) | 0.75 (0.73,0.76) | 71,031 (67,666, 74,836) | 3.77 (3.59, 3.97) |
| Washington | 42,455 (39,491, 46,039) | 2.53 (2.36, 2.75) | 9,257 (9,043,9,486) | 0.55 (0.54,0.57) | 51,712 (48,534, 55,525) | 3.09 (2.90, 3.31) |
| West Virginia | 17,888 (15,069, 21,094) | 4.98 (4.20, 5.88) | 4,713 (4,394,5,029) | 1.31 (1.22,1.40) | 22,601 (19,463, 26,123) | 6.29 (5.42, 7.28) |
| Wisconsin | 29,802 (26,821, 33,307) | 2.34 (2.10, 2.61) | 5,885 (5,647,6,052) | 0.46 (0.44,0.47) | 35,687 (32,468, 39,359) | 2.80 (2.55, 3.09) |
| Wyoming | 3,021 (1,734, 5,830) | 2.28 (1.31, 4.40) | 605 (472,784) | 0.46 (0.36,0.59) | 3,626 (2,206, 6,614) | 2.74 (1.67, 4.99) |

**Supplementary Table S6:**
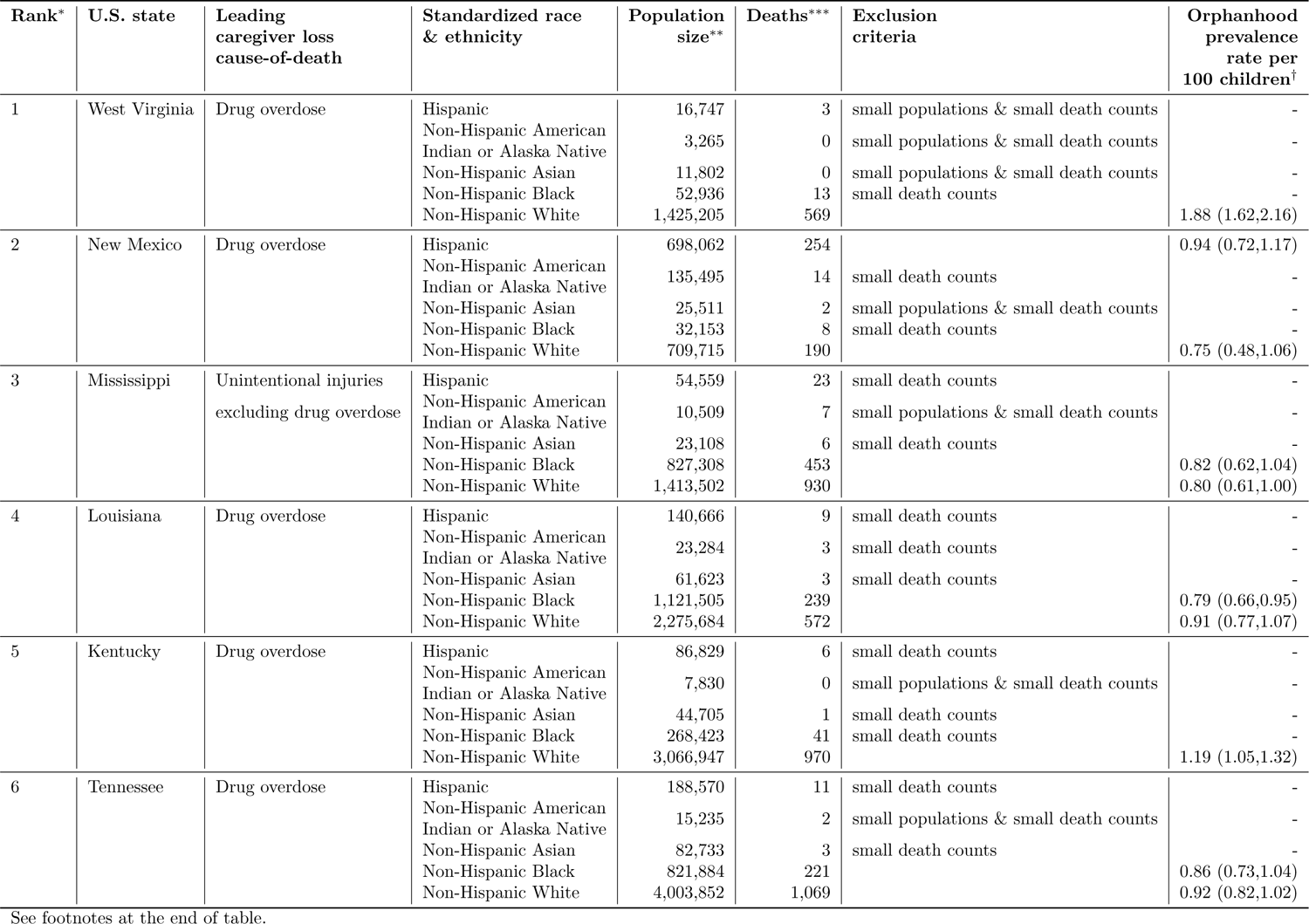
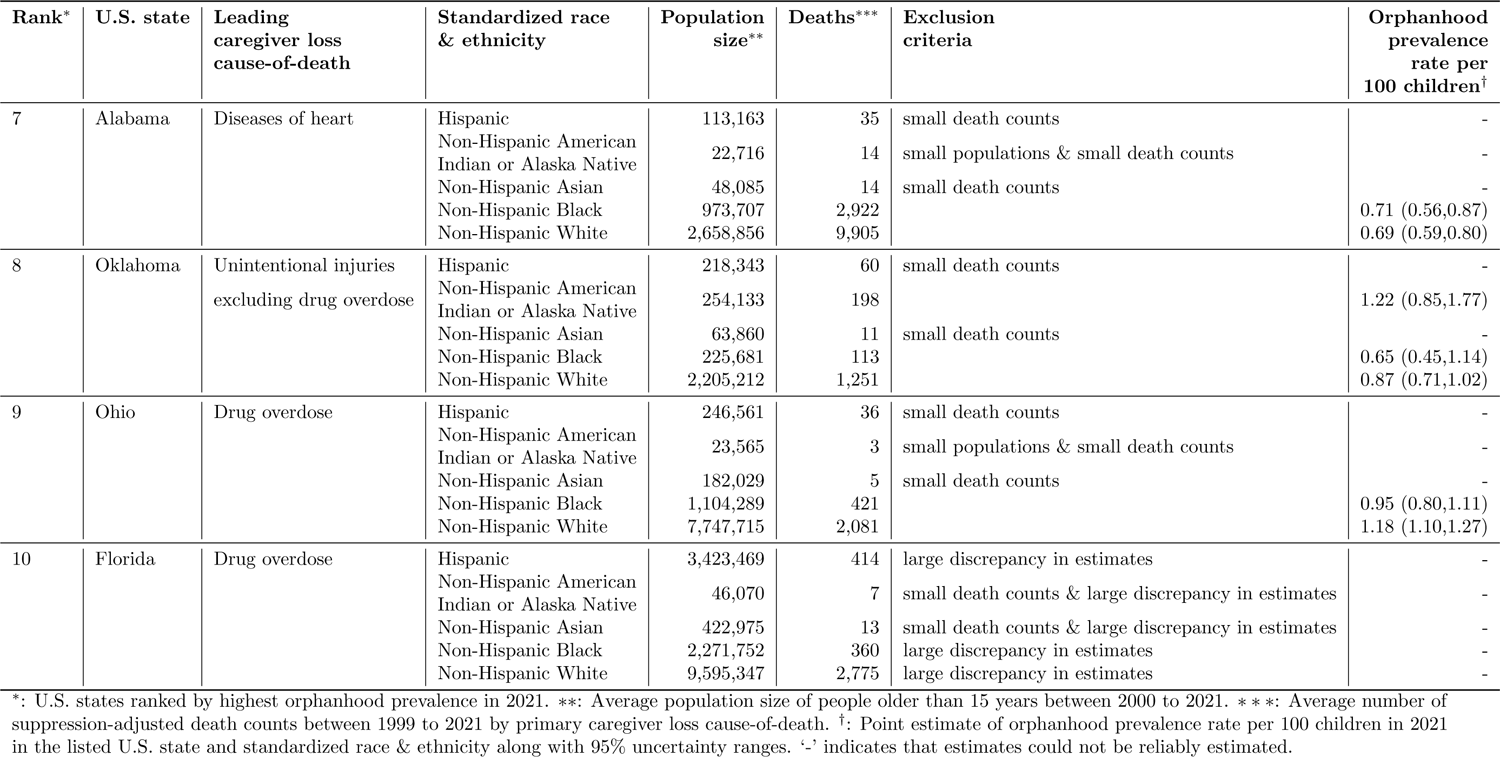
Race & ethnicity groups most affected by the leading caregiver loss cause-of-death in the ten U.S. states with highest orphanhood prevalence rates in 2021.

| Rank* | U.S. state | Leading caregiver loss cause-of-death | Standardized race & ethnicity | Population size** | Deaths*** | Exclusion criteria | Orphanhood prevalence rate per 100 children† |
| --- | --- | --- | --- | --- | --- | --- | --- |
| 1 | West Virginia | Drug overdose | Hispanic | 16,747 | 3 | small populations & small death counts | - |
|  |  |  | Non-Hispanic American Indian or Alaska Native | 3,265 | 0 | small populations & small death counts | - |
|  |  |  | Non-Hispanic Asian | 11,802 | 0 | small populations & small death counts | - |
|  |  |  | Non-Hispanic Black | 52,936 | 13 | small death counts | - |
|  |  |  | Non-Hispanic White | 1,425,205 | 569 |  | 1.88 (1.62,2.16) |
| 2 | New Mexico | Drug overdose | Hispanic | 698,062 | 254 |  | 0.94 (0.72,1.17) |
|  |  |  | Non-Hispanic American Indian or Alaska Native | 135,495 | 14 | small death counts | - |
|  |  |  | Non-Hispanic Asian | 25,511 | 2 | small populations & small death counts | - |
|  |  |  | Non-Hispanic Black | 32,153 | 8 | small death counts | - |
|  |  |  | Non-Hispanic White | 709,715 | 190 |  | 0.75 (0.48,1.06) |
| 3 | Mississippi | Unintentional injuries excluding drug overdose | Hispanic | 54,559 | 23 | small death counts | - |
|  |  |  | Non-Hispanic American Indian or Alaska Native | 10,509 | 7 | small populations & small death counts | - |
|  |  |  | Non-Hispanic Asian | 23,108 | 6 | small death counts | - |
|  |  |  | Non-Hispanic Black | 827,308 | 453 |  | 0.82 (0.62,1.04) |
|  |  |  | Non-Hispanic White | 1,413,502 | 930 |  | 0.80 (0.61,1.00) |
| 4 | Louisiana | Drug overdose | Hispanic | 140,666 | 9 | small death counts | - |
|  |  |  | Non-Hispanic American Indian or Alaska Native | 23,284 | 3 | small death counts | - |
|  |  |  | Non-Hispanic Asian | 61,623 | 3 | small death counts | - |
|  |  |  | Non-Hispanic Black | 1,121,505 | 239 |  | 0.79 (0.66,0.95) |
|  |  |  | Non-Hispanic White | 2,275,684 | 572 |  | 0.91 (0.77,1.07) |
| 5 | Kentucky | Drug overdose | Hispanic | 86,829 | 6 | small death counts | - |
|  |  |  | Non-Hispanic American Indian or Alaska Native | 7,830 | 0 | small populations & small death counts | - |
|  |  |  | Non-Hispanic Asian | 44,705 | 1 | small death counts | - |
|  |  |  | Non-Hispanic Black | 268,423 | 41 | small death counts | - |
|  |  |  | Non-Hispanic White | 3,066,947 | 970 |  | 1.19 (1.05,1.32) |
| 6 | Tennessee | Drug overdose | Hispanic | 188,570 | 11 | small death counts | - |
|  |  |  | Non-Hispanic American Indian or Alaska Native | 15,235 | 2 | small populations & small death counts | - |
|  |  |  | Non-Hispanic Asian | 82,733 | 3 | small death counts | - |
|  |  |  | Non-Hispanic Black | 821,884 | 221 |  | 0.86 (0.73,1.04) |
|  |  |  | Non-Hispanic White | 4,003,852 | 1,069 |  | 0.92 (0.82,1.02) |
| 7 | Alabama | Diseases of heart | Hispanic | 113,163 | 35 | small death counts | - |
|  |  |  | Non-Hispanic American Indian or Alaska Native | 22,716 | 14 | small populations & small death counts | - |
|  |  |  | Non-Hispanic Asian | 48,085 | 14 | small death counts | - |
|  |  |  | Non-Hispanic Black | 973,707 | 2,922 |  | 0.71 (0.56,0.87) |
|  |  |  | Non-Hispanic White | 2,658,856 | 9,905 |  | 0.69 (0.59,0.80) |
| 8 | Oklahoma | Unintentional injuries excluding drug overdose | Hispanic | 218,343 | 60 | small death counts | - |
|  |  |  | Non-Hispanic American Indian or Alaska Native | 254,133 | 198 |  | 1.22 (0.85,1.77) |
|  |  |  | Non-Hispanic Asian | 63,860 | 11 | small death counts | - |
|  |  |  | Non-Hispanic Black | 225,681 | 113 |  | 0.65 (0.45,1.14) |
|  |  |  | Non-Hispanic White | 2,205,212 | 1,251 |  | 0.87 (0.71,1.02) |
| 9 | Ohio | Drug overdose | Hispanic | 246,561 | 36 | small death counts | - |
|  |  |  | Non-Hispanic American Indian or Alaska Native | 23,565 | 3 | small populations & small death counts | - |
|  |  |  | Non-Hispanic Asian | 182,029 | 5 | small death counts | - |
|  |  |  | Non-Hispanic Black | 1,104,289 | 421 |  | 0.95 (0.80,1.11) |
|  |  |  | Non-Hispanic White | 7,747,715 | 2,081 |  | 1.18 (1.10,1.27) |
| 10 | Florida | Drug overdose | Hispanic | 3,423,469 | 414 | large discrepancy in estimates | - |
|  |  |  | Non-Hispanic American Indian or Alaska Native | 46,070 | 7 | small death counts & large discrepancy in estimates | - |
|  |  |  | Non-Hispanic Asian | 422,975 | 13 | small death counts & large discrepancy in estimates | - |
|  |  |  | Non-Hispanic Black | 2,271,752 | 360 | large discrepancy in estimates | - |
|  |  |  | Non-Hispanic White | 9,595,347 | 2,775 | large discrepancy in estimates | - |

\*: U.S. states ranked by highest orphanhood prevalence in 2021. \*\*: Average population size of people older than 15 years between 2000 to 2021. \*\*\*: Average number of suppression-adjusted death counts between 1999 to 2021 by primary caregiver loss cause-of-death. †: Point estimate of orphanhood prevalence rate per 100 children in 2021 in the listed U.S. state and standardized race & ethnicity along with 95% uncertainty ranges. ‘-’ indicates that estimates could not be reliably estimated.

### S1 Extended methods

This Supplementary Text describes the data sources and statistical methods used to estimate cause-specific incidence and prevalence of orphanhood and grandparent caregiver loss among U.S. residents between 2000 and 2021. Our methods are overall similar to Hillis et al. [2021b,a], and extended here to estimate the magnitude of children aged 0-17 years experiencing the death of one or both parents from all causes or the death of a grandparent caregiver from all causes.

The United Nations International Children’s Emergency Fund (UNICEF) defines orphanhood as children experiencing the death of one or both parents [UNAIDS, 2009]. As in Hillis et al. [2021a], we considered mothers of age 15-66 years and fathers of age 15-94 years so the maximum ages of parents at birth of a child were respectively 49 and 77 years. Mortality data were recorded among U.S. residents, and for this reason, orphanhood estimates are restricted to children of U.S. residents. Grandparents play indispensable roles as caregivers for children [United States Census bureau, 2021, Pew Research Center, 2013, US Census Bureau, 2023b, Sowden et al., 2021, Ellis and Simmons, 2014]. We further considered to provide minimum numbers of children who lost a grandparent caregiver, defined as the death of a grandparent caregiver aged older than 30 years living with children in the same household. Mortality data were recorded among U.S. residents, and so grandparent caregiver death estimates are also restricted to children of U.S. resident grandparent caregivers. In Hillis et al. [2021a], we previously stratified grandparent caregivers based on the care they provided into primary grandparent caregivers who were either the guardians of children or co-resided and provided for basic needs of children, as well as secondary grandparent caregivers who co-resided and provided for some needs of children, following the characterizations of Ellis and Simmons [2014]. Here, we report throughout the sum of primary and secondary grandparent caregiver loss (Supplementary Fig. S1).

### S1.1 National-level NCHS mortality data by rankable causes of death, 1983-2021

We obtained line-list mortality data on U.S. residents from the NCHS Vital Statistics portal for each year from 1983 to 2021 [National Center for Health Statistics, 2023]. Data were collected from 1983 onwards because the corresponding children who lost a caregiver in 1983 at age 0 were of age 17 in 2000, and so entered our estimation of orphanhood prevalence in 2000. For each mortality record we retained year of death, the corresponding codes of the underlying cause-of-death (ICD-9 code and 282 cause recode before 1999; ICD-10 code and 113 cause recode after 1999), and demographic data of the decedent including sex, age at death, and information on race and Hispanic origin [National Center for Health Statistics, 2022].

Information about the race and Hispanic origin of decedents in death certificates is typically reported by the surviving next of kin, or on the basis of observation in the absence of an informant [Rosenberg, 1999]. Race and Hispanic origin were reported in different formats across the study period [Ingram et al., 2003], which we mapped to standardized race and Hispanic origin categories as described in Supplementary Table S7. Specifically, we grouped individuals of Hispanic origin and all individuals of non-Hispanic origin by their race, i.e. ‘Hispanic’, ‘Non-Hispanic American Indian or Alaska Native’, ‘Non-Hispanic Asian or Pacific Islander’, ‘Non-Hispanic Black’, ‘Non-Hispanic White’, and we refer to the resulting categories as “standardized race & ethnicity” for simplicity. Data on Hispanic origin were not available for 1983 (see below). Individuals of other races in 1984-1991 and individuals of more than one race in 2021 were not coded consistently and not included in this study. We note this approach to harmonising race reporting over the study period did not necessarily make the primary data fully comparable. Earlier research shows inaccuracies are limited [Heron, 2021], which indicates that the incremental implementation of multiple race reporting in the U.S. is unlikely to have introduced notable bias in our orphanhood estimates.

The underlying cause-of-death is defined by the World Health Organization (WHO) as ‘the disease or injury which initiated the train of events leading directly to death, or the circumstances of the accident or violence which produced the fatal injury’ [World Health Organization, 2016]. For 1983-1998, underlying causes-of-death were in the data classified with the Ninth Revision of the International Classification of Diseases (ICD-9) [World Health Organization, 1977], and grouped further into 282 selected causes of death, termed ‘282 cause recode’ [National Center for Health Statistics, 1979]. For 1999-2021, underlying causes-of-death were in the data classified with the Tenth Revision of the International Classification of Diseases (ICD-10), and grouped further into ‘113 Selected Causes of Death’ [National Center for Health Statistics, 2020].

Due to our interest in orphanhood and caregiver loss, we defined 53 non-overlapping rankable underlying causes-of-death that we termed ‘caregiver loss causes-of-death’ and that apart from small modifications are identical to the 52 rankable underlying causes-of-death from the NCHS 113 Selected Causes of Death list [National Center for Health Statistics, 2020, Anderson et al., 2001]. The modifications are listed in Supplementary Table S8. Specifically, we re-categorized drug-induced causes-of-death into ‘Drug overdose’, joining the ICD-9/ICD-10 causes-of-death ‘Intentional self-poisoning by and exposure to nonopioid analgesics, antipyretics and antirheumatics’, ‘Intentional self-poisoning by and exposure to antiepileptic, sedative-hypnotic, antiparkinsonism and psychotropic drugs, not elsewhere classified’, ‘Intentional self-poisoning by and exposure to narcotics and psychodysleptics [hallucinogens], not elsewhere classified’, ‘Intentional self-poisoning by and exposure to other drugs acting on the autonomic nervous system’, ‘Intentional self-poisoning by and exposure to other and unspecified drugs, medicaments and biological substances’ (E950.0-E950.5; X60-X64); ‘Assault by drugs, medicaments and biological substances’ (E962.0; X85); ‘Accidental poisoning by and exposure to nonopioid analgesics, antipyretics and antirheumatics’, ‘Accidental poisoning by and exposure to antiepileptic, sedative-hypnotic, antiparkinsonism and psychotropic drugs, not elsewhere classified’, ‘Accidental poisoning by and exposure to narcotics and psychodysleptics [hallucinogens], not elsewhere classified’, ‘Accidental poisoning by and exposure to other drugs acting on the autonomic nervous system’, ‘Accidental poisoning by and exposure to other and unspecified drugs, medicaments and biological substances’ (E850-E858; X40-X44); and ‘Poisoning by and exposure to nonopioid analgesics, antipyretics and antirheumatics, undetermined intent’, ‘Poisoning by and exposure to antiepileptic, sedative-hypnotic, antiparkinsonism and psychotropic drugs, not elsewhere classified, undetermined intent’, ‘Poisoning by and exposure to narcotics and psychodysleptics [hallucinogens], not elsewhere classified, undetermined intent’, ‘Poisoning by and exposure to other drugs acting on the autonomic nervous system, undetermined intent’, ‘Poisoning by and exposure to other and unspecified drugs, medicaments and biological substances, undetermined intent’ (E980.0-E980.5; Y10-Y14).

We retained these four drug overdose causes-of-death sub-categories as separate drug overdose sub-groups for the purpose of data harmonization (see below). Correspondingly, we removed the related three drug overdose causes-of-death sub-categories respectively from ‘Intentional self-harm’, ‘Assault’ and ‘Accidents’. We renamed these resulting three categories as ‘Suicide excluding drug overdose’, ‘Homicide excluding drug overdose’ and ‘Unintentional injuries excluding drug overdose’ (see Supplementary Table S8). Finally, to simplify presentation as in Figure 1 in the main text, we primarily considered COVID-19 and the leading five causes-of-death between 1983 and 2021 due to our interest in all caregiver loss. Additionally, we included any cause in the top five for adult ages 15-44 years, as these ages specifically include younger adults who are more likely to be parents of younger children. The only one addition to our causes when taking this approach was ‘Homicide excluding drug overdose’. Supplementary Table S9 summarises our aggregation of the 53 rankable causes-of-death into these seven leading caregiver loss cause-of-death groups and ‘Other’ caregiver loss causes-of-death. The ‘Other’ caregiver loss causes-of-death comprise the remaining 46 rankable caregiver loss causes-of-death and any other causes-of-death that are not included in the NCHS 52 rankable causes-of-death.

We then mapped and aggregated line-list mortality records to the 53 rankable caregiver loss causes-of-death. This was done by mapping the 1983-1998 line list data using the 282 recodes to the 52 NCHS rankable causes-of-death as described in [National Center for Health Statistics, 1988] on page 282ff and page 290ff. For drug-induced causes, we used Table 2 in [National Center for Injury Prevention and Control (U.S.). Division of Unintentional Injury Prevention. Health Systems and Trauma Systems Branch. Prescription Drug Overdose Team, 2013]. Supplementary Table S9 summarises our approach.

For 1999-2021, we mapped ICD-10 113 cause recodes to NCHS 52 rankable causes-of-death based on Table A in [Curtin SC, 2023], and subsequently mapped the NCHS 52 rankable causes-of-death to the 53 rankable caregiver loss causes-of-death based on Supplementary Table S9.

Next, we aggregated line-list death records to annualised death counts by the 53 rankable caregiver loss causes-of-death for each year in 1983-2021, as well as by sex, age band (15-19, 20-24, 25-29, 30-34, 35-39, 40-44, 45-49, 50-54, 55-59, 60-64, 65-69, 70-74, 75-79, 80-84 and 85+ years) and standardized race & ethnicity of the decedent. Data on race and ethnicity were not available for 1983, and for this year we attributed mortality counts to standardized race categories according to the age-, sex- and cause-of-death-specific standardized race compositions in 1984. To harmonise cause-of-death data from 1983-1999 to the ICD-10 cause-of-death classifications and avoid discontinuities in our orphanhood estimates from 1998 to 1999, we used where available the comparability ratios in Table 1 of [Anderson et al., 2001]. Thirteen comparability ratios of rankable causes of death were not provided in [Anderson et al., 2001] when underlying estimations were considered imprecise, and in these cases we set the comparability ratios to 1. Supplementary Fig. S4 illustrates the aggregated annual mortality data among U.S. residents by standardized race categories, and Supplementary Fig. S5 by leading causes-of-death as relevant for caregiver loss and orphanhood.

**Supplementary Table S7:** Mapping of single-race mortality data to standardized race categories and Hispanic origin.

| Standardized race categories | Time period (years) |  |  |  |
| --- | --- | --- | --- | --- |
|  | 1984-1991 | 1992-2003 | 2004-2020 | 2021 |
| American Indian or Alaska Native | American Indian (includes Aleuts and Eskimos) | American Indian (includes Aleuts and Eskimos) | American Indian | American Indian or Alaskan Native |
| Asian or Pacific Islander | Chinese; Japanese; Filipino; Hawaiian (includes Part-Hawaiian); Other Asian or Pacific Islander | Chinese; Japanese; Filipino; Hawaiian (includes Part-Hawaiian); Asian Indian; Korean; Samoan; Vietnamese; Guamanian; Other Asian or Pacific Islander in areas; Combined other Asian or Pacific Islander | Asian or Pacific Islander | Chinese; Japanese; Filipino; Hawaiian; Asian Indian; Korean; Samoan; Vietnamese; Guamanian; Other or Multiple Asian; Other or Multiple Pacific Islander |
| Black or African American | Black | Black | Black | Black |
| White | White | White | White | White |
| Not included in study<sup>†</sup> | Other races | - | - | More than one race |
| **Ethnicity** |  |  |  |  |
| Hispanic origin | Spaniard; Mexican; Puerto Rican; Cuban; Dominican; Central or South American; Central American; South American; Latin American; Other and Unknown Spanish; Other Hispanic |  |  |  |

<sup>†</sup>: Individuals of other races in 1984-1991 and individuals of more than one race in 2021 were not coded consistently and not included in this study, which corresponded to 0.0068% of all death records in 1983-1991 and 0.47% in 2021.

**Supplementary Table S8:** Definitions of caregiver loss causes-of-death used in this analysis relative to the 52 rankable causes-of-death included in the NCHS 113 Selected Causes of Death.

| ID | Caregiver loss<br>causes-of-death | NCHS 52 rankable<br>underlying causes-of-death<sup>†</sup> | Further modifications<sup>‡</sup> |  |
| --- | --- | --- | --- | --- |
|  |  |  | ICD-9 | ICD-10 |
| 1 | Acute bronchitis and bronchiolitis | Acute bronchitis and bronchiolitis | - | - |
| 2 | Acute poliomyelitis | Acute poliomyelitis | - | - |
| 3 | Alzheimer disease | Alzheimer disease | - | - |
| 4 | Anemias | Anemias | - | - |
| 5 | Aortic aneurysm and dissection | Aortic aneurysm and dissection | - | - |
| 6 | Arthropod-borne viral encephalitis | Arthropod-borne viral encephalitis | - | - |
| 7 | Atherosclerosis | Atherosclerosis | - | - |
| 8 | Cerebrovascular diseases | Cerebrovascular diseases | - | - |
| 9 | Certain conditions originating in the<br>perinatal period | Certain conditions originating in the<br>perinatal period | - | - |
| 10 | Cholelithiasis and other disorders of gallbladder | Cholelithiasis and other disorders of gallbladder | - | - |
| 11 | Chronic liver disease and cirrhosis | Chronic liver disease and cirrhosis | - | - |
| 12 | Chronic lower respiratory diseases | Chronic lower respiratory diseases | - | - |
| 13 | Complications of medical and surgical care | Complications of medical and surgical care | - | - |
| 14 | Congenital malformations, deformations<br>and chromosomal abnormalities | Congenital malformations, deformations<br>and chromosomal abnormalities | - | - |
| 15 | COVID-19 | COVID-19 | - | - |
| 16 | Diabetes mellitus | Diabetes mellitus | - | - |
| 17 | Diseases of appendix | Diseases of appendix | - | - |
| 18 | Diseases of heart | Diseases of heart | - | - |
| 19 | Drug overdose |  | Adding: | Adding: |
|  | ... Assault by drugs |  | E962.0 | X85 |
|  | ... Intentional self-poisoning |  | E950.0-E950.5 | X60-X64 |
|  | ... Poisoning with undetermined intent |  | E980.0-E980.5 | Y10-Y14 |
|  | ... Unintentional self-poisoning |  | E850-E858 | X40-X44 |
| 20 | Enterocolitis due to Clostridium difficile | Enterocolitis due to Clostridium difficile | - | - |
| 21 | Essential hypertension and hypertensive<br>renal disease | Essential hypertension and hypertensive<br>renal disease | - | - |
| 22 | Hernia | Hernia | - | - |
| 23 | Homicide, excluding drug overdose | Assault | Removing E962.0 from<br>(E960-E969) | Removing X85 from<br>(*U01 - *U02,X85-Y09,Y87.1) |
| 24 | Human immunodeficiency virus | Human immunodeficiency virus | - | - |
| 25 | Hyperplasia of prostate | Hyperplasia of prostate | - | - |
| 26 | In situ neoplasms, benign neoplasms<br>and neoplasms of uncertain or unknown behavior | In situ neoplasms, benign neoplasms<br>and neoplasms of uncertain or unknown behavior | - | - |
| 27 | Infections of kidney | Infections of kidney | - | - |

| ID | Caregiver loss<br>causes-of-death | NCHS 52 rankable<br>underlying causes-of-death<sup>†</sup> | ICD-9 | Further modifications<sup>‡</sup><br>ICD-10 |
| --- | --- | --- | --- | --- |
| 28 | Inflammatory diseases of female pelvic organs | Inflammatory diseases of female pelvic organs | - | - |
| 29 | Influenza and pneumonia | Influenza and pneumonia | - | - |
| 30 | Legal intervention | Legal intervention | - | - |
| 31 | Malaria | Malaria | - | - |
| 32 | Malignant neoplasms | Malignant neoplasms | - | - |
| 33 | Measles | Measles | - | - |
| 34 | Meningitis | Meningitis | - | - |
| 35 | Meningococcal infection | Meningococcal infection | - | - |
| 36 | Nephritis, nephrotic syndrome and nephrosis | Nephritis, nephrotic syndrome and nephrosis | - | - |
| 37 | Nutritional deficiencies | Nutritional deficiencies | - | - |
| 38 | Operations of war and their sequelae | Operations of war and their sequelae | - | - |
| 39 | Parkinson disease | Parkinson disease | - | - |
| 40 | Peptic ulcer | Peptic ulcer | - | - |
| 41 | Pneumoconioses and chemical effects | Pneumoconioses and chemical effects | - | - |
| 42 | Pneumonitis due to solids and liquids | Pneumonitis due to solids and liquids | - | - |
| 43 | Pregnancy, childbirth and the puerperium | Pregnancy, childbirth and the puerperium | - | - |
| 44 | Salmonella infections | Salmonella infections | - | - |
| 45 | Scarlet fever and erysipelas | Scarlet fever and erysipelas | - | - |
| 46 | Septicemia | Septicemia | - | - |
| 47 | Shigellosis and amebiasis | Shigellosis and amebiasis | - | - |
| 48 | Suicide, excluding drug overdose | Intentional self-harm | Removing E950.0-E950.5 from E950-E959 | Removing X60-X64 from (*U03, X60-X84, Y87.0) |
| 49 | Syphilis | Syphilis | - | - |
| 50 | Tuberculosis | Tuberculosis | - | - |
| 51 | Unintentional injuries, excluding drug overdose | Accidents | Removing E850-E858 from (E800-E869, E880-E929) | Removing X40-X44 from (V01-X59, Y85-Y86) |
| 52 | Viral hepatitis | Viral hepatitis | - | - |
| 53 | Whooping cough | Whooping cough | - | - |
|  | Other | Other | - | - |

<sup>†</sup>: The subset of the NCHS 113 Selected cause list with the reported cause names. Empty cells represent that the corresponding causes of death are not in the NCHS 113 Selected cause list. <sup>‡</sup>: '-' denotes there were no modifications from the NCHS 52 rankable causes-of-death to caregiver loss causes-of-death.

**Supplementary Fig. S4:**
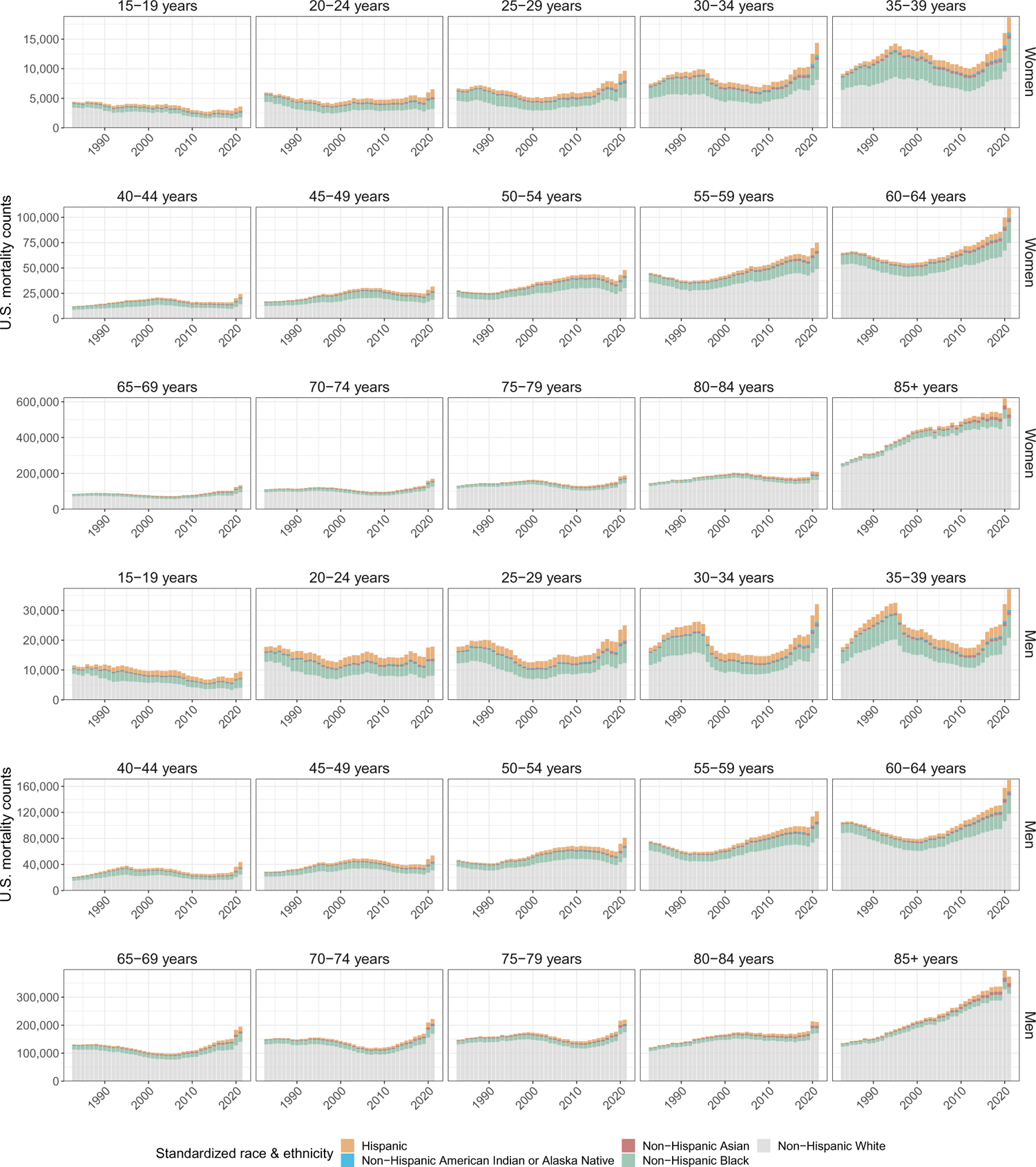
Aggregated age- and sex-specific U.S. mortality counts in 1983-2021, by standardized race & ethnicity.

**Supplementary Fig. S5:**
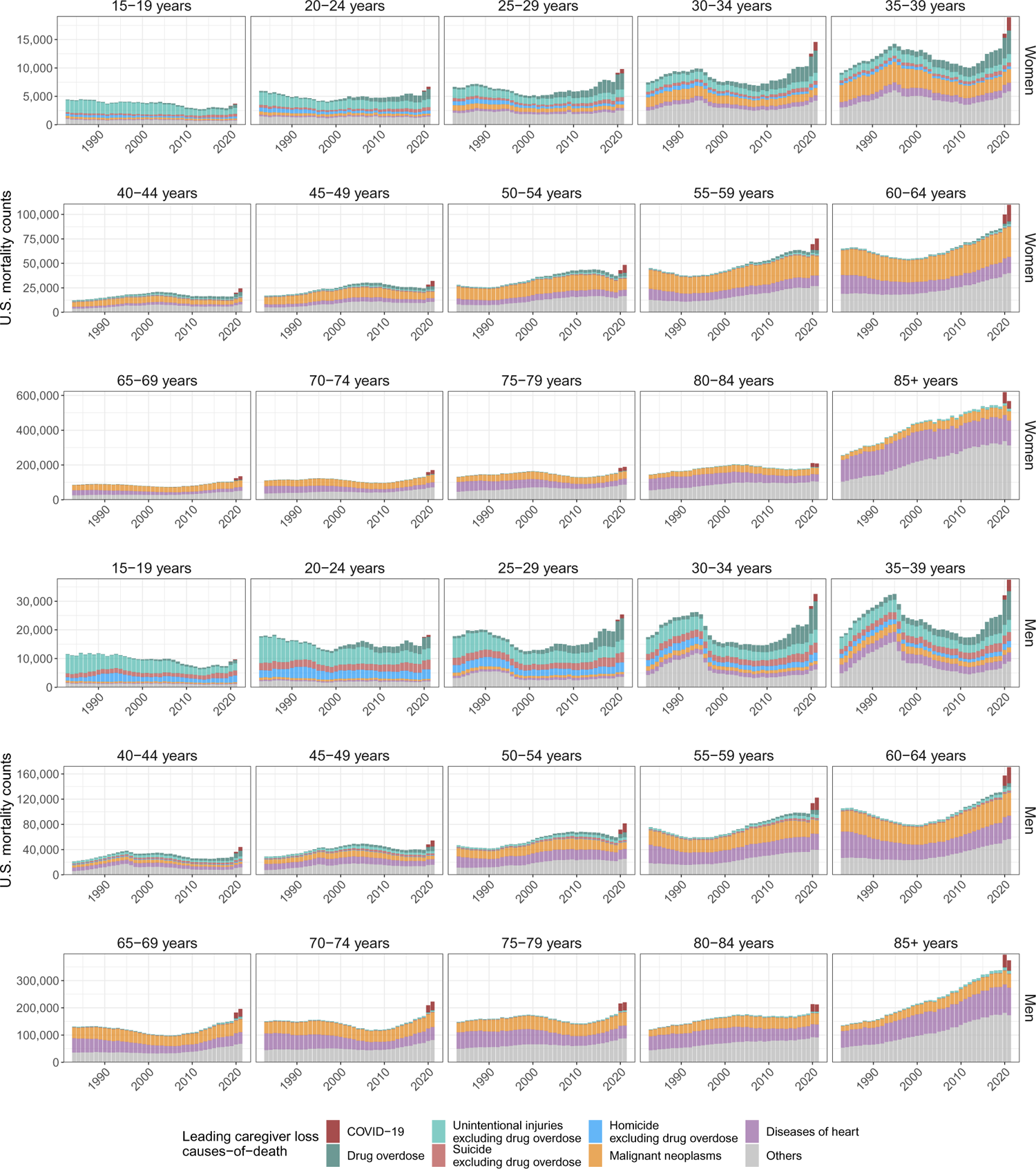
Aggregated age- and sex-specific U.S. mortality counts in 1983-2021, by leading causes-of-death as relevant for caregiver loss and orphanhood.

### S1.2 National-level live births data, 1969-2021

To calculate fertility rates and attribute children experiencing orphanhood to decendents between 1983 to 2021, we required for our approach to estimating orphanhood live births data from before 1983 because the children who lost a caregiver in 1983 at age 1-17 years were born between 1966-1982. However due to limitations in publicly available population size data, we only considered in the central analysis live births data from 1990 and assumed constant fertility rates before 1990. We investigated the sensitivity of our orphanhood estimates to this assumption using live birth records since 1980 together with corresponding population size estimates, and found that orphanhood prevalence estimates since 2008 were not affected by our assumptions on historic fertility, while in the sensitivity analysis orphanhood prevalence estimates for 2000-2007 were slightly lower due to overall lower fertility rates in the 1980s (Supplementary Text S2). Line-list live births with demographic data on both mothers and fathers were available and downloaded from the NCHS Vital Statistics portal for each year between 1969 to 2021 [National Center for Health Statistics, 2023]. We considered only live birth records to U.S. resident mothers for consistency with the mortality data available. Information on residency status of fathers was not available, and we assumed fathers were also U.S. residents in the retained birth records associated with U.S. resident mothers. For each live birth, we retained year of birth, the age of mothers and fathers, and information on race and Hispanic ethnicity [National Center for Health Statistics, 2023].

Age of mothers and fathers was reported by single year of age. Following Hillis et al. [2021a], we considered live births to women aged 15-49 years and men aged 15-77 years, to match the mortality data of women aged 15-66 years and men aged 15-94 years. In total, less than 0.01% of line-list records were removed from further analysis because parent were outside of these age ranges. Age information was mapped to 5-year age bands 15-19 years, *…*, 45-49 years for mothers and age bands 15-19 years, *…*, 50-54 years, and 55 years and older (55+ years) for fathers.

Information about the race and Hispanic origin of mothers and fathers was between 1969-2021 reported in different standards due to the revision of the U.S. certificates of live births on race in 1989 and 2003 [National Center for Health Statistics, 2006]. Information on the race of mothers and fathers is publicly available since 1969, and information on their ethnicity since 1978. For 1978-2021, we mapped available information on race and ethnicity to the same standardized race & ethnicity categories used to stratify the mortality data, according to the mapping described in Supplementary Table S10.

Next, we aggregated line-list live birth records to annualized live birth counts to mothers (in age bands 15-19, *…*, 45-49 years) and fathers (in age bands 15-19, *…*, 55+ years) for each year in 1969-1977, and stratified further by standardized race & ethnicity categories for each year in 1978-2021. Before 1985, data reporting was incomplete for some U.S. states and we used NCHS sampling weights reported in each year (see Appendix A of National Center for Health Statistics [1978]) to extrapolate reported live birth counts to state populations. Supplementary Fig. S6 illustrates the aggregated annual live birth counts by mothers and fathers and standardized race & ethnicity categories since 1978.

**Supplementary Fig. S6:**
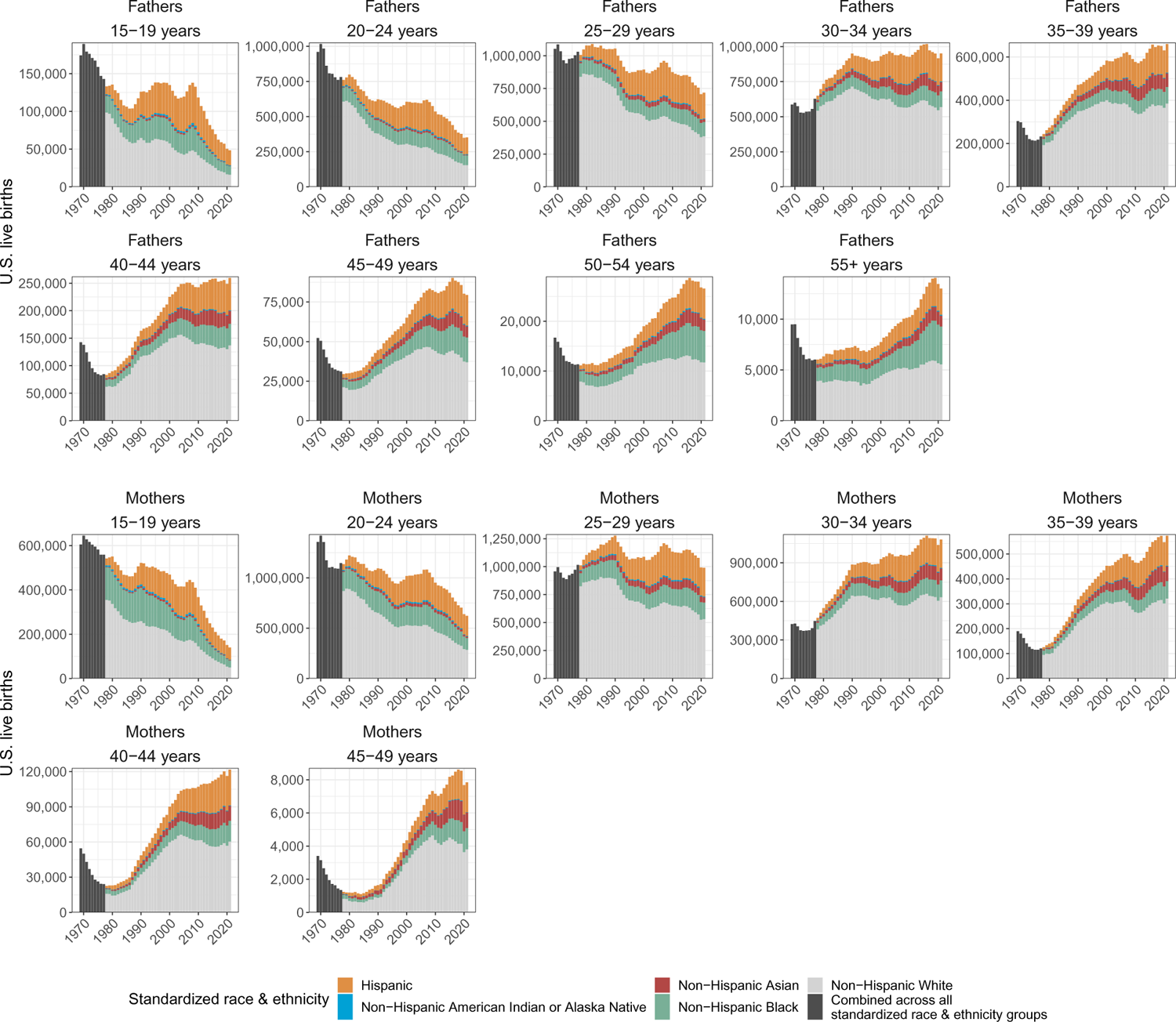
Aggregated age-specific live births counts in 1969-2021, by standardized race & ethnicity categories. Prior to 1978, information on ethnicity was not publicly available and live birth counts are shown without standardized race & ethnicity stratification.

**Supplementary Table S9:**
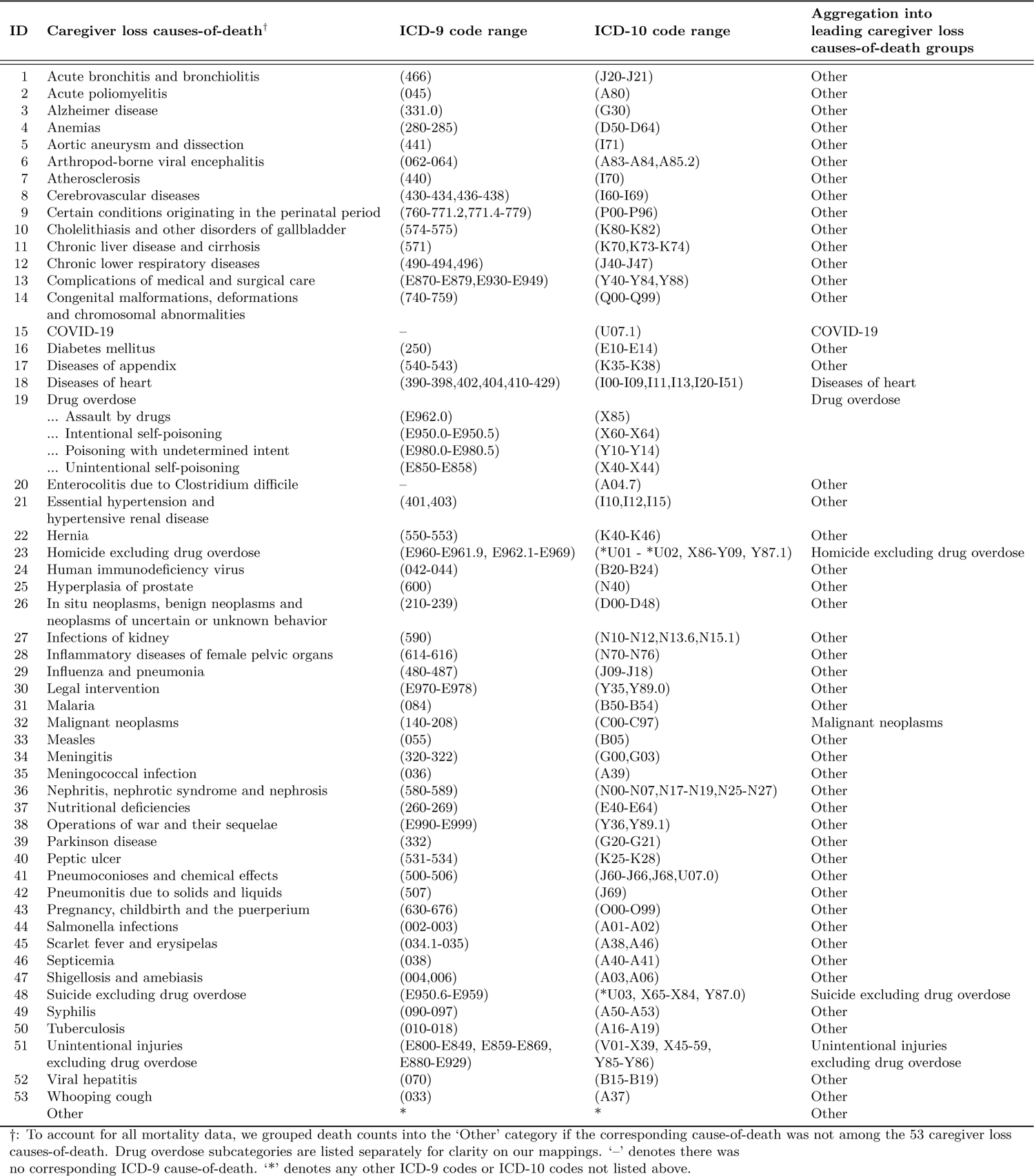
Aggregation of 53 rankable caregiver loss causes-of-death into leading caregiver loss causes-of-death groups.

**Supplementary Table S10:**
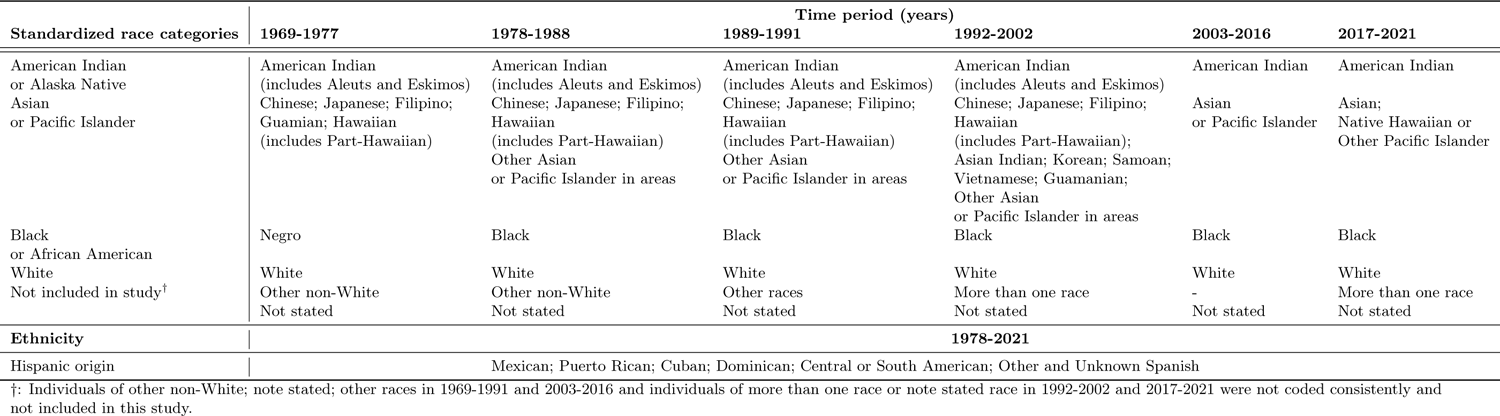
Mapping of single-race live birth data to standardized race categories and Hispanic origin.

### S1.3 National-level population size data, 1990-2021

To calculate fertility rates, we further required population size data of U.S. residents in the age-, sex- and standardized race & ethnicity categories of the live births data. We obtained from the CDC WONDER Vintage bridged-race postcensal & ethnicity population size estimates for each year in 1990-1999 and 2000-2020 [CDC, 2021a], and single race population size estimates for 2021 [CDC, 2021b]. Data were extracted by 5-year age bands (15-19 years, *…*, 70-74 years) and single years of age from 75 to 77 years, and data for individuals aged 55-77 years were summed into a single age band as required for the purposes of our analyses. Race-specific population size estimates were aggregated to the standardized race & ethnicity categories in Supplementary Table S11.

Bridged-race & ethnicity population size data were not publicly available prior to 1990. For sensitivity analyses, we also obtained national-level population size estimates by 5-year age bands, sex, and U.S. states without race & ethnicity stratification for each year in 1969-1989 from the U.S. National Cancer Institute Surveillance, Epidemiology, and End Results (SEER) Program [National Cancer Institute, 2023].

**Supplementary Table S11:**
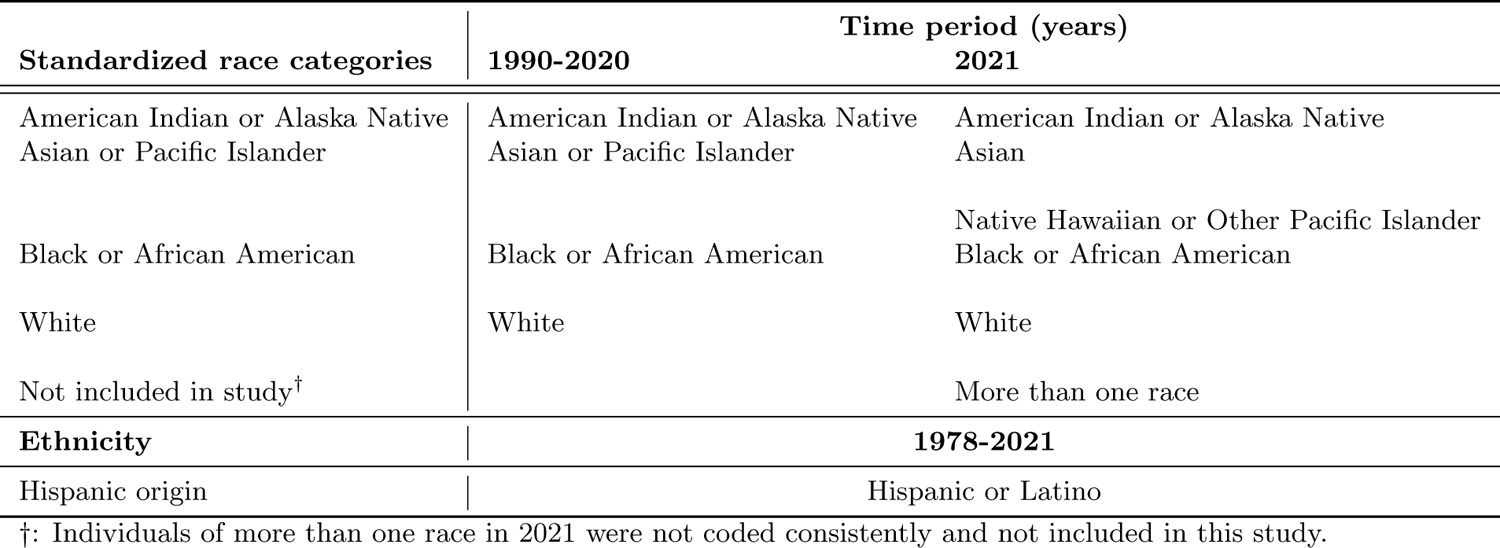
Mapping of single-race population size data to standardized race categories and Hispanic origin.

### S1.4 Estimates of national-level fertility rates, 1990-2021

We calculated age-, sex- and standardized race & ethnicity-specific fertility rates in each year in 1990-2021 for women in one of the age bands *a ∈ {*15 *−* 19*, …,* 45 *−* 49*}* years and men in one of the age bands *a ∈ {*15 *−* 19,, 55 *−* 77*}* years according to

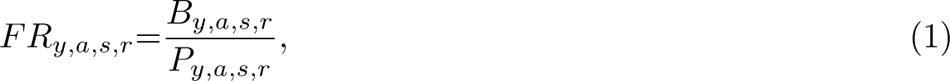

where the number of live births and population sizes in each strata are denoted by *B_y,a,s,r_*and *P_y,a,s,r_* respectively. Calculations were done for each of the five standardized race categories ‘Hispanic’, ‘Non-Hispanic American Indian or Alaska Native’, ‘Non-Hispanic Asian or Pacific Islander’, ‘Non-Hispanic Black’ and ‘Non-Hispanic White’. In the central analysis, we assumed the same fertility rates in each year in 1966-1989 as in 1990, and considered alternative assumptions in several sensitivity analyses (see below). Supplementary Fig. S7 shows the calculated U.S. fertility rates by standardized race & ethnicity.

Mortality rates were calculated in analogy to Eq. (1). We found correlations by standardized race & ethnicity between fertility and mortality rates that changed primarily by age of mothers and fathers, and less so over calendar years (Supplementary Fig. S8 for women); findings were similar in men. These correlations prompted us to estimate national-level incidence and prevalence of orphanhood by standardized race & ethnicity and then sum the standardized race & ethnicity-specific estimates to obtain national-level estimates.

**Supplementary Fig. S7:**
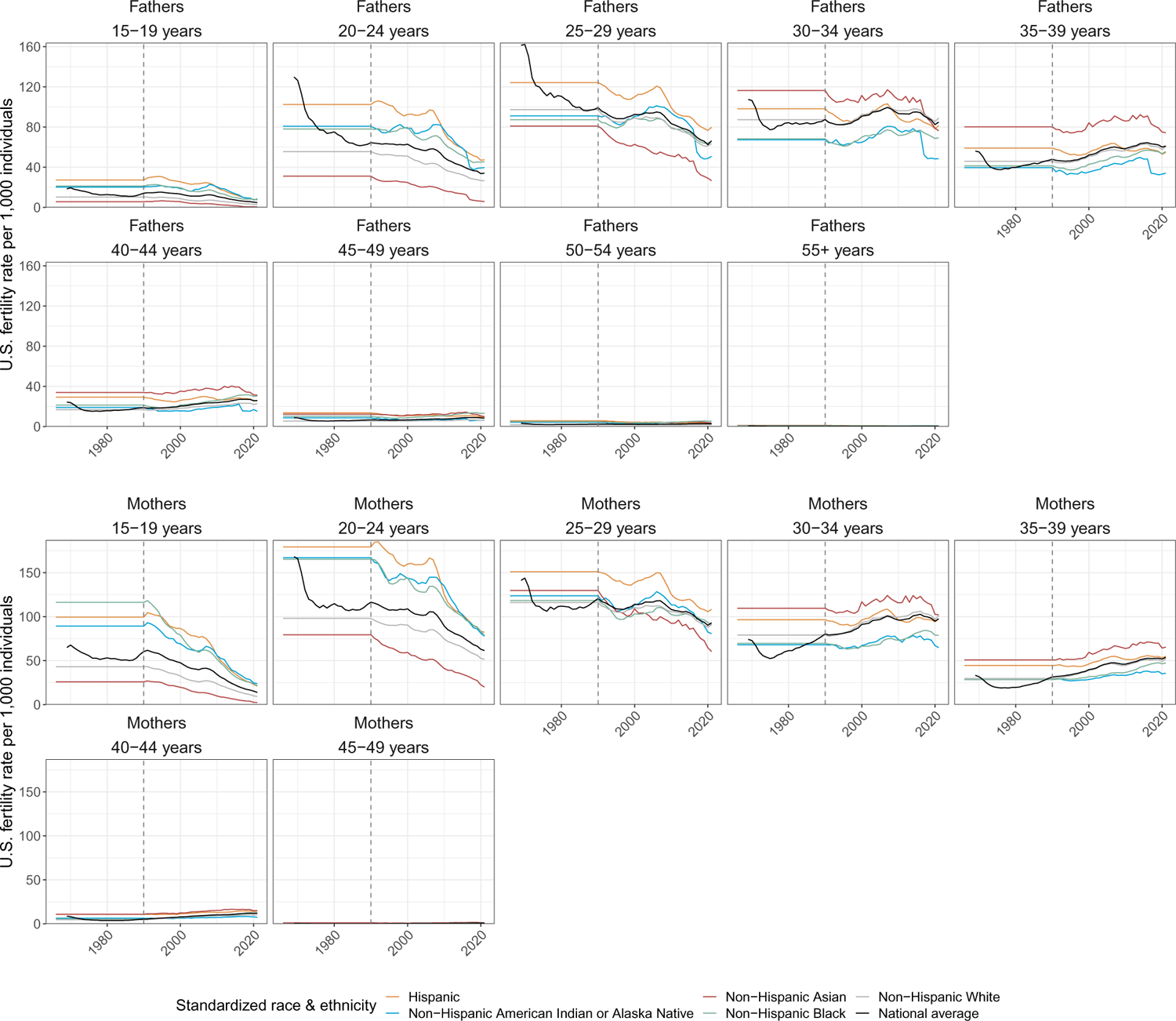
National-level fertility rates per 1,000 population by standardized race & ethnicity in 1990-2021, and assuming constant rates in 1966-1989. For comparison, the black lines show national-level fertility rates, which we calculated directly in 1969-1989 and calculated as weighted average of the standardized race & ethnicity-specific fertility rates in 1990-2021. The dashed vertical line represents 1990, where before 1990 we assumed the same fertility rates in each year in 1966-1989 as in 1990.

**Supplementary Fig. S8:**
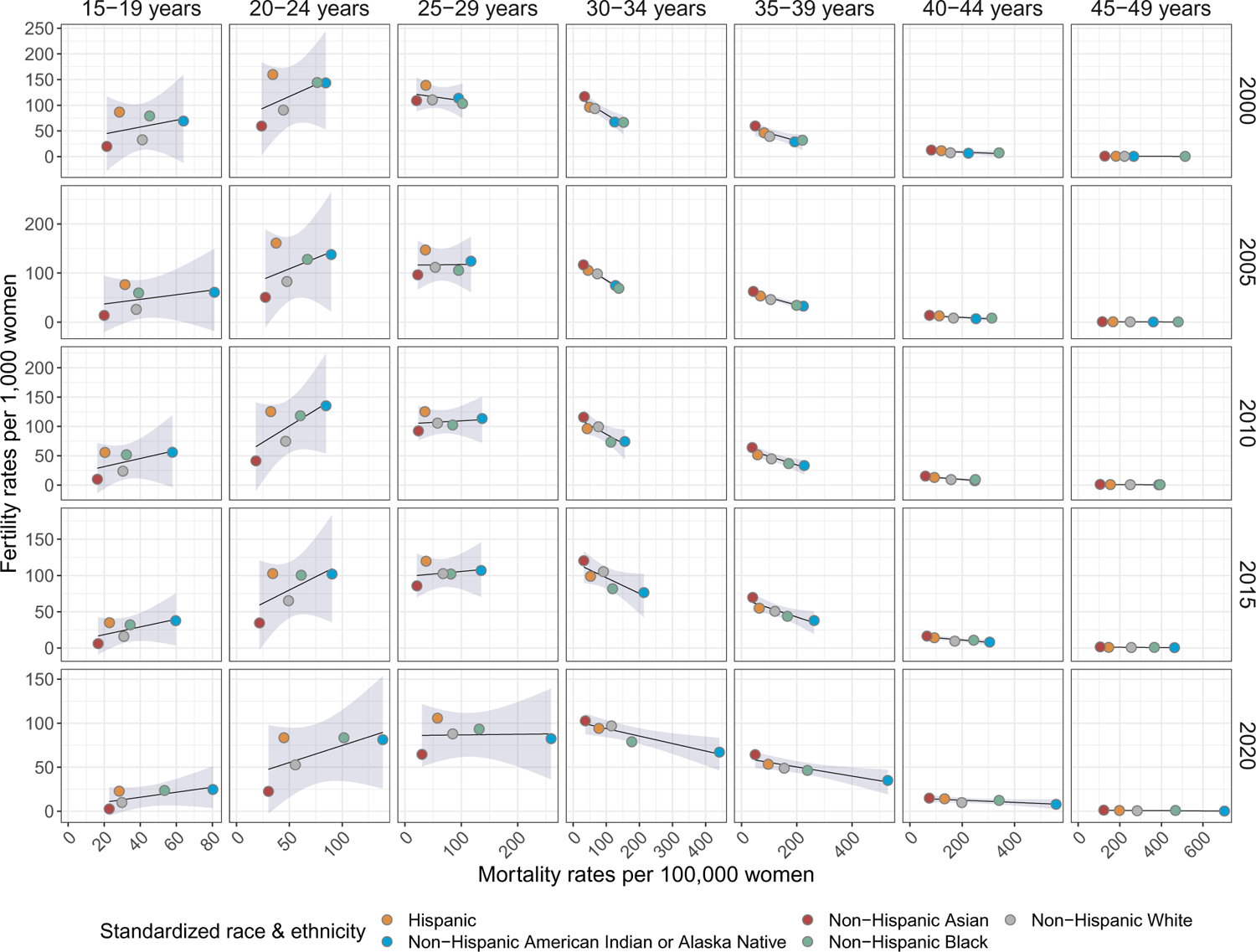
Correlations between standardized race & ethnicity-specific female fertility rates and standardized race & ethnicity-specific female mortality rates by age of mothers and calendar year. Empirical fertility rates are shown as dots, and linear regression fits with 95% prediction intervals are added to visualize correlations.

### S1.5 Estimates of national-level orphanhood, 1983-2021

We estimated the number of children who newly experienced orphanhood in year *y*, *y* = 1983*, …,* 2021, from the population-level mortality records of U.S. residents in year *y* and the number of children each decedent was expected to leave behind. Our approach follows the methods described in [Hillis et al., 2021a, Unwin et al., 2022], with modifications to account for causes-of-death.

We obtained the expected number of children per one U.S. resident of age *a* years, sex *s*, and standardized race & ethnicity *r* in year *y* who are of age *b* = 0, 1*, …,* 17 years (denoted with *C_y,a,g,r,b_*) by multiplying the corresponding fertility rates of Eq. (1) with pediatric survival probabilities of children born in year *y − b* and surviving until age *b* + 1 (denoted with *p*^survive^). Specifically,

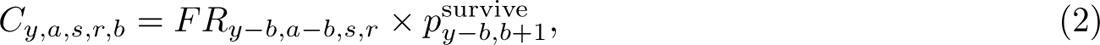

where *y* ranges from 1983 to 2021, the single year of age of mothers ranges from *a ∈ {*15*, …,* 66*}* years, the single year of age of fathers ranges from *a ∈ {*15*, …,* 94*}* years, the standardized race & ethnicity categories are as described before, and *b* = 0, 1*, …,* 17 years. We obtained the pediatric survival probabilities from the child mortality data in [United Nations Population Division, 2020] and use in Eq. (2) the fertility rates of the age band that includes age *a − b*. We then estimated the number of children aged *b* who newly experienced in year *y* the death of a parent *s* of age specified in 5-year age bands *a^′^ ∈ A* = *{*15 *−* 19*, · · ·,* 80 *−* 84, 85 +years*}*, and standardized race & ethnicity *r* who died of caregiver loss cause-of-death *c* by

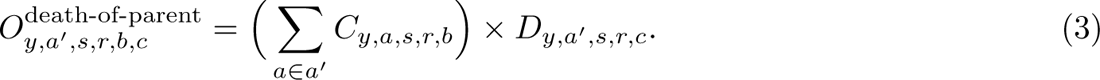

Due to the correlations between standardized race & ethnicity specific fertility and mortality rates, we calculated Eq. (3) for each standardized race & ethnicity. In Eq. (3), we assumed that population-level fertility rates are not correlated with population-level mortality rates, which may lead to upwards or downwards bias in orphanhood estimates that we explored in several sensitivity analyses (see below). We also assumed that parents and their children have the same standardized race & ethnicity.

Since orphanhood considers children who experienced the death of their mother, father or both, we are interested in the sum of Eq. (3) for both mothers and fathers, but need to subtract children who lost their other parent in the previous *b −* 1 years, or who lost their other parent in the same year *y*. Throughout, we assumed that the other parent 1 *− s* is in the same age band *a^′^*and of the same standardized race & ethnicity as parent *s*. The probability that the other biological parent of the children in Eq. (3) died in the same year *y* is based on standard life table calculations [Shryock et al., 1975, Lee and Wang, 2003, Arias et al., 2022], specifically the probability that an individual of sex 1 *− s* and standardized race & ethnicity *r* died in year *y* between (continuous) age *a* and *a* + *n* conditional on survival up to age *a*, where *n* = 5 corresponds to the width of the 5-year age bands considered. This mortality hazard is approximated using midpoints *x* = (*a* + *a* + 5)*/*2 in each age interval, through

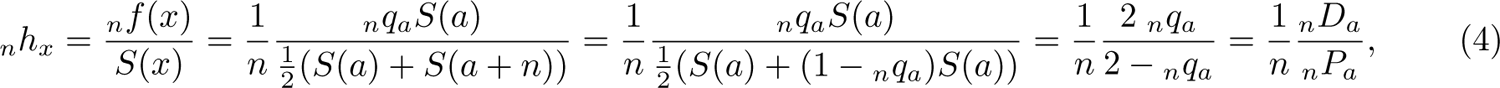

where for ease of readability we have suppressed *y*, *s* and *r*, and *_n_P_a_* and *_n_D_a_*are respectively the estimated population sizes and observed death counts by the end of the corresponding calendar year in each 5-year age band *a^′^* = [*a, a* + 5). The intermediate quantities in (4) are the approximated (unknown) mortality probability density function *_n_f* (*x*) at age midpoint *x* and (unknown) survival function *S*(*a*) up to age *a*, that can be expressed in terms of the age-specific mortality rate *_n_q_a_*, the proportion of individuals alive at age *a* and who die before reaching age *a* + *n* in the corresponding calendar year. It is standard to estimate *_n_q_a_* with (*_n_D_a_*)*/*(*_n_P_a_* + ^1^ *_n_D_a_*), from which Eq. (4) follows [Shryock et al., 1975, Lee and Wang, 2003, Arias et al., 2022]. Using Eq. (4), we estimated the number of children aged *b* who newly experienced in year *y* the death of one parent due to cause-of-death *c* and the death of the other parent due to any cause with

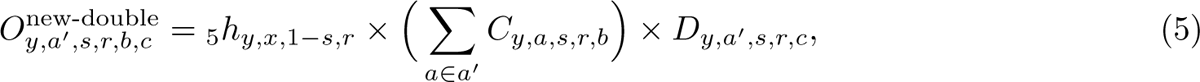

where *x* is the midpoint age in age band *a^′^*. In Eq. (5), we assumed that deaths among parents occurred independently of each other and ignored correlations of deaths among parents by the same cause-of-death such as COVID-19 [Hillis et al., 2021a], as well as correlations of deaths among parents who died of different causes-of-death. Similarly, we estimated that the number of children aged *b* who newly experienced in year *y* the death of one parent *s* due to cause-of-death *c* and the death of their other parent 1 *− s* due to any cause in any of the previous *i* = 1*, …, b −* 1 years with

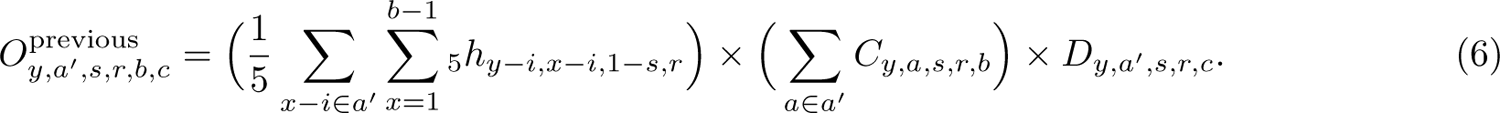

With these considerations, we estimated the number of children aged *b* who newly experienced orphanhood in year *y* = 1983*, …,* 2021, by the death of one or both parents of age *a^′^* and standardized race & ethnicity *r* who died of cause-of-death *c* with

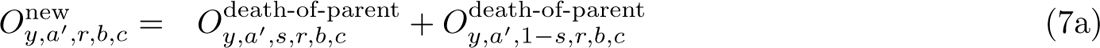

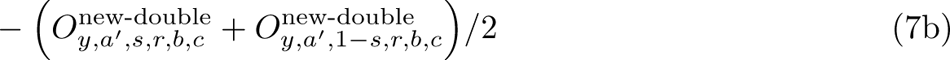

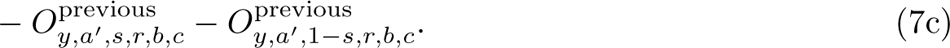

Eq. (7b) subtracts the children who lost in year *y* a parent due to cause *c* and the other parent due to any cause, which are counted twice in Eq. (7a). Without line-list family data and working from individual-level live birth and death statistics, the two terms *O*^new-double^, *O*^new-double^ are not identical and for this reason we subtracted the average of both. Eq. (7c) subtracts the children who already lost the other parent in previous years. In our previous studies on COVID-19 associated orphanhood, we did not consider the possibility of reinfection with COVID-19 and for this reason did not subtract children who already lost the other parent due to COVID-19 in previous years in these studies [Hillis et al., 2021a]. Further arguments show that across ages, standardized race & ethnic groups, and caregiver loss causes-of-death, the values in Eq. 7b-7c are approximately equal to orphanhood prevalence divided by four, and so do not exceed 1% of the average values in Eq. 7a.

To estimate the prevalence of orphanhood in year *y* = 2000*, …,* 2021, we accrued the number of children who newly experienced orphanhood in the previous 17 years and current year *y*, while accounting for ageing. Specifically, for each calendar year since 2000, we estimated the total number of children aged *b* = 0*, …,* 17 years in calendar year *y* and of race & ethnicity *r* who experienced orphanhood in their lifetime by cause-of-death *c* in one or both parents with

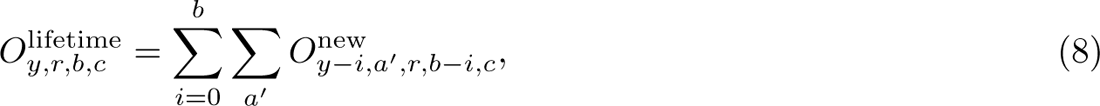

which sums over children who newly experienced orphanhood at younger ages in previous years.

Following Eqs. (7-8), we derived additional key quantities such as the number of children aged *b* in calendar year *y* = 1983*, …,* 2021 and standardized race & ethnicity *r* who newly experienced orphanhood in year *y* by parental sex *s* and cause-of-death *c*,

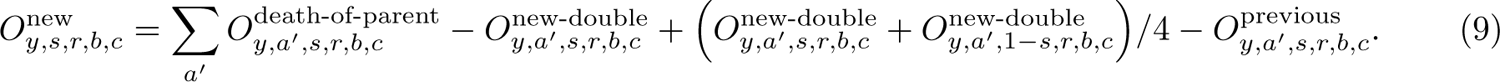

We chose the specific form of Eq. (9) over simpler approaches to ensure that the sum of *O*^new^ and equals Eq. 7. Further, we derived the number of children aged *b* in calendar year *y* = 2000*, …,* 2021 and standardized race & ethnicity *r* who experienced orphanhood in their lifetime by parent sex *s* and cause-of-death *c* by

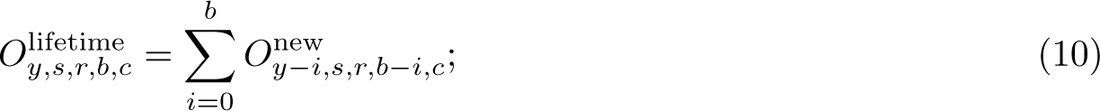

and the number of children of standardized race & ethnicity *r* who experienced orphanhood in calendar year *y* = 2000*, …,* 2021 in their lifetime by cause-of-death *c* in one or both parents by

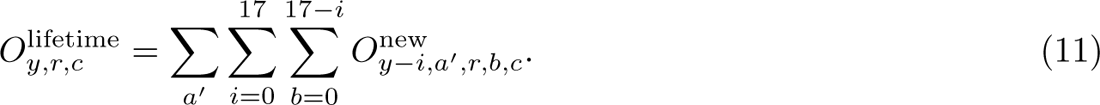

All other total numbers reported in this manuscript are aggregations of Eqs. (7-11).

### S1.6 Estimates of national-level grandparent caregiver loss, 1983-2021

We estimated the number of children who newly experienced grandparent caregiver death in year *y*, *y* = 1983*, …,* 2021 from the population-level mortality records in year *y* of U.S. residents aged 30 years and above (30+), and assuming that each decedent who is estimated to have been a grandparent caregiver leaves a minimum of one grandchild behind. Our approach follows the methods described in [Hillis et al., 2021a], with modifications to estimate the age of children who experience the death of a grandparent caregiver.

Starting from 2010, the American Community Survey (ACS) [Ellis and Simmons, 2014] collects data on the total number of adults aged 30+ years living with grandchildren of age 17 or under in the U.S., and proportions of these by sex, and separately by bridged-race and Hispanic origin [United States Census bureau, 2021].

For each year *y* = 2010*, …,* 2021, we estimated the number of adults aged 30+ years living with grandchildren of age 17 or under for each sex and standardized race & ethnicity by multiplying totals and proportions made available by the ACS. We then divided these numbers with the respective population size estimates to estimate sex- and standardized race & ethnicity-specific proportions of adults aged 30+ years who live with their grandchildren of age 17 or under, which we denoted with *γ_y,s,r_*. For *y* = 1983*, …,* 2009, we assumed that *γ_y,s,r_* is the same as in 2010. We further assumed that each grandparent caregiver leaves upon death a minimum of one child behind (corresponding to Eq. (2)) and estimated the minimum number of grandchildren who newly experienced grandparent caregiver death in year *y* = 1983*, …,* 2021, with a U.S. resident grandparent caregiver aged 30+ years, sex *s* and standardized race & ethnicity category *r* who died of leading cause *c* by

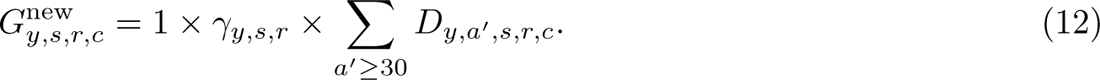

The ACS surveys did not collect data on whether both grandparents are alive and live with their grandchildren of age 17 or under, and for this reason we did not adjust Eq. (12) further for loss of other grandparents. We investigated in Sensitivity analyses our assumption that *γ_y,s,r_* was approximately constant from 1983 to 2010 using longitudinal United Nations Population Division data on Households and Living Arrangments of Older Persons for the U.S., which suggested that the proportion of older persons who live with children or who are the primary caregivers of children has in the U.S. remained fairly constant since 1990 (Supplementary Fig. S23).

To estimate the minimum number of children who experienced grandparent caregiver death in their lifetime, we additionally need disaggregations of Eq. (12) by single year of age *b* = 0*, …,* 17 years. For the central analysis, we assumed that the age composition of *G*^new^ is the same as the age composition of children who lost parents older than 30 years, are obtained from Eq. (7) by summing over the years *y* = 2000*, …,* 2021, and *c* is one of the leading caregiver loss causes-of-death. Supplementary Fig. S9 shows that the age compositions of children who lost parents older than 30 years

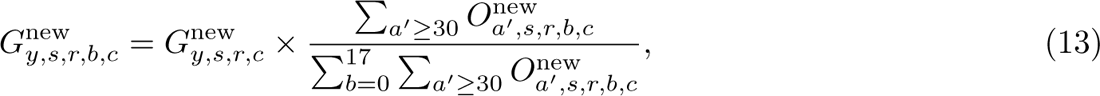

differ across causes-of-death while they are relatively stable across standardized race & ethnicity. It is plausible that the age composition of *G*^new^ may differ from theage composition of children who lost parents older than 30 years, and we explored alternative approaches to Eq. (13) in sensitivity analyses; none of these had a considerable impact on our overall estimates.

To obtain minimum estimates of caregiver loss, we then summed Eq. (9) and Eq. (13).

### S1.7 Uncertainty quantification in national estimates

This section describes how we derived uncertainty intervals in the national-level estimates of orphanhood incidence and prevalence (Eqs. (7-11)) and grandparent caregiver loss (Eq. (13)). To capture uncertainty, we followed the same phenomenological approach as in [Hillis et al., 2021a], and added Poisson noise around mortality, natality, and population size data for all year, sex, age, standardized race & ethnicity, and cause-of-death strata. We generated 10,000 Poisson noise data sets, and in terms of the uncertainty in mortality data, additionally considered uncertainty related to the harmonization of causes-of-death counts from 1983-1999 to the ICD-10 classification to avoid discontinuities in orphanhood estimates between 1998 and 1999. Specifically, we resampled the comparability ratios 10,000 times, assuming these were normally distributed with the standard deviations reported in Table 1 of [Anderson et al., 2001] where available, and multiplied these ratios with the Poisson noise mortality data. Then, we repeated orphanhood incidence and prevalence estimates on each data set, and calculated 2.5% and 97.5% quantiles to generate 95% uncertainty intervals around central estimates.

In the ACS data, we accounted for uncertainty in the estimated number of adults aged 30+ years living with grandchildren and in the attribution to sex, and standardized race & ethnicity groups using published uncertainty ranges. The ACS provides for each of their published estimates margins of error that correspond to 90% confidence intervals. We converted the 90% margins of error into standard deviations and bootstrap re-sampled the available totals and separate proportions 1,000 times assuming normal distributions around their central estimates and the corresponding standard deviations, multiplied the bootstrap resampled totals and proportions, and then divided by the corresponding population sizes. Across years, we found these sources of uncertainty amounted on average to 95% bootstrap intervals on the order of *±*0.79% of the central estimate of the number of Hispanic women aged 30+ years who live with grandchildren, *±*8.23% among Non-Hispanic American Indian or Alaska Native women, *±*1.93% among Non-Hispanic Asian women, *±*0.97% among Non-Hispanic Black women and *±*0.69% among Non-Hispanic White women; and similarly for men. We similarly generated 10,000 resampled data sets, repeated grandparent caregiver loss incidence and prevalence estimates on each data set, and then calculated 2.5% and 97.5% quantiles to generate 95% uncertainty intervals around central estimates.

**Supplementary Fig. S9:**
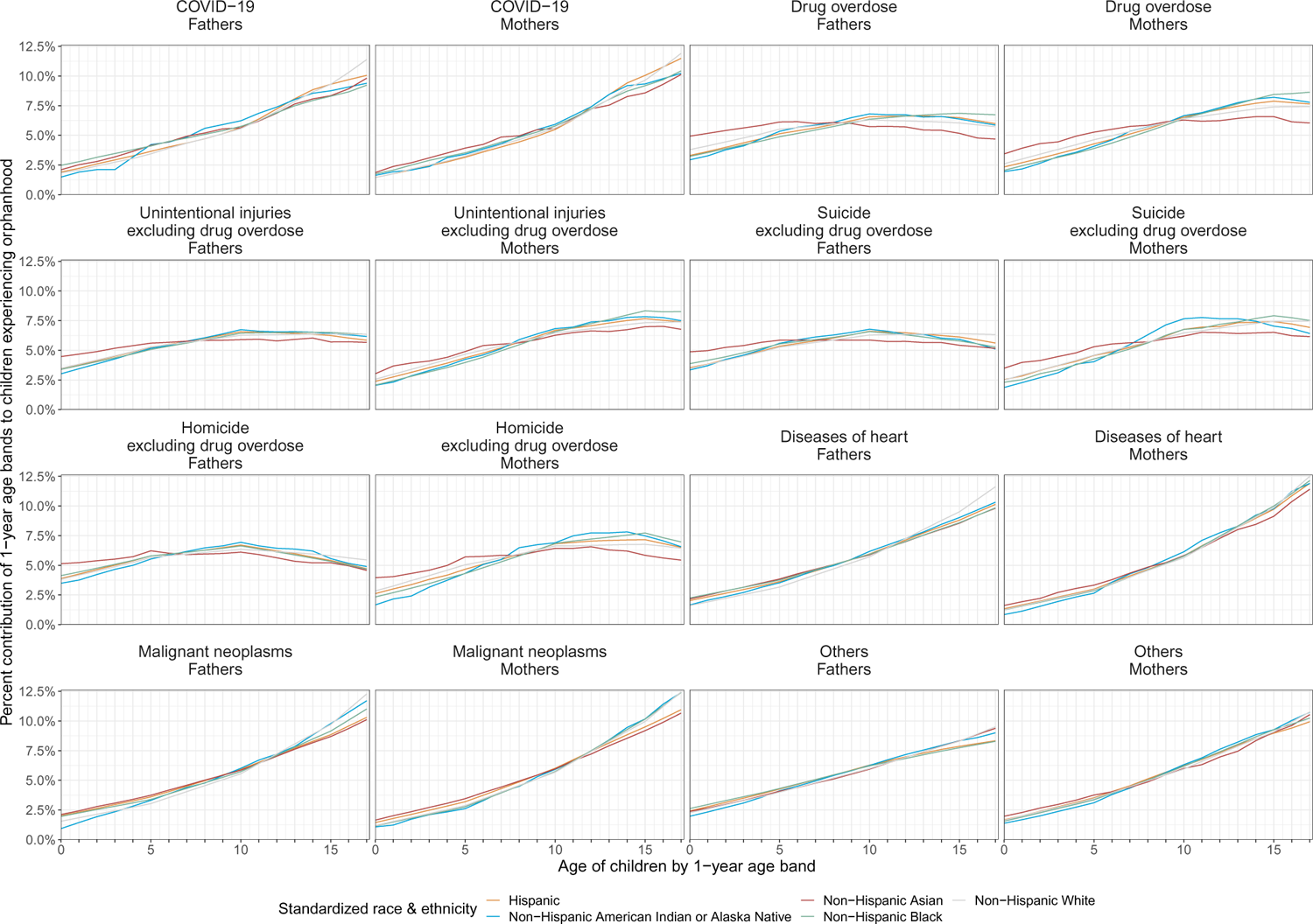
Percent contributions of 1-year age bands to children newly experiencing the loss of parents older than 30 years, stratified by standardized race & ethnicity and leading caregiver loss causes-of-death.

### S1.8 Estimates of state-level orphanhood, 2021

To guide policy at the state level, we further estimated orphanhood and grandparent caregiver loss in each of the 50 U.S. states and District of Columbia. We focussed on estimating the number of children who newly experienced orphanhood (incidence) and the number of children who experienced orphanhood in their lifetime (prevalence) in each state in 2021, by the leading caregiver loss causes-of-death (see Supplementary Table S9). To obtain state-level orphanhood prevalence estimates for 2021, as before we had to accrue and age state-level orphanhood incidence estimates from 2004, and to obtain state-level fertility rates from 1987. Overall, we proceeded analogously as for the national estimations, but obtained data from CDC WONDER [CDC, 2023] as state-level mortality data were not publicly available from NCHS from 2005 onwards [National Center for Health Statistics, 2023]. Counts below 10 were suppressed in CDC WONDER and for this reason we performed all estimations without stratification by standardized race & ethnicity. We adjusted for data suppression, and known biases in this approach that are due to correlations in mortality and fertility rates across standardized race & ethnicity. We note these are modelled adjustments and we cannot exclude that this resulted in bias in state-specific orphanhood estimates.

We extracted annual death counts by state, sex, age bands (15 *−* 19 years, *…*, 95 *−* 99 years, *>* 100 years), and cause-of-death (ICD-10 113 Selected Causes of Death) from the CDC WONDER mortality portal [CDC, 2023] from 2005 to 2021; and combined these data with the previously described, cause-specific NCHS mortality data sets from 2004 for which information on the state of residence of the decedent was publicly available. Throughout, we focused only on the eight leading caregiver-loss causes of death chronic liver disease and cirrhosis, COVID-19, diseases of the heart, drug overdose, homicide excluding drug overdose, malignant neoplasms, suicide excluding drug overdose, and unintentional injuries excluding drug overdose. For the leading caregiver-loss causes of death, the average discrepancy in the state-, sex-, age-, and cause-of-death-specific data from CDC WONDER was, when aggregated to national-level and compared to the corresponding NCHS mortality data, 22.9% among women and 13.5% among men. As a first step, we imputed suppressed entries with a value of 2, which resulted in average discrepancies of 6.80% among women and 2.53% among men. To account for these discrepancies, as a second step, we adjusted the state-level mortality counts according to

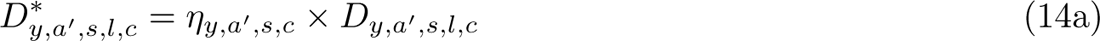

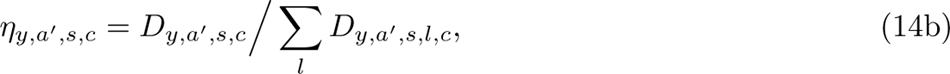

where *y* in 2005 to 2021, *a^′^* denotes the age band of the parent, *s* the sex of the parent and *c* the leading caregiver loss cause-of-death of the parent, and *D_y,a_′_,s,c_* are the national-level deaths derived from the line-list NCHS data in Eq. (3), and *D_y,a_′_,s,l,c_* are the state-specific deaths derived from CDC WONDER for each of the 50 U.S. states and District of Columbia, indexed by *l*. The suppression adjusted, cause-specific, state-level mortality counts matched the cause-specific, national-level mortality counts well, except when all deaths *D_y,a_′_,s,c,l_* were entirely suppressed across all states, which was the case for deaths due to homicide in women aged 55 years or older (Supplementary Fig. S10, compare pink versus blue). Overall, these suppression adjustments did not noticeably change the contribution of causes-of-death to mortality, because the majority of death counts were not suppressed when the data were aggregated to leading caregiver loss causes-of-death (Supplementary Fig. S11).

We next extracted live birth counts for mothers by state and age bands (15 *−* 19, *…*, 44 *−* 49 years) from the CDC WONDER natality portal [CDC WONDER, 2022] from 2005 to 2021; and combined these data with the previously described NCHS live birth data sets from 1987 to 2004 for which information on the state of residence of mothers was available. We proceeded analogously for fathers using the age bands 15 *−* 19, *…*, 44 *−* 49, 50 *−* 54, 55+ years from 2016 to 2021, as natality records were not available by demographic characteristics of fathers from 2005 to 2015. Data suppression was not an issue for the stratifications required to estimate orphanhood: for the years 2005-2021, the average discrepancy in the CDC WONDER and NCHS natality data sets was 0.026% for mothers and 0.032% for fathers (as compared to male NCHS live birth data with ages up to 77 years). To compute fertility rates at the state level, we further extracted age- and sex-stratified annual population size estimates for each state from 1987 to 1989 from the NIH National Cancer Institute Surveillance, Epidemiology, and End Results (SEER) database [National Cancer Institute, 2023]; and CDC WONDER from 1990 to 2021 [CDC, 2021c, 2022]. State-level fertility rates were calculated as in Eq. (1) when live births counts were available, but without stratification by race & ethnicity. For mothers, live birth counts were not publicly available for women aged 44-49 years in several years and 8 states (Alaska, Delaware, Montana, North Dakota, South Dakota, Vermont, West Virginia and Wyoming), and in these cases we interpolated state-specific fertility rates with locally estimated scatterplot smoothing (LOESS) as implemented in the R stat package version 4.2.3 with span argument 0.85. For Wyoming, no data were publicly available after 2019 and so interpolation was not possible and we used 2019 values. For fathers, live births counts were not publicly available from 2005 to 2015, and for these years we estimated state-specific fertility rates with LOESS with span argument 0.85, and using NCHS data from 2000 to 2004 and CDC WONDER data from 2016 to 2020 (Supplementary Fig. S12).

We then estimated state-specific incidence and prevalence of orphanhood from the suppression-adjusted state-level, cause-specific mortality counts and partially imputed fertility rates as outlined in Eqs. (7-11), with one modification. As described above the state-level mortality and fertility data were not disaggregated by race & ethnicity. To account for potential bias arising from correlations in mortality and fertility rates across race & ethnicity,

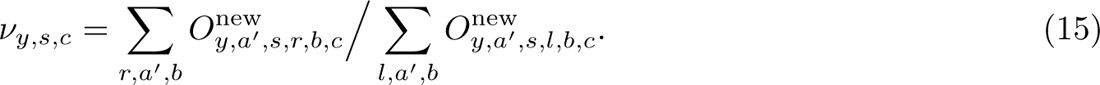

Overall, the correction factors *ν_y,s,c_* tended to be close to one, except for Homicide excluding drug overdose in women and ‘Other’ caregiver loss causes-of-death (Supplementary Fig. S13). We then adjusted the state-specific estimates as follows,

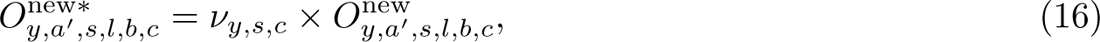

we compared the national-level estimates of the number of children who newly experienced orphanhood in (9) to the state-specific estimates with the correction factors
 and used Eq. (16) to calculate the state-specific analogues to Eqs. (8-11).

**Supplementary Fig. S10:**
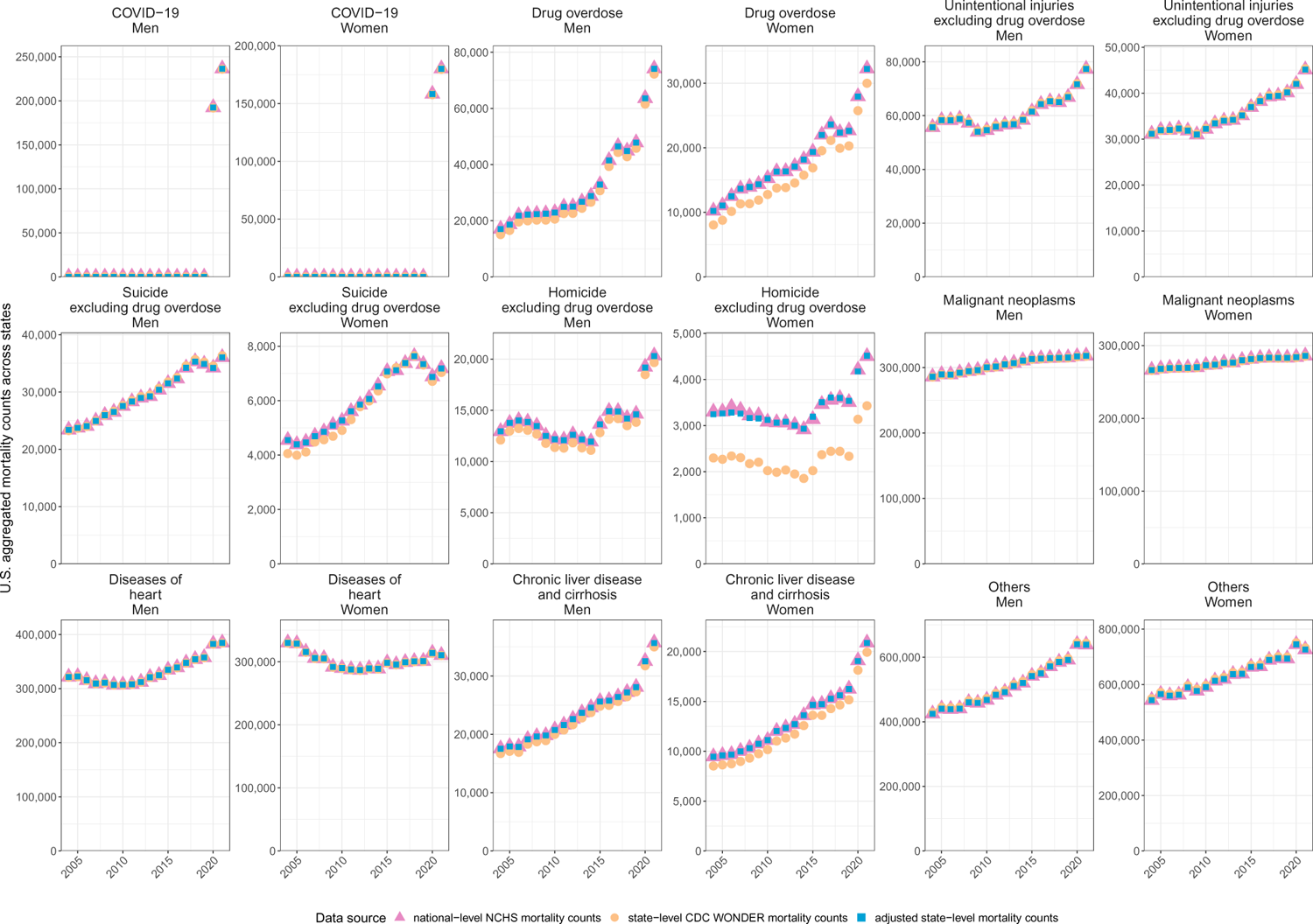
Completeness of state-level mortality counts. State-level mortality counts were extracted from CDC WONDER by year, sex, age band, and cause-of-death; and subject to suppression of counts below 10 individuals. Counts were aggregated to national level to assess their completeness as described in the Supplementary Text. Shown are the cause-specific, state-level mortality counts extracted CDC WONDER, the suppression-adjusted, cause-specific, state-level counts, and for comparison the cause-specific, national-level mortality counts obtained from NCHS.

We estimated the number of children who newly experienced grandparent caregiver death in year *y*, *y* = 2004*, …,* 2021 from the state-level mortality records in year *y* of U.S. residents aged 30 years and above (30+), and assuming that each decedent who is estimated to have been a grandparent caregiver in each state leaves a minimum of one grandchild behind. Overall, we proceeded as for the national-level estimation of grandparent caregiver loss as ACS also published state-specific data on adults aged 30+ years who live with grandchildren of age 17 or under [United States Census bureau, 2021]. For each year *y* = 2010*, …,* 2021, we calculated the expected number of adults aged 30+ years who live with their grandchildren of age 17 or under by multiplying the corresponding total co-resident numbers with the sex-specific proportions for each state; and then divided these with corresponding population sizes to obtain the corresponding proportions *γ_y,s,l_*. Overall, these proportions were considerably more uncertain than in the national-level analysis due to smaller ACS sample sizes (see below). For *y* = 2004*, …,* 2009, we assumed that *γ_y,s,l_*is the same as in 2010. We further assumed that each grandparent caregiver leaves upon death a minimum of one child behind (corresponding to Eq. (2)) and estimated the minimum number of grandchildren who newly experienced grandparent caregiver death in year *y* = 2004*, …,* 2021, with a U.S. resident grandparent caregiver aged 30+ years, sex *s* and state category *l* who died of leading caregiver loss cause-of-death *c* by

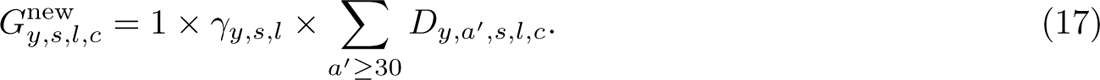

To estimate the minimum number of children who experienced grandparent caregiver death in their lifetime in each state, we additionally need disaggregations of Eq. (17) by single year of age *b* = 0*, …,* 17 years. For the central analysis, we assumed that the age composition of *G*^new^ is the same as the age composition of children who lost parents older than 30 years in the same state,

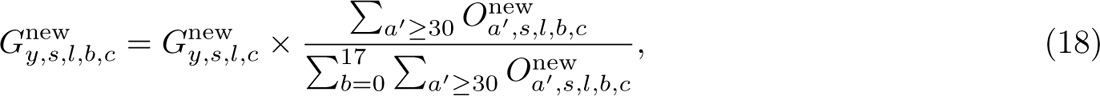

are obtained from Eq. (16) by summing over the years *y* = 2004*, …,* 2021, and *c* is one of the leading caregiver loss causes-of-death. Supplementary Fig. S14 shows that the age compositions of children who lost parents older than 30 years differed across causes-of-death while they are relatively stable across states.

**Supplementary Fig. S11:**
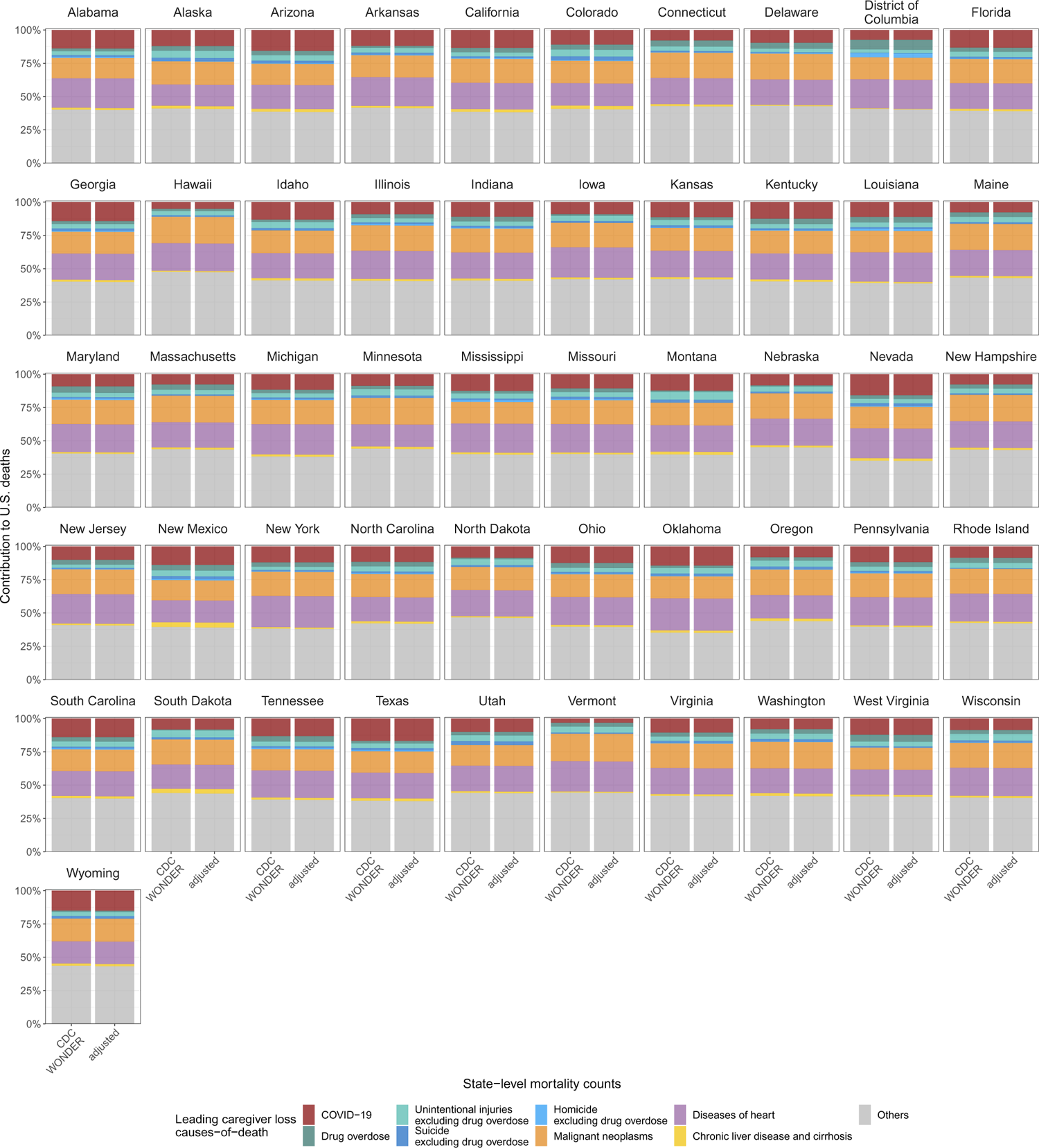
Contribution of leading causes-of-death to U.S. deaths by states. State-level mortality counts were extracted from CDC WONDER by year, sex, age band, and cause-of-death and adjusted for suppression. Shown are the contributions of leading causes-of-death to the unadjusted state-specific deaths in 2021, and the adjusted state-specific deaths in 2021.

**Supplementary Fig. S12:**
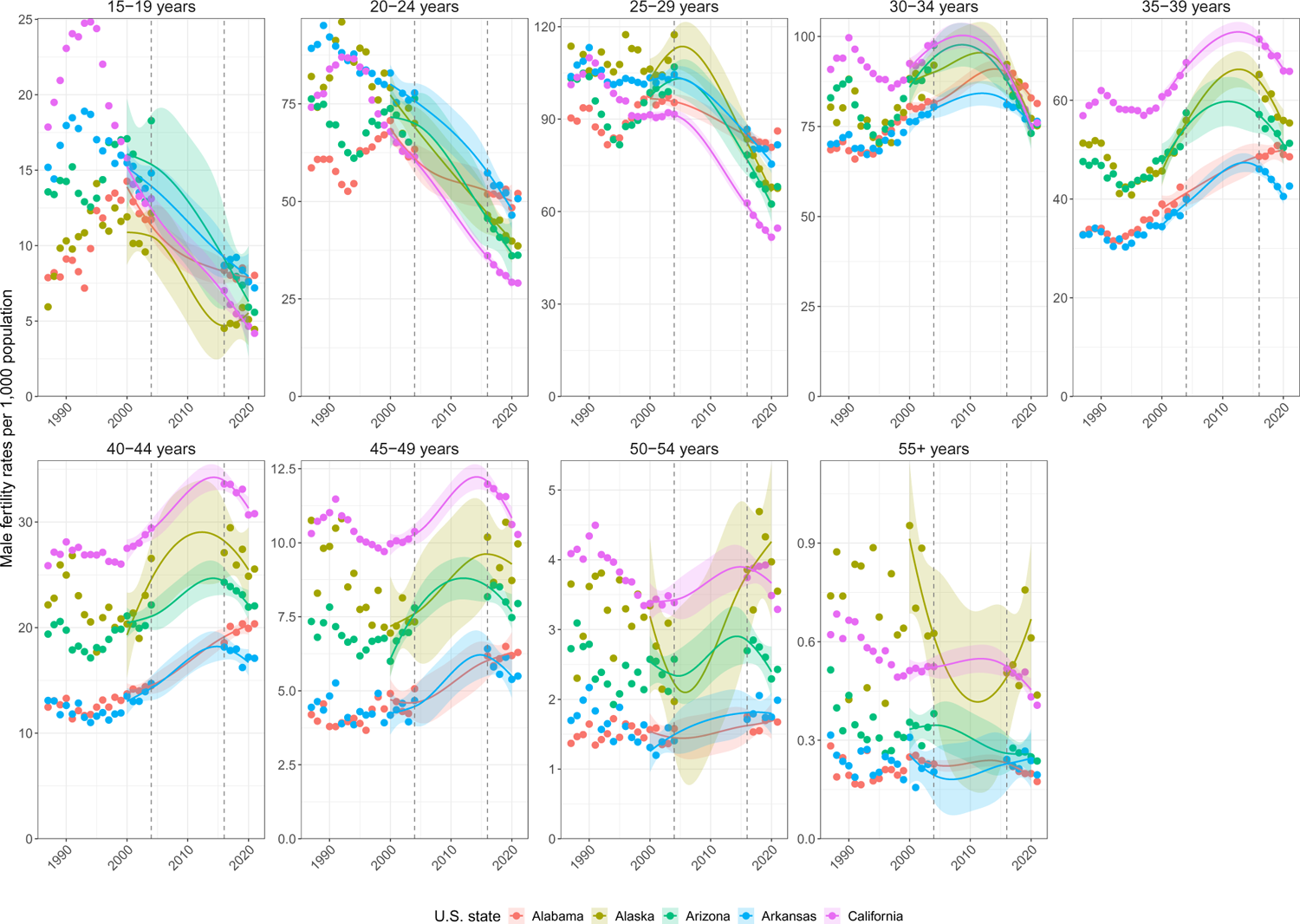
Interpolation of missing state-specific male fertility rates. State-level live birth counts were extracted from CDC WONDER by year, sex, and age band; and combined with line-list NCHS live birth data up to 2004 when information on state residence was made available. For fathers, demographic information was only available since 2016 in CDC WONDER and state-specific male fertility rates were interpolated with LOESS estimators as described in the Supplementary Text. Shown are empirical male fertility rates (dots) for 5 states from 1987 to 2004 and from 2016 to 2021, along with the LOESS estimates (lines) for 2000 to 2020 and 95% prediction intervals (ribbon).

### S1.9 Uncertainty quantification in state-level estimates

This section describes how we derived uncertainty intervals in the state-level estimates of orphanhood incidence and prevalence (Eqs. (7-11)) and grandparent caregiver loss (Eq. (13)). Overall, the approach here is similar to the national-level uncertainty analysis.

We added Poisson noise around state-specific mortality, natality, and population size data for all year, sex, age, and cause-of-death strata. We generated 10,000 Poisson noise data sets, repeated orphanhood incidence and prevalence estimates on each data set, and then calculated 2.5% and 97.5% quantiles to generate 95% uncertainty intervals around central estimates.

To estimate grandparent caregiver loss, we accounted for uncertainty in the estimated number of adults aged 30+ years living with grandchildren by U.S. state using published uncertainty ranges. The ACS provides for each of their state-specific estimates margins of error that correspond to 90% confidence intervals. We converted the 90% margins of error into standard deviations and bootstrap re-sampled the available totals and separate proportions 10,000 times assuming normal distributions around their central estimates and the corresponding standard deviations, and then multiplied the sampled totals and proportions. As expected given available ACS sample sizes, across years we found that uncertainty in the proportions *γ_y,s,l_* was in some states considerably larger than for the national race & ethnicity-specific analysis, with 95% bootstrap intervals of up to *±*13.5% of the central estimates (Supplementary Fig. S15). We similarly generated 10,000 resampled data sets, repeated grandparent caregiver loss incidence and prevalence estimates on each data set, and then calculated 2.5% and 97.5% quantiles to generate 95% uncertainty intervals around central estimates.

### S1.10 Estimates of racial & ethnic groups impacted by leading causes of caregiver loss in U.S. states, 2021

Characterizing the racial & ethnic groups of children that are impacted by orphanhood and grandparent caregiver loss at state-level is challenging due to limits in publicly available data. We focused on estimating and comparing 2021 orphanhood and grandparent caregiver loss prevalence rates by standardized race & ethnicity in the 10 U.S. states with highest orphanhood prevalence rates and for the primary (first-ranked) caregiver-loss cause-of-death only, West Virginia (‘Drug overdose’), New Mexico (‘Drug overdose’), Mississippi (‘Unintentional injuries’), Louisiana (‘Drug overdose’), Kentucky (‘Drug overdose’), Tennessee (‘Drug overdose’), Alabama (‘Diseases of heart’), Oklahoma (‘Unintentional injuries excluding drug overdose’), Ohio (‘Drug overdose’) and Florida (‘Drug overdose’). Throughout, we obtained publicly available data from CDC WONDER [CDC, 2023] from 2005 onwards [National Center for Health Statistics, 2023] and adjusted for data suppression, but note these are modelled adjustments and we cannot exclude that this resulted in bias in state-specific orphanhood estimates.

Annualized live birth counts were extracted as described for the state-level analysis for 2005 to 2019, but now also stratified by bridged-race and Hispanic origins, and then combined with NCHS live births data from 1987 to 2004. For 2020-2021, live birth counts to women were not available by bridged-race at state-level. Analogously, annual death counts for the leading caregiver loss causes-of-death were compiled as described above for 2005 to 2021, and stratified into the standardized race & ethnicity categories in Supplementary Table S7, and then combined with the NCHS mortality data sets from 2004. Suppressed values were imputed by 1 and adjusted further for suppression as in Eq. (14). We followed previous criteria on data reliability and excluded from consideration for each state those race & ethnicity groups that had more than two age bands with fewer than 20 live birth counts Mathews and Hamilton [2019]. Supplementary Table S6 lists the corresponding strata as ‘small populations’, and also describes the average completeness of live birth records for the years 1995-2004 when directly comparable line-list live birth records were also available from NCHS. For the remaining year-state-sex-race & ethnicity strata, we considered the corresponding mortality data for the leading caregiver-loss cause-of-death. Again, we excluded from consideration those standardized race & ethnicity groups that had more than two age bands with fewer than 20 death counts Mathews and Hamilton [2019]. Supplementary Table S6 lists the corresponding strata as ‘small death counts’, and also describes the average completeness of death records for the years 1999-2004 when directly comparable line-list death records were also available from NCHS. Supplementary Fig. S16 illustrates the state and standardized race & ethnicity specific live birth data from CDC WONDER, the line-list live birth data from NCHS and the state and standardized race & ethnicity specific live birth data for the three states with highest orphanhood prevalence rates in 2021. Similarly, Supplementary Fig. S17 illustrates the state and standardized race & ethnicity specific mortality data from CDC WONDER for the primary caregiver loss cause-of-death, the line-list live death from NCHS and the adjusted state and standardized race & ethnicity specific mortality data for the three states with highest orphanhood prevalence rates in 2021. Population-size estimates for each state were obtained from the CDC WONDER population size portal for 1990 to 2021 [CDC, 2021c, 2022] by bridged-race, and converted to the standardized race & ethnicity categories described in Supplementary Table S11.

**Supplementary Fig. S13:**
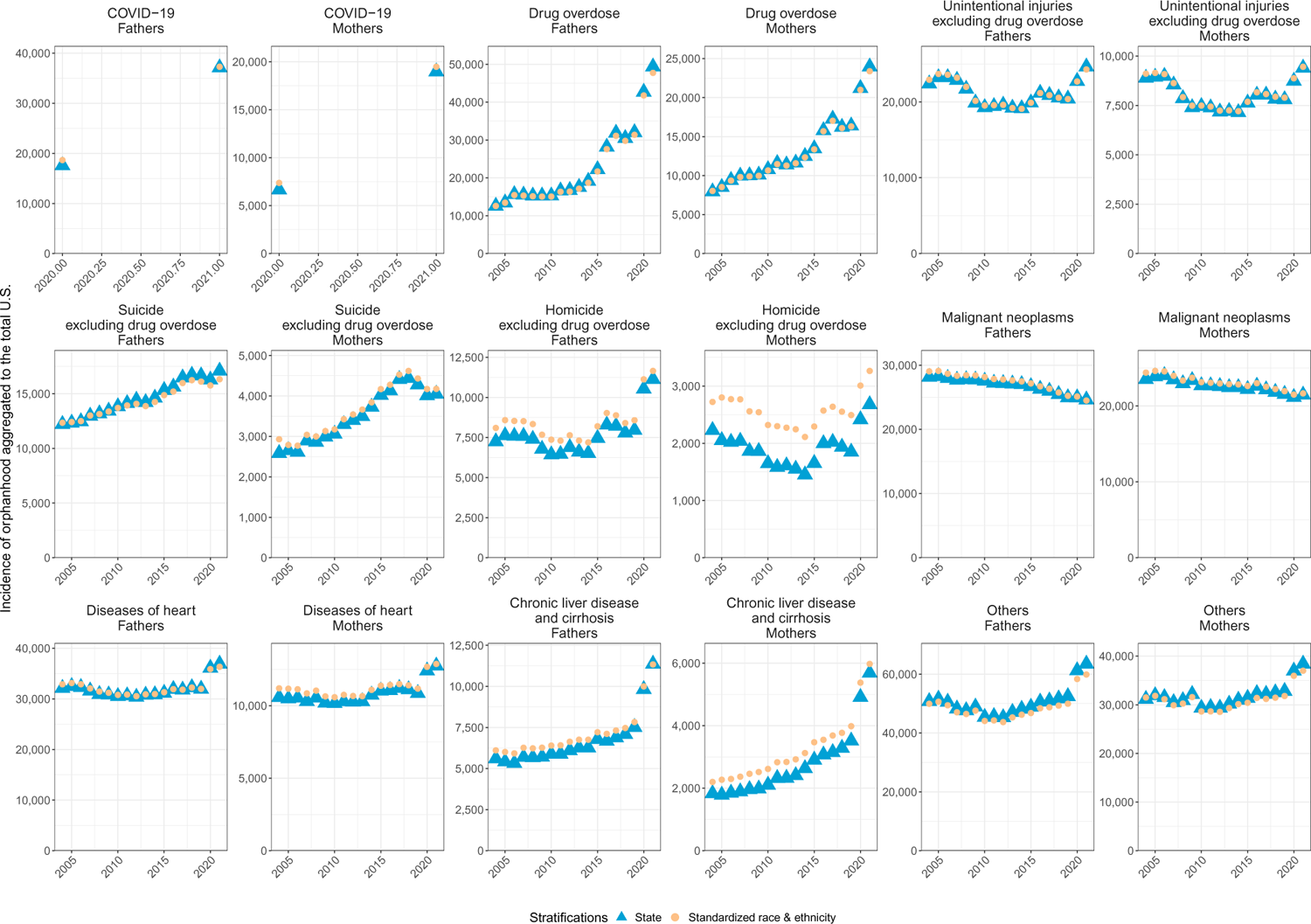
Comparison of estimated national-level incidence of orphanhood and estimated state-level incidence of orphanhood. At the national level, incidence of orphanhood was estimated from line-list live birth and death data stratified by race & ethnicity; whereas as the state-level, incidence of orphanhood was estimated from aggregated and partially interpolated live birth data and partially suppressed cause-specific deaths that were not stratified by race & ethnicity. Shown are incidence estimates using either approach by leading caregiver loss causes-of-death. Overall, incidence estimates were very similar except for ‘Homicide excluding drug overdose’ in women, ‘Chronic liver disease and cirrhosis’ in women and ‘Others’ causes-of-death. State-level incidence estimates were adjusted to match the national-level totals as described in the Supplementary Text.

**Supplementary Fig. S14:**
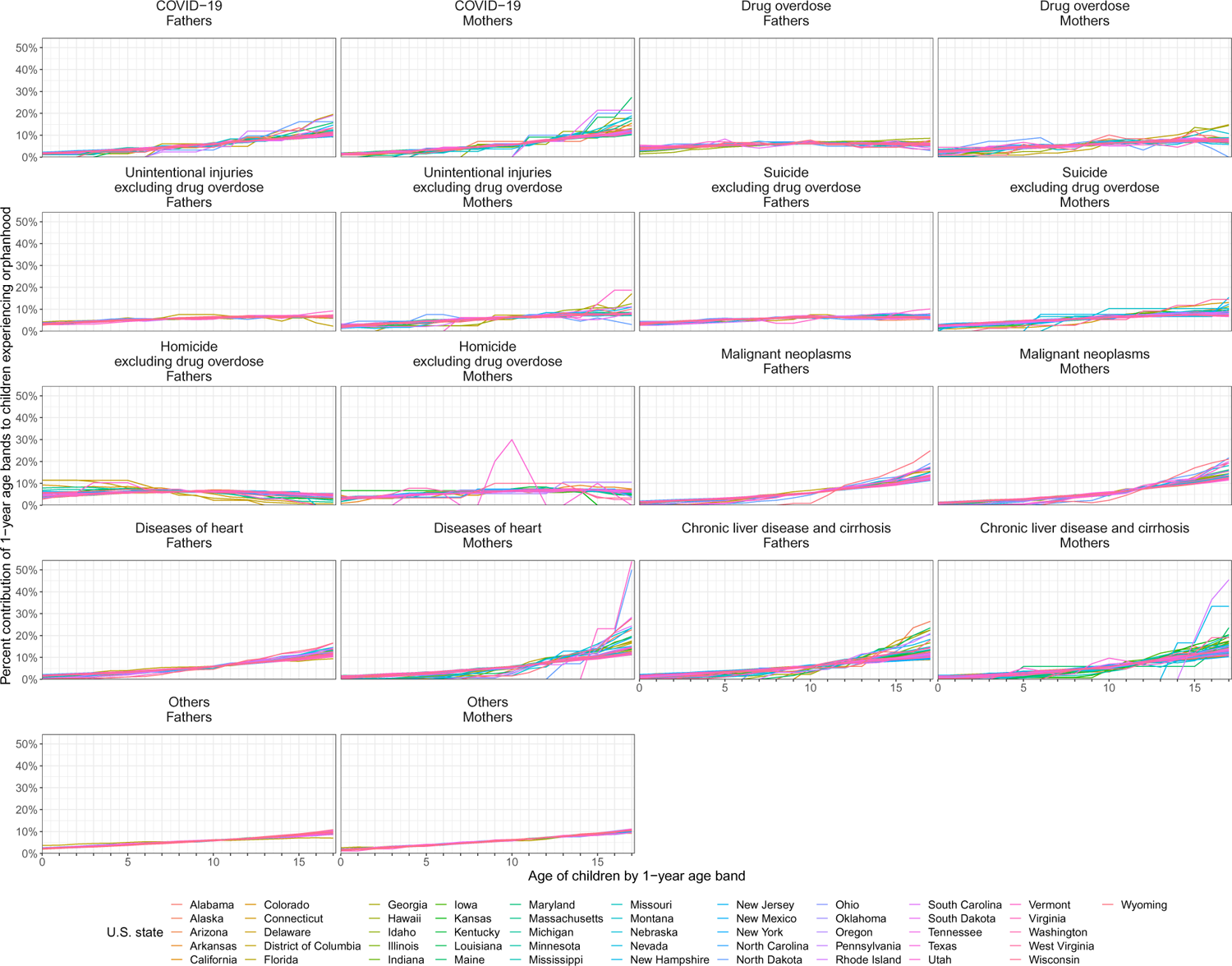
State-specific percent contributions of 1-year age bands to children newly experiencing the loss of parents older than 30 years. Percent contributions were stratified by sex and leading caregiver loss causes-of-death of parents (facets) and are shown for each U.S. state (colours).

**Supplementary Fig. S15:**
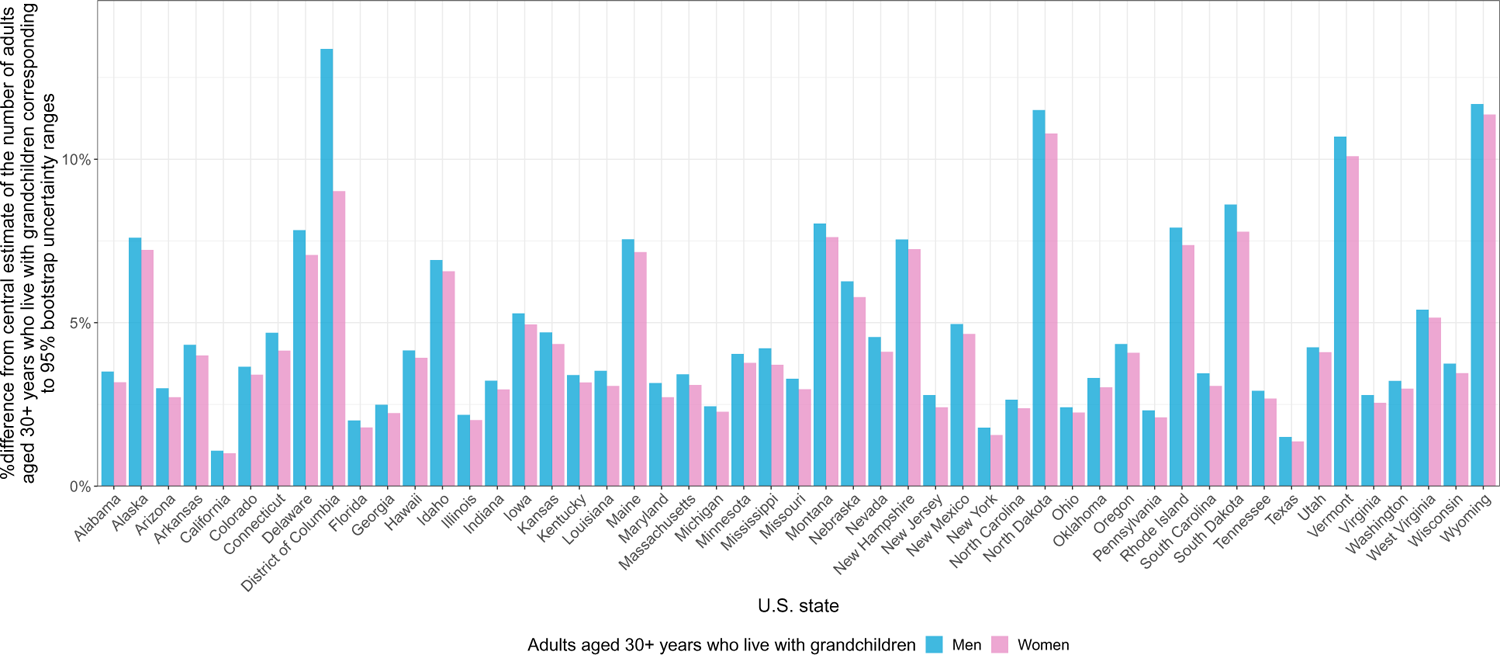
Magnitude of uncertainty in the number of adults aged 30+ who live with grandchildren in each U.S. state. Estimates of uncertainty were derived using bootstrap resampling from the margins of error reported by ACS for each of their estimates. Uncertainty for women and men is shown separately in colour.

**Supplementary Fig. S16:**
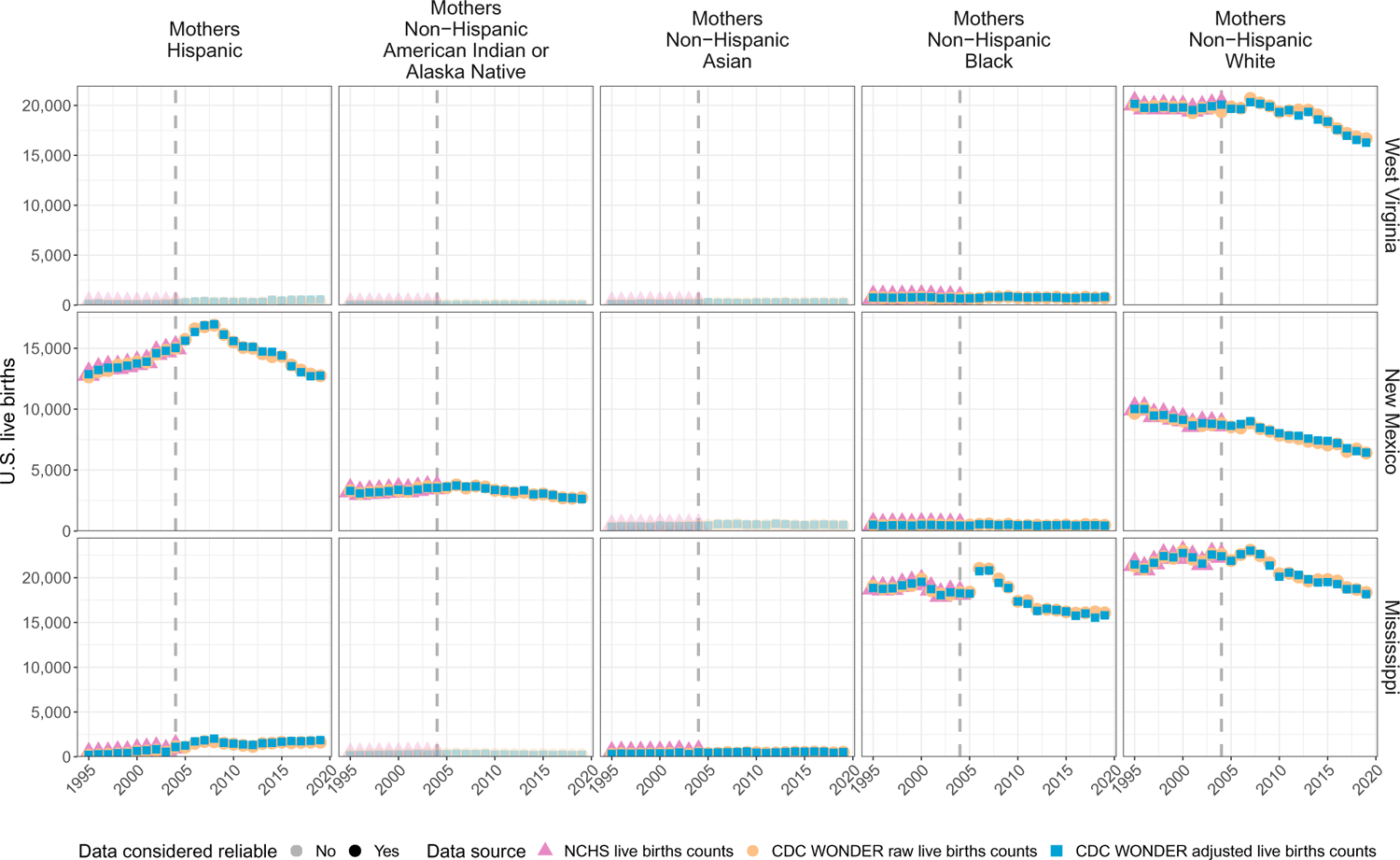
Data completeness evaluation for race & ethnicity- and state-specific live birth counts for three U.S. states, West Virginia, New Mexico and Mississippi. Standardized race & ethnicity live birth counts were extracted from CDC WONDER by state, year, sex, and age band; and subject to suppression of counts below 10 individuals. Counts from NCHS were aggregated to state-level by standardized race & ethnicity to assess their completeness as described in the Supplementary Text. Shown are the state-level live birth counts extracted from CDC WONDER, the suppression-adjusted, standardized race & ethnicity-specific counts, and for comparison the live birth counts obtained from NCHS by the same stratification. Points shown in opaque colours represent standardized race & ethnicity categories that were excluded from further analysis due to the extent of suppressed data.

**Supplementary Fig. S17:**
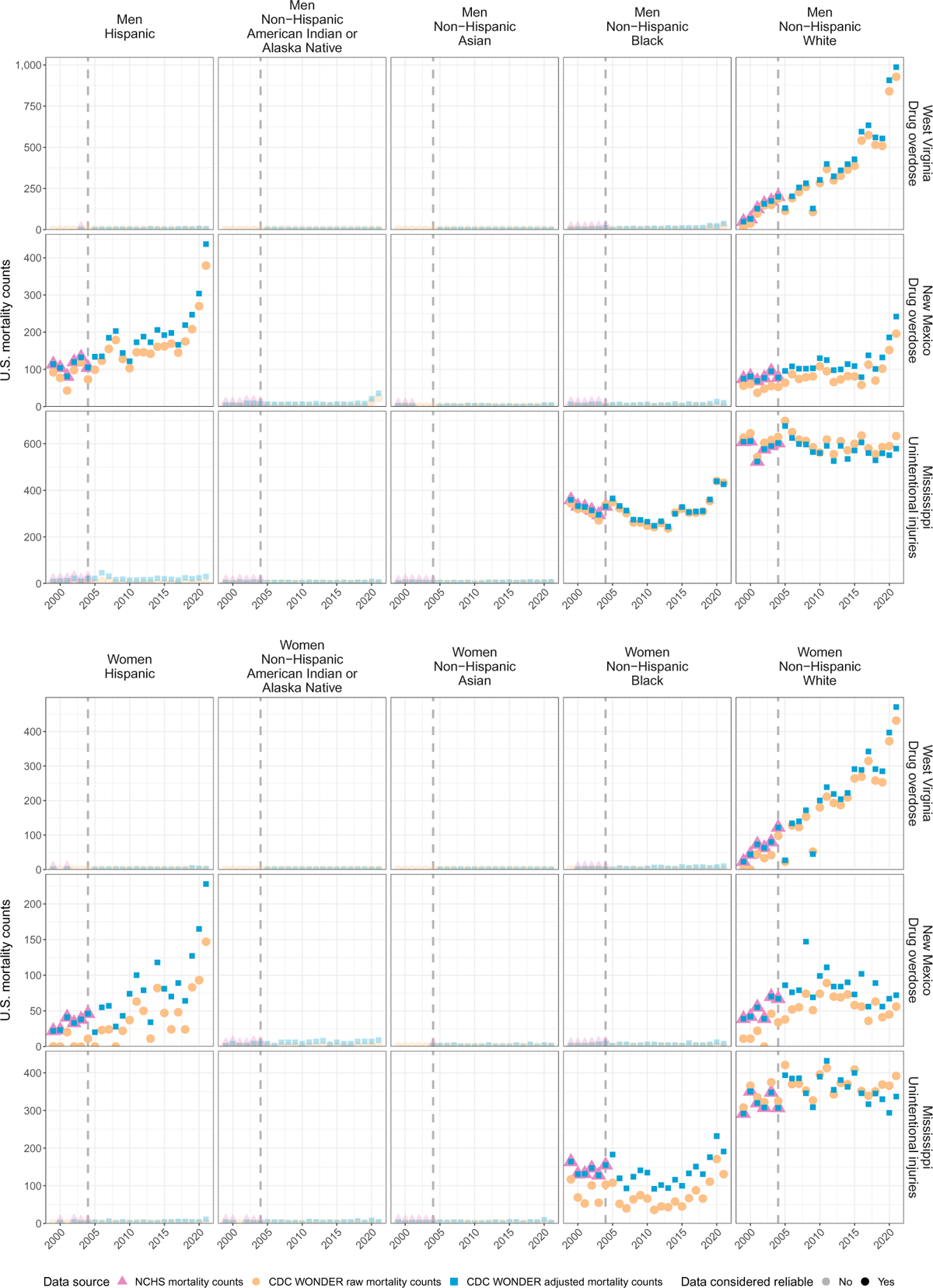
Data completeness evaluation for race & ethnicity- and state-specific mortality counts for three U.S. states, West Virginia, New Mexico and Mississippi. Standardized race & ethnicity-specific mortality counts were extracted from CDC WONDER by state, year, sex, age band, and leading caregiver loss cause-of-death; and subject to suppression of counts below 10 individuals. Counts were aggregated to state-level by standardized race & ethnicity to assess their completeness as described in the Supplementary Text. Shown are the mortality counts extracted from CDC WONDER, the suppression-adjusted, race & ethnicity-specific counts, and for comparison the mortality counts obtained from NCHS by the same stratification. Points shown in opaque colours represent standardized race & ethnicity categories that were excluded from further analysis due to the extent of suppressed data.

We estimated race & ethnicity- and state-specific incidence and prevalence of orphanhood as outlined in Eqs. (7-11), except that female race & ethnicity- and state-specific fertility rates in 2020-2021 were as assumed to be as in 2019 due to limitations in publicly available data. To investigate estimation accuracy, we compared the resulting sum of the race & ethnicity- and state-specific orphanhood incidence estimates to the previous state-specific orphanhood incidence estimates (Supplementary Fig. S18). For Alaska and Oklahoma, the sum of the race & ethnicity- and state-specific orphanhood incidence estimates was more than 20% below the state-specific orphanhood incidence estimates attributable to the leading caregiver loss cause-of-death. Supplementary Table S6 lists these states as ‘large discrepancy in estimates’. The table then reports, for all remaining states, 2021 race & ethnicity- and state-specific orphanhood prevalence rate estimates that are attributable to the leading caregiver loss cause-of-death.

**Supplementary Fig. S18:**
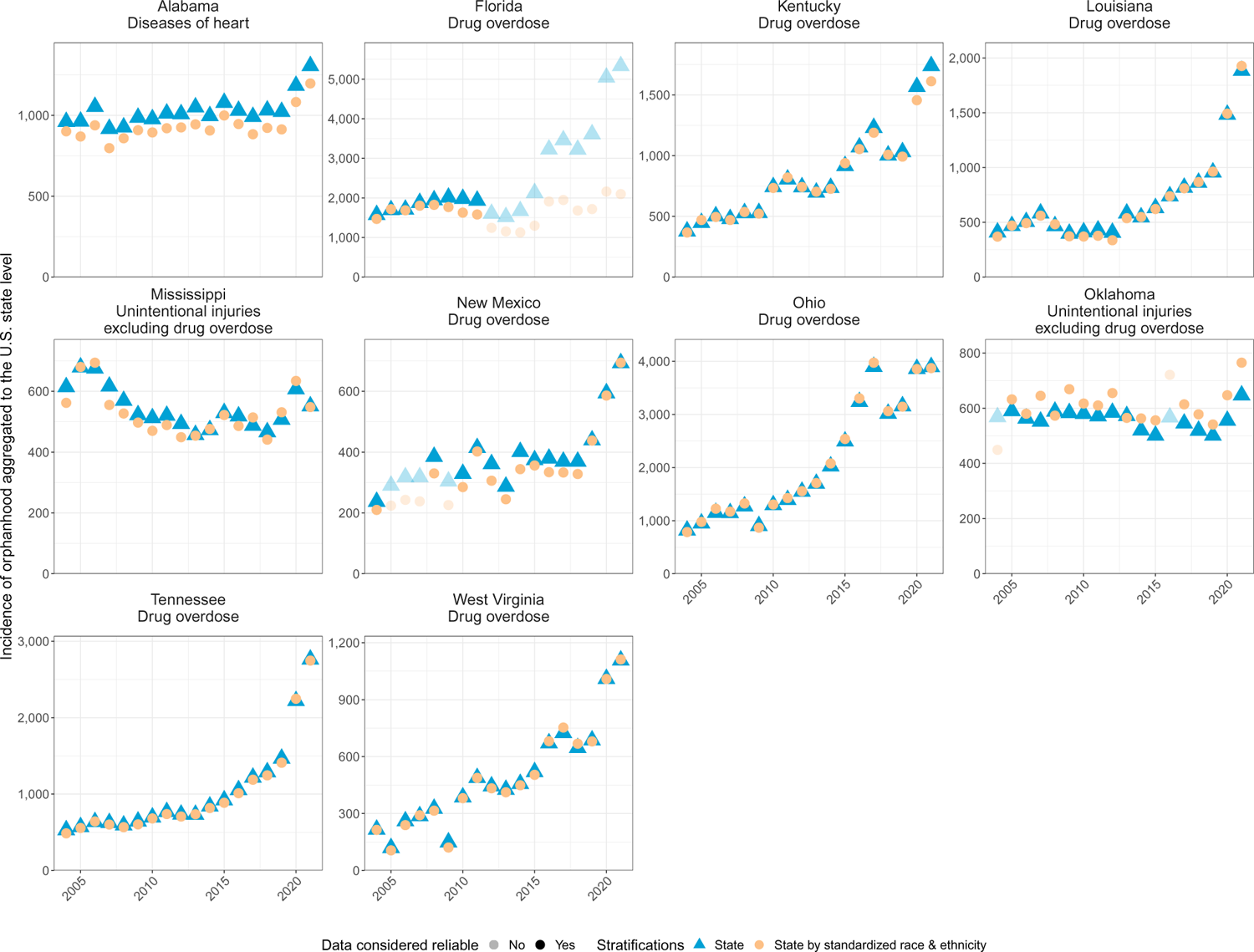
Reliability evaluation of orphanhood incidence estimates by state, year, standardized race & ethnicity and leading caregiver loss cause-of-death. Race & ethnicity- and state-specific orphanhood incidence rates for the leading caregiver loss cause-of-death were estimated from suppression-adjusted live births and deaths counts from CDC WONDER as described in the Supplementary Text. To assess the reliability of these estimates, we considered weighted averages of the orphanhood incidence estimates across race & ethnicity (yellow triangles), and then compared the weighted averages against state-level estimates (blue circles). Triangles and circles are shown in opaque colour when race & ethnicity-specific estimates disagreed by more than 20% and were considered unreliable.

## S2 Sensitivity analyses

### Sensitivity in mortality data

In the central analysis, we derived mortality counts by year, sex, age band, standardized race & ethnicity and caregiver loss causes-of-death from NCHS line-list mortality records from 1983 to 2021. Aggregate mortality counts are also available from CDC WONDER CDC [2023] from 1999 to 2021. We extracted data from the NCHS Vital Statistics portal because this allowed us to use the same data source across all years, bypass data suppression, and incorporate uncertainties in standardized race & ethnicity reporting. The CDC WONDER mortality counts allowed us to check our in-house data aggregations, if we obtain data from CDC WONDER at coarser population strata. For this purpose, we extracted annual death counts by sex, and age bands (15-19 years, *…*, 95-99 years, *>*100 years) from the CDC WONDER mortality portal CDC [2023] from 2000 to 2021. The overall mortality counts that we aggregated from the NCHS line list data were identical with the CDC WONDER mortality data without cause-of-death stratification. Secondly, we extracted annual death counts by sex, age bands (15-19 years, *…*, 95-99 years, *>*100 years) and cause-of-death (ICD-10 113 Selected Causes of Death) from the CDC WONDER mortality portal CDC [2023] from 2000 to 2021, without stratification by race & ethnicity. We then compared the mortality counts from the two data sources for each year and the leading caregiver loss causes-of-death (Supplementary Fig. S19). We found that across years, the maximum discrepancy in the CDC WONDER counts relative to the NCHS mortality counts was 0.0068% in women and 0.0057% in men, which reflected data suppression especially due to homicide excluding drug overdose in women, and overall indicated consistency of our data with that from CDC WONDER.

### Sensitivity in live births data

In the central analysis, we derived live birth counts by year, sex, age band, and standardized race & ethnicity from NCHS line-list natality records from 1969 to 2021. Aggregate live birth counts are also available from CDC WONDER CDC WONDER [2022] from 1995 to 2021. We chose to extract data from the NCHS Vital Statistics portal because this allowed us to use the same data source across all years. The CDC WONDER live birth counts allowed us to check our in-house data aggregations. For this purpose, we extracted annual live birth counts from the CDC WONDER natality portal for women by age bands 15-19 years, *…*, 45-49 years from 2000 to 2021, and separately for men by age bands 15-19 years, *…*, 55+ years from 2016 to 2021 without stratification by race & ethnicity, which avoids data suppression. We then compared the live birth counts from the two data sources for each year and found that both data sets matched across all years.

### Sensitivity in national-level orphanhood estimates to assumptions on historic fertility rates

In the central analysis, we assumed that male and female fertility rates by age and standardized race & ethnicity were in 1966-1989 constant and as in 1990 (Supplementary Fig. S7). This assumption was made because population size data stratified by detailed race categories (more than three categories) were not publicly available before 1990. Considering that age- and sex-specific live births data were available by race & ethnicity since 1978, we estimated in this sensitivity analysis the historic composition of population sizes by race & ethnicity in 1980-1989, and then updated the corresponding fertility rates and national-level orphanhood incidence and prevalence estimates. More specifically, we considered time trends in the proportion of each race & ethnicity in each 5-year age band and both sexes in the U.S. intercensal population size estimates in 1990 to 1995 (Supplementary Fig. S20A), and extrapolated these with linear models backwards in time to 1980-1989. We do not think that this estimation approach resulted in accurate estimates of population sizes, but rather that this approach conveys possible sensitivities in our orphanhood estimates. We identified minor sensitivities to national orphanhood prevalence estimates up to 2007, with no impact on incidence and prevalence estimates for recent years (Supplementary Fig. S20).

### Sensitivity in national-level orphanhood estimates to potentially correlated fertility rates

In the central analysis, we assumed that population-level fertility rates were not correlated with population-level mortality rates. We considered in sensitivity analyses possible deviations from this assumption that might lead to upwards bias in our central estimates. For example, a person who died in year *y* may have experienced poor health in the preceding years *y −* 1, *y −* 2, *…* and in this case may also have been less likely to mother or father a child in years *y*, *y −* 1, *y −* 2, We modelled this possibility through four scenarios of dampened fertility rates close to individual death events. Specifically, we considered cumulative logistic adjustment factors that dampened fertility rates to either zero or half of the corresponding population-level, year-, age-, sex-, and standardized race & ethnicity-specific fertility rates in the year of death *y*. We also considered sensitivity analyses so that the onset of lower fertility rates preceded the year of death *y* by 1 or 3 years as shown in Supplementary Fig. S21A. We found that orphanhood incidence estimates were up to 8.4% lower than in the central analysis, and orphanhood prevalence estimates were up to 15% lower than in the central analysis (Supplementary Fig. S21B). It is also possible that our central orphanhood estimates might be biased downward. For example, women experiencing premature death are more likely economically disadvantaged and in turn are more likely to have had reduced access to contraceptives and corresponding higher fertility rates [Price et al., 2013], or earlier sexual debut resulting in higher cumulative fertility until death. It is also possible that overall, at population-level, there is no strong correlation between fertility and mortality rates especially as the lag between birth and death events is typically relatively large, and in the absence of data we have opted for this middle approach in the central analysis.

**Supplementary Fig. S19:**
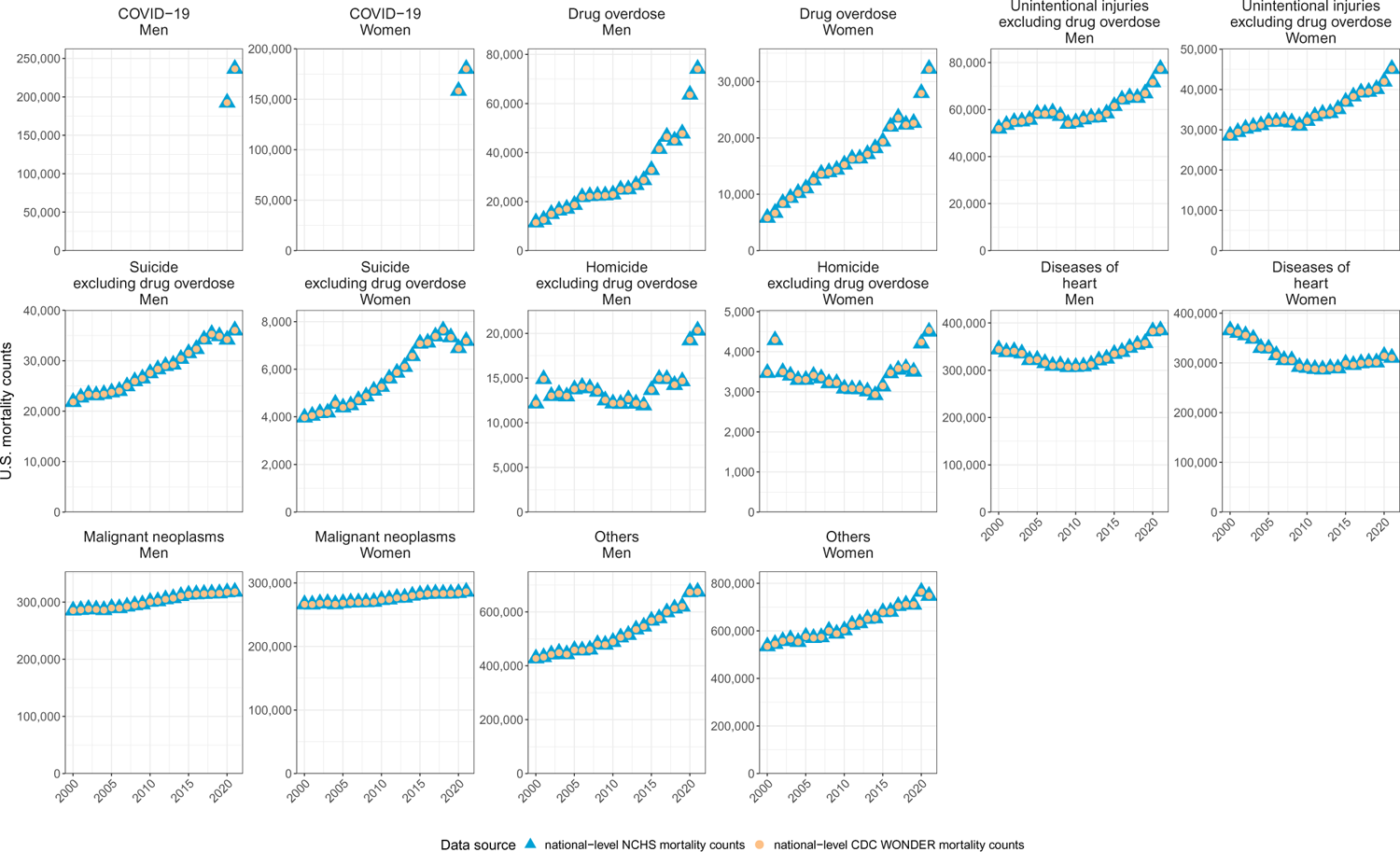
Comparison of national-level U.S. mortality counts from NCHS and CDC WONDER. In the central analysis, we derived mortality counts by year, sex, age band, standardized race & ethnicity and caregiver loss causes-of-death from NCHS line-list mortality records from 1983 to 2021. To check consistency of our data aggregations, we extracted annual death counts by sex, age bands (15-19 years, *…*, 95-99 years, *>*100 years) and cause-of-death (ICD-10 113 Selected Causes of Death) from CDC WONDER (i.e. without further stratification by standardized race & ethnicity). Mortality counts from both data sources were identical except for deaths due to homicide excluding drug overdose in women, and overall indicated consistency of our data with that from CDC WONDER.

### Sensitivity in national-level grandparent caregiver loss estimates to assumptions on the age of children experiencing loss of a grandparent caregiver

In the central analysis, we considered in Eq. (13) as a proxy to the age composition of children experiencing grandparent caregiver loss the age composition of children experiencing orphanhood. Calculations were done independently for each leading caregiver loss cause-of-death. We performed two analyses to characterise the sensitivity of this approach to national-level grandparent caregiver loss estimates. First, we repeated calculations using as proxy the age composition of children experiencing orphanhood across all causes of caregiver loss, i.e.

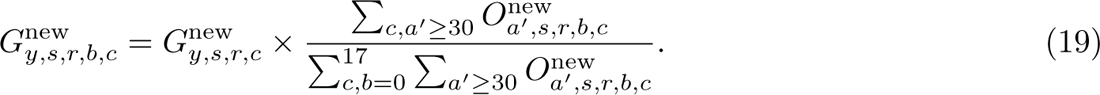

**Supplementary Fig. S20:**
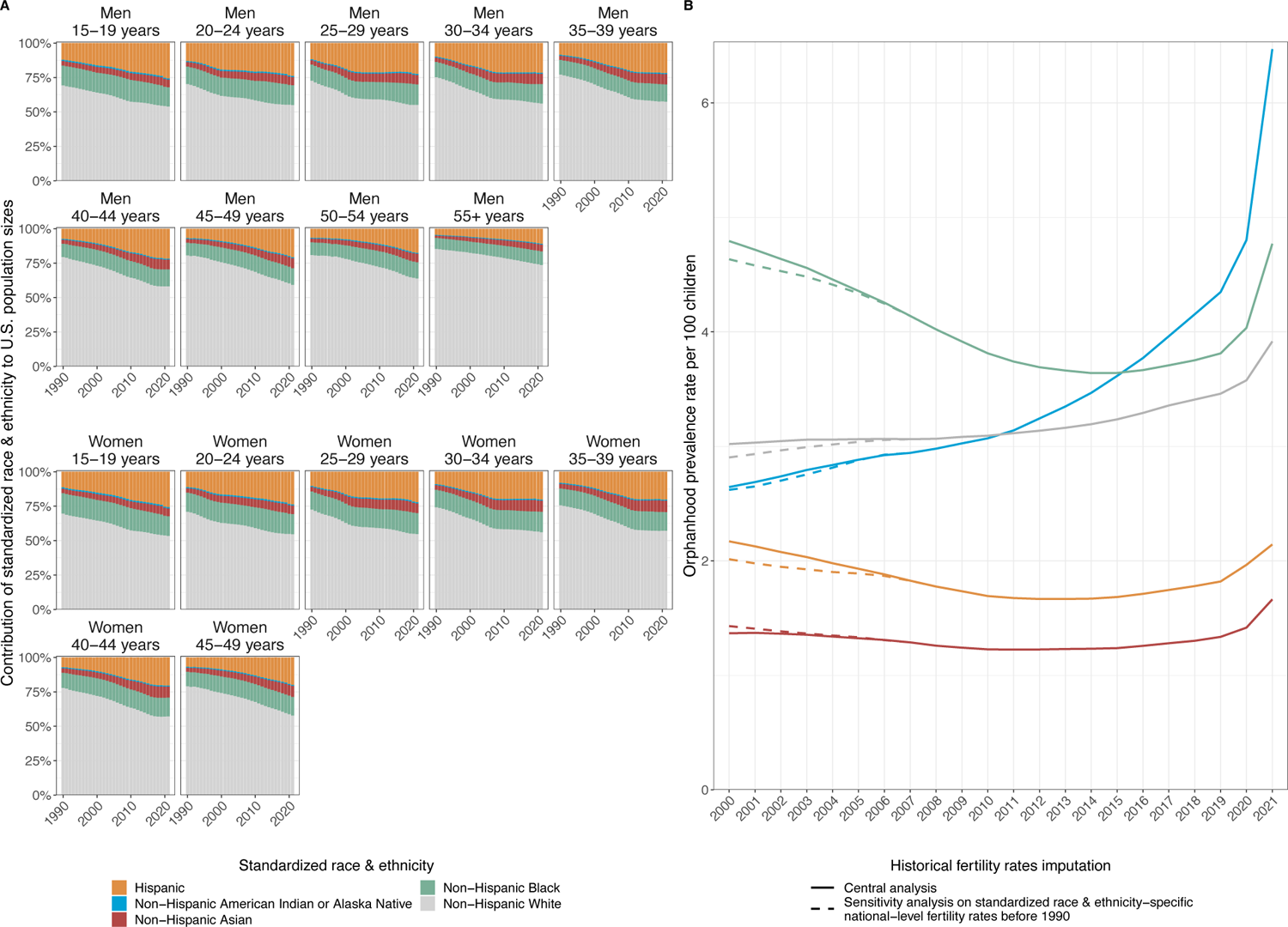
Sensitivity in national-level orphanhood estimates to assumptions on historic fertility rates. (A) Composition of U.S. population by standardized race & ethnicitiy (fill colours) from 1990 to 2021, for both men and women in the age bands relevant for orphanhood estimation. (B) Estimated national-level orphanhood prevalence rates by standardized race & ethnicity (colours) in the sensitivity analysis (dashed) and central analysis (solid line). We identified minor sensitivities to national orphanhood prevalence estimates up to 2007, with no impact on incidence and prevalence estimates for recent years.

Second, we used data from the ‘Topical survey’ of the National Survey of Children’s Health (NSCH) [US Census Bureau, 2023a], filtered to respondents reporting to be grandparents and living with at least one child in the household. Respondents were asked to report on demographic characteristics including the age and race & ethnicity of one randomly selected child. We pooled these data across six years from 2016 to 2021 to characterise the age composition of children who live with grandparents due to small sample sizes in each survey after filtering (approximately 2, 500 grandparents per survey round). Supplementary Fig. S22A compares the age compositions used in the central analysis to those in the two sensitivity analyses. The comparison of the grandparent caregivers prevalence estimates are shown between the central estimate and two sensitivity analyses by age of child in Supplementary Fig. S22B. We found that grandparent caregiver incidence estimates deviated by up to *±*0.028% and *±*0.032% of the central estimate in the two sensitivity analyses due to the rounding numbers, respectively. Additionally, in the two sensitivity analyses, grandparent caregiver prevalence estimates were respectively 12.0% fewer and 13.6% higher than in the central analysis.

**Supplementary Fig. S21:**
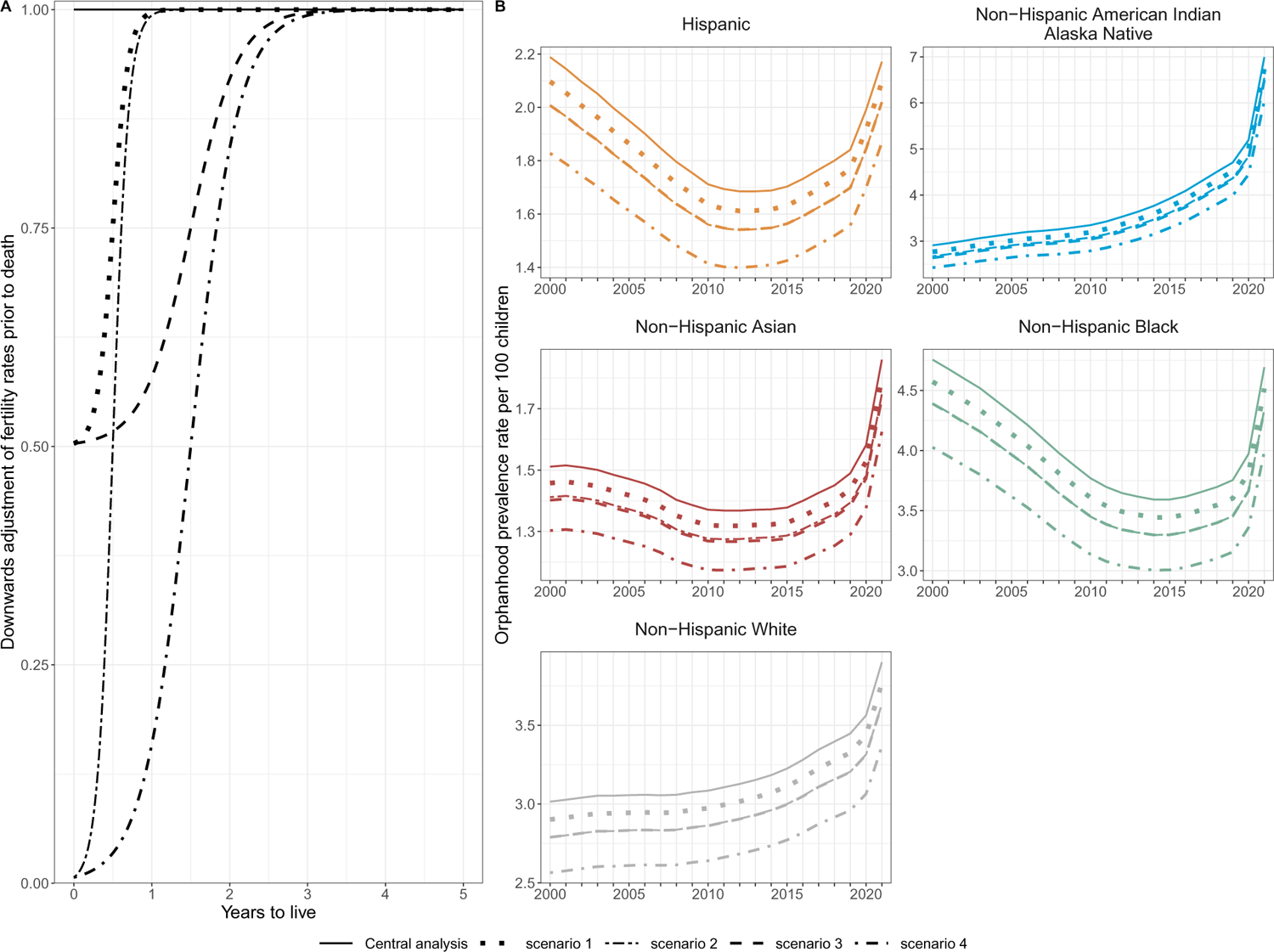
Sensitivity in national-level orphanhood estimates to assumptions on correlations between fertility and mortality rates. (A) We considered four scenarios of lower fertility rates preceding a death event, which we implemented through time-dependent multiplication factors to fertility rates by the time prior to a death event. (B) Estimated national-level orphanhood prevalence rates by race and ethnicity (facets and colours) in the sensitivity analyses (dashed, dotted, dotdash and twodash lines) and central analysis (solid line).

**Supplementary Fig. S22:**
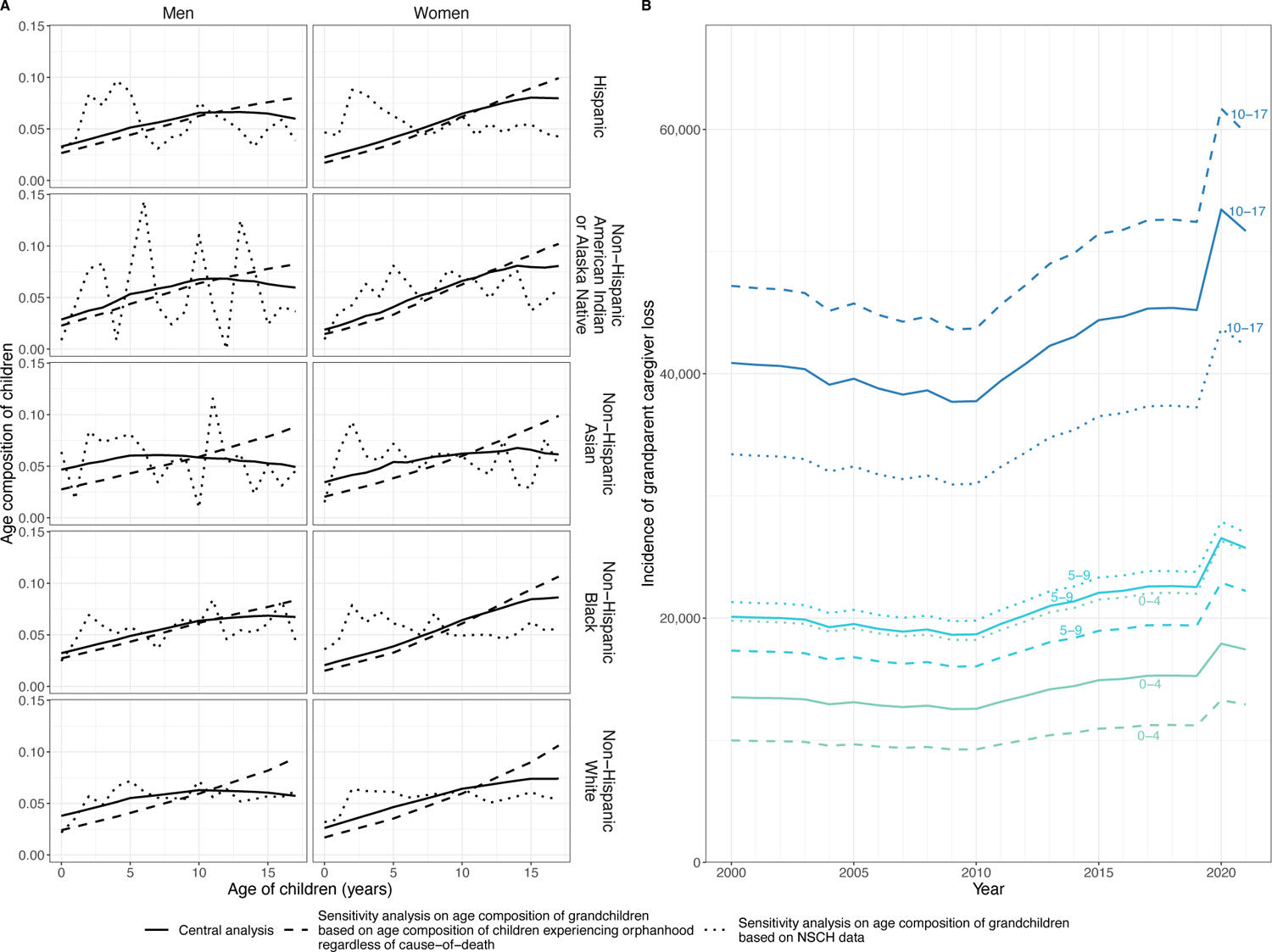
Sensitivity in national-level grandparent caregiver loss estimates to assumptions on the age of children experiencing loss of a grandparent caregiver. (A) Proxies for the age distribution of children who lost grandparent caregivers in two sensitivity analyses. In the first sensitivity analysis (dashed line), we computed the age distribution of children experiencing the death of a parent aged 30+ years across all causes-of-death and by race & ethnicity of the decedent parent. In the second sensitivity analysis (dotted line), we computed the age distribution of randomly selected children living with a grandparent caregiver by race & ethnicity of the child from NSCH surveys in 2016-2021. For comparison, we show the age distribution of children experiencing the death of a parent aged 30+ years due to drug overdose (solid line). (B) Estimated national-level grandparent caregivers loss incidence by age of child (colours) in the two sensitivity analyses (dashed, dotted line) and central analysis (solid line). Labels in colour represent the age of child.

### Sensitivity in national-level caregiver loss estimates to assumptions on historic numbers of grandparent caregivers

In the central analysis, we assumed that the proportion of adults aged 30+ years who live with their grandchildren of age 17 or under (denoted *γ_y,s,r_*) was the same in the years *y* = 1983*, …,* 2009 as in 2010. This assumption was made because data were not available prior to 2010. To investigate the implications of this assumption, we considered longitudinal United Nations Population Division data on Households and Living Arrangments of Older Persons for the U.S. since 1960 United [2022]. These data indicate that the proportion of older persons who live with children or who are the primary caregivers of children has in the U.S. remained fairly constant since 1980 (Supplementary Fig. S23), and for this reason suggests that our assumptions on the historic number of grandparent caregivers is unlikely to have a substantive impact on caregiver loss estimates.

**Supplementary Fig. S23:**
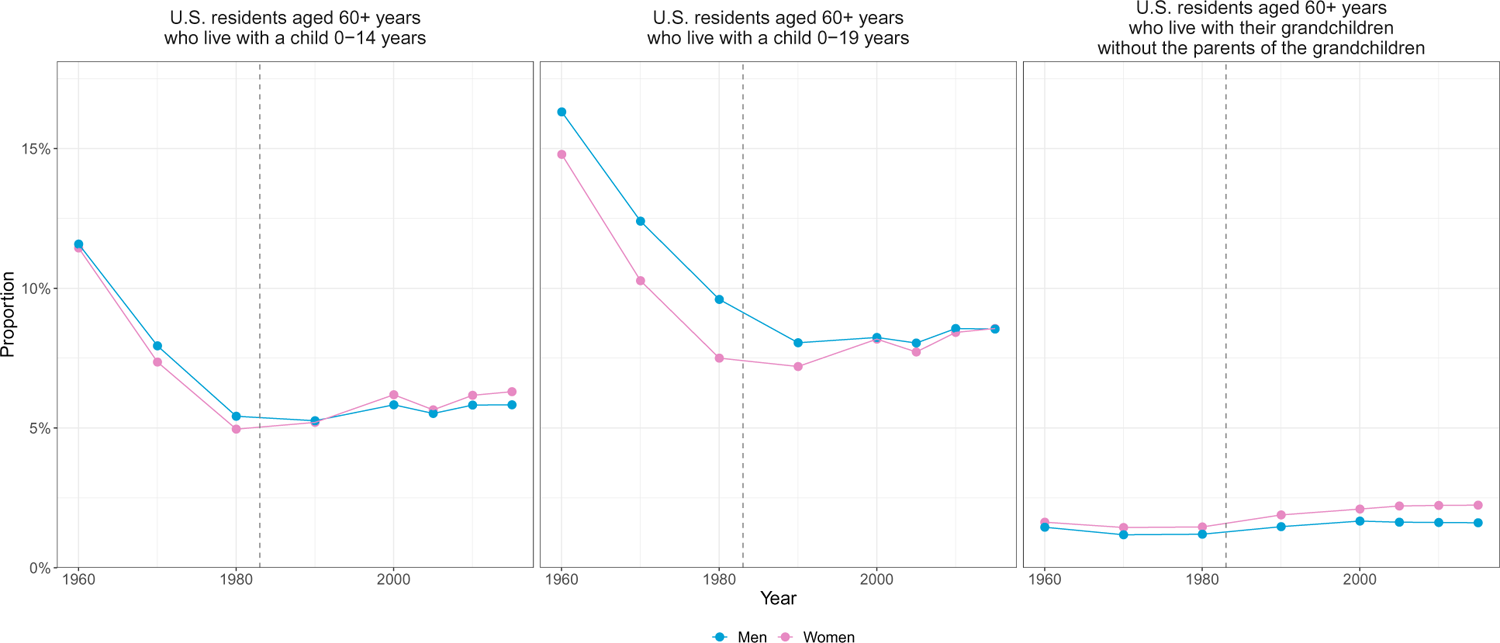
Time trends in the proportion of U.S. residents older than 60 years who are living or caring for children. Data were obtained from United Nations Population Division, data on Households and Living Arrangments of Older Persons between 1960 and 2015. The dashed line represents the year since when grandparent caregiver loss estimates were generated, 1983.

